# Why are girls more distressed than boys? A multi-domain framework applied to longitudinal data across four countries

**DOI:** 10.64898/2026.10.15.26365610

**Authors:** Steven Allsopp, Thomas Steare, Vanessa Rojas Arangoitia, Barkot Milkias, Revathi Ellanki, Hai-Anh Dang, Ola Demkowicz, Praveetha Patalay

**Affiliations:** Unit for Lifelong Health and Ageing, University College London, UK; Centre for Economic and Social Studies, Hyderabad, Telangana, India; Grupo de Análisis para el Desarrollo, Perú; Department of Psychiatry, College of Health Sciences, Addis Ababa University, Addis Ababa, Ethiopia; Living Standards Measurement Study, Development Data Group, World Bank, Washington, District of Columbia, USA; Paul O’Neill School of Public and Environmental Affairs, Indiana University, Bloomington, Indiana, USA; International Inequalities Institute, London School of Economics and Political Science, London, UK; IZA – Institute of Labor Economics, Bonn, Germany; Manchester Institute of Education, The University of Manchester, Manchester, UK; Centre for Longitudinal Studies, Social Research Institute, University College London, London, UK

**Keywords:** Adolescent mental health, Gender differences, Low– and middle-income countries, Social determinants of mental health, Young Lives

## Abstract

**Background:** Girls are more likely than boys to experience psychological distress during adolescence. Drivers of this gender gap remain under-studied, despite its large impact on later life and outcomes, especially for women. Explanatory factors can be gendered in their prevalence (differential exposure) and/or in their impact (differential susceptibility). We developed a multi-domain framework of explanatory factors for distress and applied it in four low– and middle-income countries.

**Methods:** We first proposed a framework of explanatory factors for psychological distress grouped into seven domains (biological, cognitive, psychological, behavioural, social, environmental and broader societal). We applied this framework to identify explanatory variables assessed in childhood and adolescence in the Young Lives longitudinal cohort studies in Ethiopia (N=908), India (N=952), Peru (N=634) and Vietnam (N=887). Distress symptoms at 18-19 years were measured with the Strengths and Difficulties Questionnaire Emotional Symptoms scale. Gendered prevalence of each explanatory factor was tested with Mann-Whitney tests, and gendered impact with country-specific linear regressions including a gender-by-factor interaction term. Exploratory boosted-tree machine-learning models, interpreted with SHAP values and permutation feature importance, examined the contribution of these factors to the gender gap in each country.

**Results:** Distress was higher in girls than boys in every country, the gap ranging from 0.62 points in Ethiopia and Vietnam to 1.76 in India and 2.45 in Peru. Some explanatory factors were consistently gendered in prevalence (e.g. girls experienced greater role overload in each country), while others were gendered only in some countries (e.g. study amount was higher for girls in Peru and Vietnam, but not Ethiopia or India). Gendered impacts on distress displayed more variability across countries. For example, higher self-esteem was more protective for boys than girls in India, Peru and Vietnam, but more protective for girls in Ethiopia. The machine-learning models identified important predictors across all framework domains, though their salience varied by country.

**Conclusions:** Our findings underscore the importance of considering multi-domain explanations for mental-health outcomes and observed inequalities in mental health. Large cross-country differences in the gender gap and in the prevalence and influence of explanatory factors caution against generalising drivers of mental-health outcomes and inequalities across contexts.

## Introduction

Depression and anxiety are leading causes of disease burden worldwide. The World Health Organization estimated that approximately 280 million people globally experienced depression in 2019 (World Health Organization, 2025). These conditions have a substantial gender gap in their prevalence across the life-course that emerges in adolescence (Keyes and Platt, 2024), with approximately twice as many females experiencing depression as males during their lifetime (Salk, Hyde and Abramson, 2017). There is also a considerable gender gap in the distribution of psychological distress symptom levels that underlies the differences observed in the prevalence of conditions such as anxiety and depression (Patalay and Fitzsimons, 2021). The explanatory factors underlying the gender gap are still not fully understood, however there is considerable cross-context variation, with the gender gap varying markedly between countries (Li et al., 2023, Campbell et al., 2021), highlighting the relevance of more than just biological explanations. Understanding the factors which drive the gender gap in psychological distress, their distribution across genders during adolescence and how they vary between countries has the potential to reveal actions to help reduce the overall levels of these conditions as well as the gender gap.

### Gendered prevalence and gendered impacts

Any explanatory factor for the gender gap in psychological distress can either: 1) vary in its prevalence or distribution (differential exposure) making it a gendered risk factor; and/or 2) vary in its association with mental health across genders, reflecting differential vulnerability to the risk factor. In other words, risk factors can have gendered prevalence and/or gendered impacts. An example of a gendered risk factor is sexual violence. Bentivegna and Patalay (2022) found that while exposure to sexual violence resulted in elevated psychological distress in both adolescent boys and girls, girls were five times more likely to experience sexual assault than boys. An example of a non-gendered risk factor with gendered effects includes bullying victimisation. Morken et al. (2023) found that while both the adolescent boys and girls in their sample were equally exposed to bullying victimisation, the impact of bullying on subsequent depression was stronger in girls than in boys.

### Framework of potential explanatory factors for the gender gap in distress

Several authors have proposed frameworks of explanatory factors contributing to the gender gap in depression (Hyde and Mezulis, 2020; Hyde et al., 2008; Kuehner, 2017; Piccinelli and Wilkinson, 2000). These existing frameworks are limited because they restrict their focus to particular domains, have a limited focus on wider societal factors and exclude considerations outside those relevant predominantly to Western, high-income countries (e.g. child marriage). For instance, a recent framework by Kuehner (2017) predominantly focuses on biological, psychological and individual level environmental factors with a brief mention of broader societal factors such as structural gender inequality. An older framework of explanatory factors by Piccinelli and Wilkinson (2000) lists childhood familial environment and adverse childhood experiences, social roles and cultural norms, adverse life events, vulnerability and coping style, social support, genetic factors, hormones and neurotransmitter systems. They note the limited evidence available for most of these factors and highlight the need for greater research focused on understanding the drivers of the observed gender gap in these conditions.

Hyde and Mezulis (2020) apply an ABC model of vulnerability-stress with vulnerabilities categorised into affective, biological and cognitive (ABC). They list temperament and negative emotionality as relevant affective factors, genetic, hormonal and pubertal timing as biological factors and negative cognitive style, objectified body consciousness and rumination as cognitive factors. They also posit that negative life events and stress play a major role and end with a brief overview of sociocultural issues such as structural gender inequalities and culture, with a focus on media and social media culture.

The potential importance of artefactual or measurement issues such as gendered patterns in tendency to and ways of reporting distress, healthcare-seeking behaviour, recall differences or symptom profiles that might make it seem like there is a gender difference in these outcomes is also covered by existing frameworks (Kuehner, 2017; Piccinelli and Wilkinson, 2000). However, they highlight that the evidence for these artefactual explanations is also limited.

In this study, we first aimed to address some of these limitations in the existing frameworks by creating a comprehensive framework that includes a wide range of explanatory factors for the gender gap in adolescent psychological distress from multiple domains. We subsequently applied this framework to analyse data from countries where adolescent psychological distress has been understudied to-date.

### Proposed Framework

We propose a comprehensive framework of factors with the potential to explain the gender gap in psychological distress. We draw upon and consolidate several existing frameworks such as those by Kuehner (2017); Piccinelli and Wilkinson (2000); Hyde et al. (2008) and Hyde & Mezulis (2020) and also include other potential explanatory factors from existing empirical research findings and expert consultation. We include a new behavioural domain, which most prior frameworks excluded, as there is evidence that health-related behaviours, their social costs and healthcare seeking behaviours are gendered in many societies (Thompson et al., 2016, Das et al., 2018). We also focus more on social and broader societal factors including gender norms, gender inequalities and gendered risk factors that are relevant in a broader global context, such as gender preferences, sex-selective abortions, child marriage, female genital mutilation.

We present this framework in Figure 1, where potential explanatory factors are grouped into biological, reporting artefacts, cognitive, psychological, behavioural, social, environmental and broader societal domains. The explanatory factors included are typically experiences that are causes of anxiety and depression which girls experience more often than boys, or experiences that might not vary across genders in their prevalence but are evidenced or theorised to have a differential impact on mental health outcomes.

**Figure 1:**
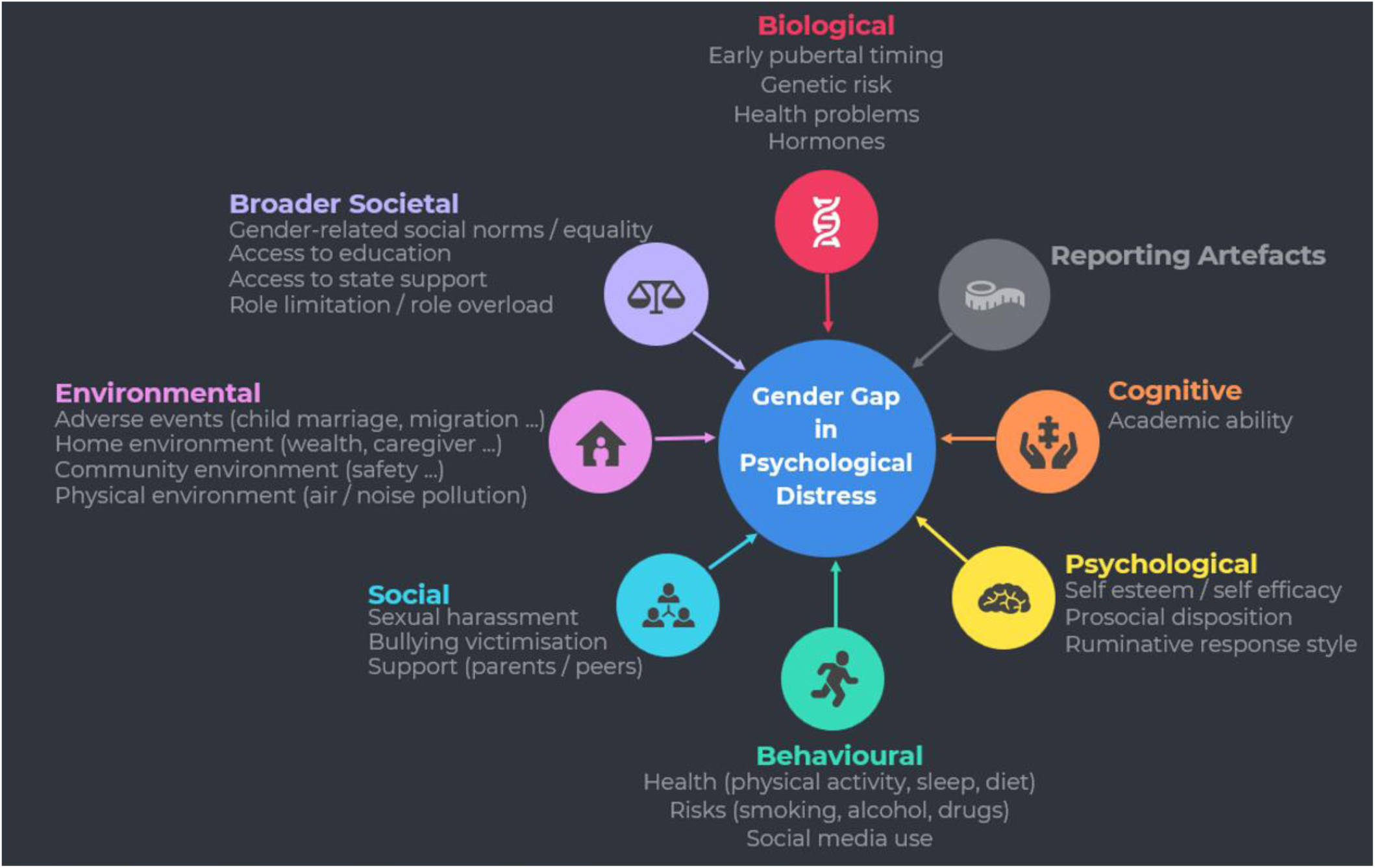
Overview of the proposed framework of explanatory factors for the gender gap in adolescent psychological distress.

While there is limited evidence for the impact of genetic factors on the gender gap, *biological factors*, including hormonal changes, early pubertal timing and stress response, have been associated with the emergence of the gender gap in adolescence (Kuehner, 2017). *Cognitive factors* have also been linked to the gender gap in psychological distress. For instance, there is evidence that lower academic performance can be predictive of experiencing depressive symptoms in adolescents. Yong et al. (2022) observed a gender difference in this association, with poor GPA scores predicting depressive symptoms only in girls. There is also evidence of gender differences in some cognitive indicators, such as maths scores, however these differences have been found to vary between countries, with the broader societal context in a country affecting the gender difference. For example, more gender-equitable countries have a smaller gender gap in maths performance (Else-Quest et al., 2010). Other researchers have hypothesised that *psychological factors* including cognitive style, such as tendency to ruminate, and affective factors, such as emotional reactivity, interact with biological factors and adverse life events to influence the gender gap (Hyde et al., 2008; Hyde and Mezulis, 2020). For example, rumination, and in particular the tendency to passively and repetitively reflect on negative events, not only has a gender difference in frequency, with more adolescent girls than boys having a ruminative response style, but has also been found to partially mediate the relationship between gender and depression (Espinosa et al., 2022, Treynor et al., 2003). A number of studies have identified *behavioural factors*, which display gender differences both in prevalence and association with depression, including smoking (Wilkinson et al., 2016) and alcohol consumption (Jeong et al., 2019). For example, Sun et al. (2023) found that starting in adolescence, disrupted sleep is more common in girls than in boys, while sleep disorders are more predictive of depression in adolescent girls than in boys. *Social factors* such as social pressure from peers and parents, as well as cultural norms can result in children conforming to traditional gender roles and multiple expectations about femininity as they move through adolescence, which leads to role limitation or role overload in girls as they have to balance household and caring responsibilities with study and work. These factors have been linked to increased stress and higher prevalence of depression in girls (Nolen-Hoeksema, 2001). Tiggemann and Slater (2015) found that self-objectification, whereby adolescent girls internalise the culturally pervasive sexual objectification of women’s bodies (Fredrickson and Roberts, 1997), was predictive of depressive symptoms. The authors also concluded that other social factors, including media consumption and peer interactions about appearance were predictive of self-objectification. Demkowicz et al. (2025) also found evidence that for adolescent girls’ experiences of social media can amplify gendered expectations of appearance and behaviour, while encouraging comparison to peers and other public figures.

*Environmental factors* include those related to the family, home, community and physical environments in which children grow up. While adverse childhood experiences (ACEs) within this domain have been consistently linked to depression and anxiety in adolescents (Elmore and Crouch, 2020), there is less consensus on the relationship between ACEs and gender differences in psychological distress, partly due to inconsistent definitions of what constitutes an ACE (Keyes and Platt, 2024). Some ACEs differ in their prevalence between genders, for example, forced marriage is more common among girls, while exposure to physical violence is more common among boys. Gajos et al. (2022) found evidence of gender differences in the association between exposure to ACEs and subsequent experience of psychological distress in adolescence, however they also observed gender differences in the impact of protective factors such as parental support, suggesting a complex relationship between gender, ACEs, protective factors and psychological distress.

*Broader societal factors* can include anything that is impacted by structural gender inequalities and also economic factors such as income inequality, wealth and poverty levels, which have been found to be associated with the gender gap in psychological distress, although findings about the direction and nature of these impacts remain unclear (Campbell et al., 2021). For example, Salk et al. (2017), counterintuitively, found evidence of a larger gender gap in depression in countries with greater gender equity, whereas other studies have reported that greater macro-level gender equality was associated with smaller gender gaps in depression (Van de Velde et al., 2013). Of note, most of this research is limited to higher income country settings and wider gender equality issues such as access to education, male preference, sex-selective abortions, and child marriage have not been included in prior frameworks.

### Context and gender inequality

Wider societal expectations, and the ways difficulties are framed in children’s key contexts such as schools, are highly gendered, shaping how gender inequalities in mental health emerge and are maintained. For example, girls are celebrated for their emotional openness and maturity while being positioned as unfairly advantaged by the perceived receipt of more mental health support (Stentiford et al., 2025).

Political, geographical and historical contexts provide important settings for how contextually specific gendered norms and structural inequalities emerge and operate. In the current study, we analyse data from four countries outside of Europe and North America – Ethiopia, India, Peru and Vietnam. While none of the four is a high-income country, they are vastly different in their geographies and histories and in the roles of and pressures on women, which are themselves changing over time. In terms of political participation, Ethiopia has 41% of seats in parliament held by women, followed by Peru (38%), Vietnam (31%), and India (14%) (World Bank Group, 2024). However, representation varies significantly between national and local levels across all countries. Economic participation also varies greatly with female labour force participation in Vietnam being around 69%, followed by Peru (65%), Ethiopia (57%) and India (33%) (World Bank Group, 2024). In all four countries more of women’s labour is informal labour and gender wage gaps still exist. Each country’s gender (in)equality reflects unique social and historical influences. For instance, historically women have had more freedom in Vietnam compared to neighbouring countries with a similar culture, and alongside socialism for several decades, this might have resulted in greater female labour force participation that persists to this day. On the other hand, India still has some of the world’s highest rates of sex-selective abortions, female infanticide and child marriage globally, with many barriers to women’s labour market participation. The 2023 scores on the Gender Inequality Index created by the United Nations Development Programme (UNDP) also vary (lower score indicates better gender equality), Ethiopia (0.497), India (0.403), Peru (0.34), Vietnam (0.299) (UNDP, 2023).

### Current study

While some studies have explored the factors underlying the gender gap in distress, the causes of the gender gap remain understudied (Patalay and Demkowicz, 2023). The focus of the little existing research has predominantly been on high income, White-majority countries, with low and middle income (LMIC) countries remaining relatively under-researched (Salk et al., 2017; Sun et al., 2023), despite accounting for around 90% of the world’s adolescents.

In this study, we used data from four diverse LMICs, Ethiopia, India, Peru and Vietnam, to examine the extent and drivers of the gender gap in psychological distress in adolescents in each country. Using our framework to guide variable selection, we examined potential explanatory factors from the biological, cognitive, psychological, social, environmental, behavioural and broader societal domains. For each factor we examined, by gender, the prevalence in childhood and early adolescence (between ages of 7 and 15 years) and longitudinal associations with adolescent psychological distress at age 18/19 years. By looking at each country separately, we hoped to derive insights into the variability in prevalence of risk factors and associations by gender across different contexts.

Our primary research objectives were: (a) to understand the magnitude of the gender gap in adolescent psychological distress in each country; (b) for each explanatory factor in our proposed framework, to determine whether the prevalence of the factor differed across boys and girls (i.e. was the risk factor gendered) or whether its association with psychological distress differed across genders (i.e. a non-gendered risk factor which had gendered effects); (c) to explore important factors for adolescent psychological distress and whether these are similar across boys and girls.

For all objectives we also aimed to describe country differences and examine whether the gender gap in adolescent psychological distress, the predictive factors, their prevalence and association with mental health differed between the four countries.

## Data

### Methods

We used data from the older cohort of the Young Lives study. Young Lives is an ongoing longitudinal cohort study of poverty and inequality, which since 2001 has been following the lives of children in four LMICs – Ethiopia, India, Peru and Vietnam. In each country the children were chosen from 20 locations, both rural and urban. The locations were identified non-randomly, excluding wealthy locations and over-sampling poorer ones. Within each location, the children were selected randomly. While children from poorer locations and families were over-represented in the sample, the study aimed to select children from a range of backgrounds. The Young Lives study seeks to apply a comparative design, with consistent core questions being asked in each country and across rounds, albeit with questions evolving to reflect the change in age of the children at each round.

In this study, we used data collected in the first four waves of the Young Lives study, starting with round 1 in 2002 when children were aged 7.5-8.5 years (Ethiopia N=1,000; India N=1,000; Peru N=714; Vietnam N=1,000) and ending when they were aged 18-19 years in round 4 in 2013 (Ethiopia N=908; India N=952; Peru N=634; Vietnam N=887). Intermediate rounds were conducted in 2006 (round 2) and 2009 (round 3). The participant numbers quoted above reflect both attrition and participants excluded due to insufficient data being available. Attrition rates in the study were relatively low, with more than 91% of the original participants still being involved in the fourth round of the study. The data and analysis included in this paper is based upon participants who were still present at round 4 of the study (N=3,381).

### Measures

Young Lives is a multi-disciplinary study, and a range of data were gathered from the children directly, from caregivers and other household members as well as data on the community contexts in which each child lived. Data collection was coordinated by local research teams in each of the four countries. The data cover a range of outcome variables, including the health and emotional wellbeing of the children along with a broad spectrum of potential explanatory factors relevant to the biological, cognitive, psychological, behavioural, social, environmental and broader societal domains in our framework (shown in Supplementary Figure S1).

In this study we focused on binary gender, based upon recorded biological sex, given the lack of more detailed gender identity data. This information was recorded based upon interviews conducted with an adult in the child’s household at the very start of the Young Lives study when the children were aged 8 years. The interviewees were asked for the gender of the child and were just given two options to choose from – male or female. We adopted the terms ‘girls’ and ‘boys’ in line with gender rather than sex-based language and have sought to avoid implying attributions to sex and biological difference.

### Psychological distress

Distress symptoms were measured using the Emotional Symptoms scale of the Strengths and Difficulties Questionnaire (SDQ) (Goodman, 1997) administered to participants when they were aged between 18 and 19 years. Participants responded to five statements (see Supplementary Data 1) each describing an emotional difficulty, with which participants had to indicate the strength of their agreement or disagreement. This resulted in a score between 0 and 15 for each participant.

### Explanatory factors

Supplementary Figure S1 displays the potential explanatory factors in our framework for which data were available in the Young Lives study, while Supplementary Table S1 sets out the rationale for their inclusion in the framework. Descriptions of how each of these factors is measured are shown in Table 1, while further details of the data fields used to construct each factor are included in Supplementary Table S2.

**Table 1:**
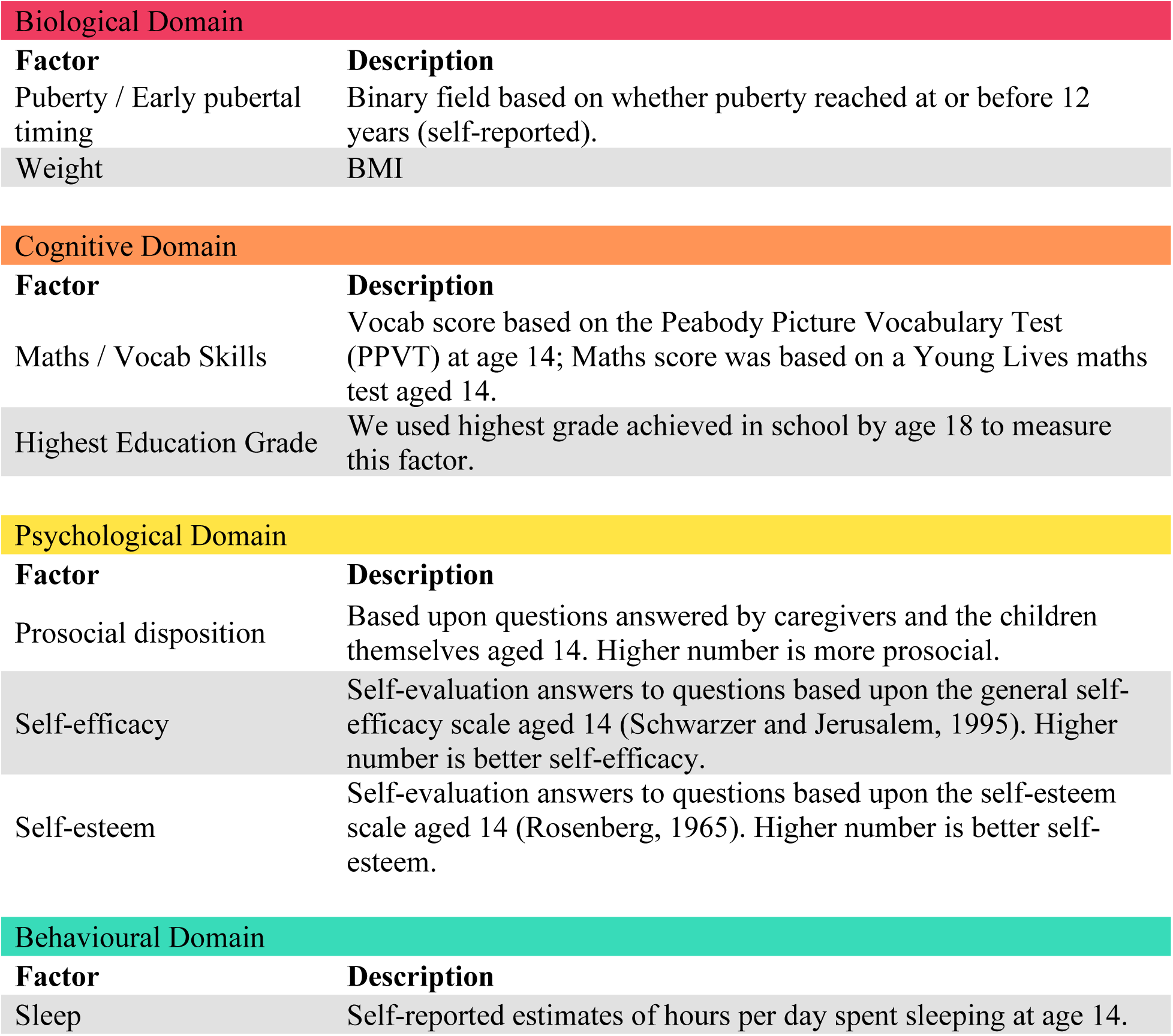
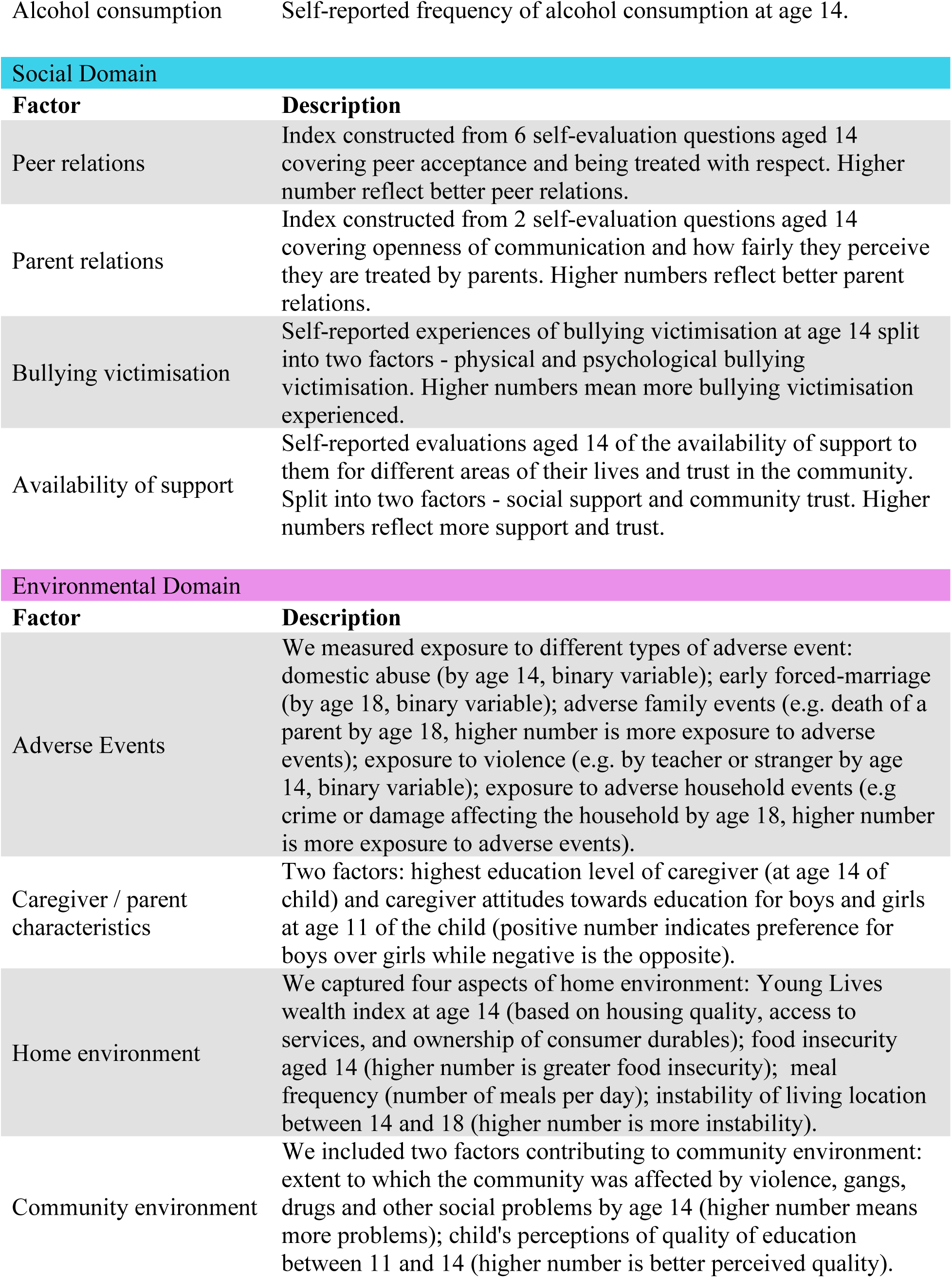
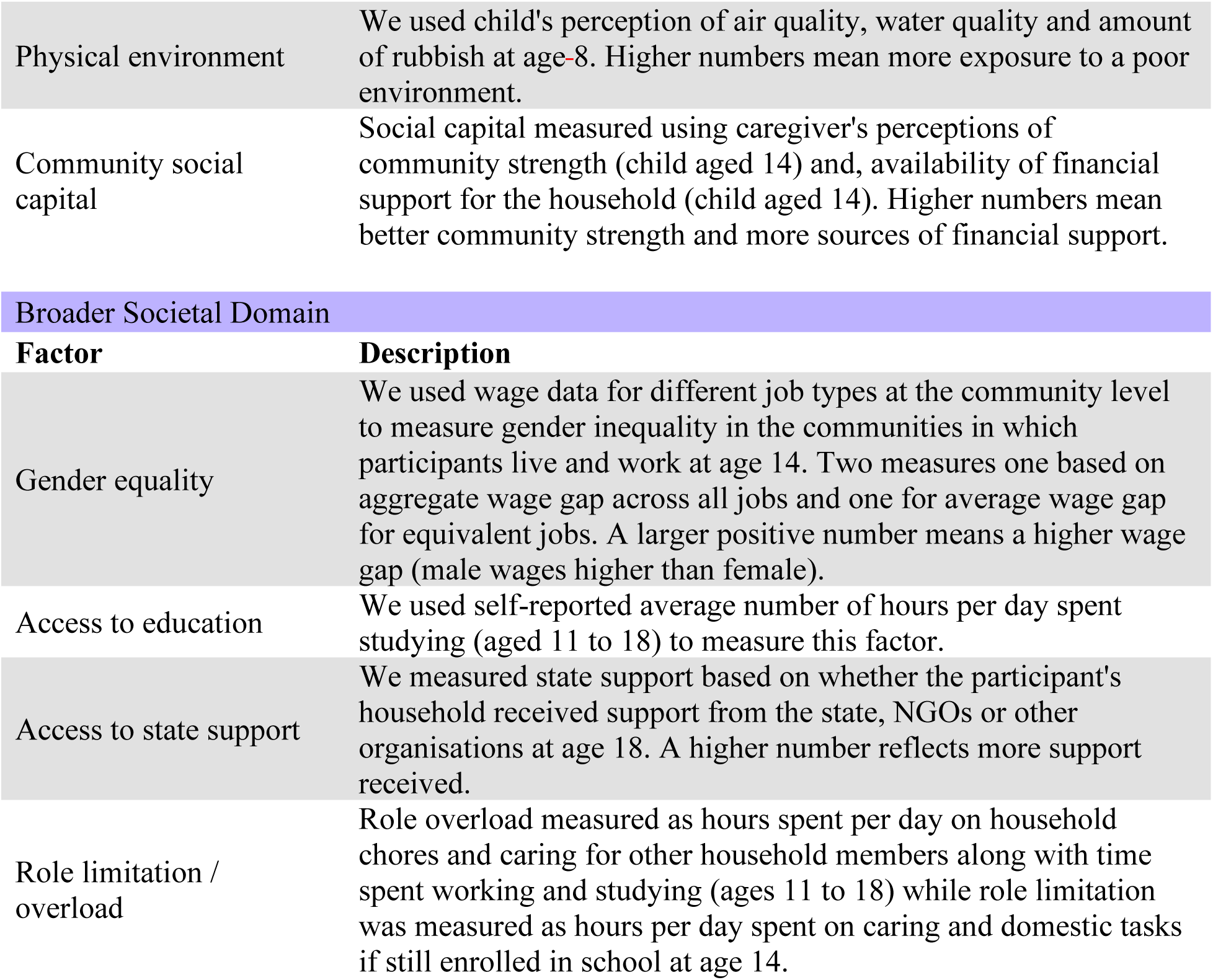
Explanatory factors from our framework available in the Young Lives data and used in our quantitative analysis.

We were not able to include all relevant factors from our conceptual framework due to data availability. For example, no appropriate data were available to capture genetic risk within the biological domain and social media use within the behavioural domain. Some relevant factors were only assessed in one country (e.g. substance use was only assessed in Peru). However, this is to be expected from data analyses of existing cohorts, and the current study still presents the most comprehensive multi-factor analyses undertaken on this question to-date.

### Analytic strategy

Data were analysed using RStudio (R4.4.0).

We first described the gender gap in psychological distress in late adolescence (age 18-19 years) by presenting differences in means and distributions by gender and by country. Next, we analysed the gender gap in each explanatory factor by country, using Mann-Whitney tests to identify significant gender differences (at 0.05 significance). We then used regression analysis to estimate the association of each individual explanatory factor with psychological distress in each country and for each gender. These regressions took the form y = x_i_ + e, where y is psychological distress and the x_i_ are the individual explanatory factors. To identify whether the association between each explanatory factor and psychological distress was gendered, we performed another series of regression analyses, separately for each country, with gender, explanatory variable and interaction between gender and explanatory variable as independent variables. These regressions took the form y = gender + x_i_ + gender * x_i_ + e. For models where there was evidence of a potential gender difference in association with mental health (based on interaction p<0.10), we illustrate the interaction using model predicted marginal means. In the above analyses, we consciously considered single-factor associations rather than using mutually-adjusted models. This is because many of the factors in our framework are not independent, for example the broader societal factors will shape some of the individual behavioural and psychological factors, hence mutual adjustment would risk conditioning on mediators and obscuring the associations of interest. Given the breadth of factors, countries and genders examined, our analyses involve substantial multiple testing, and we applied no formal correction because of the exploratory rather than hypothesis confirming nature of the study. We treated the p<0.10 interaction threshold as a deliberately sensitive screen suitable for the exploratory objective of identifying candidate gendered effects, rather than as confirmatory inference. Missing data on each factor was low, and hence for every model available data on each factor is used.

Finally, we conducted a more exploratory piece of analysis by fitting ensemble boosted tree models, using the XGBoost package in R (Chen and Guestrin, 2016), to the data for each country in order to predict psychological distress using the explanatory factors outlined above as well as the interactions of those factors with gender. To avoid loss of sample due to missingness on any one factor, we use imputation to ensure the full analytic sample is retained in these models. Tree boosting is a supervised machine-learning method, which sequentially trains an ensemble of predictive decision trees, each learning from the residual errors of the previous trees. We used SHAP values (Shapley additive explanations) and Permutation Feature Importance (PFI) measures to determine the relative contribution of the different explanatory factors and their gender interactions to the model predictions and model performance respectively. SHAP values (Lundberg et al., 2020) quantify the change in expected model prediction resulting from each explanatory factor in each prediction. They also allow us to quantify interaction effects between explanatory factors. PFI (Breiman, 2001), by contrast, measures the increase in model prediction error which results from randomly permuting the values of each explanatory factor. The use of ensemble boosted tree models with SHAP and PFI complements the regression models which form the analytical backbone of the paper. The tree models, as non-parametric models, make no assumptions about linearity or additivity, and can detect which explanatory factors are most associated with the dependent variable (psychological distress) even when their relationships are complex or context dependent.

## Results

Adolescent psychological distress at age 18/19 was higher in girls than in boys in each of the four countries. The size of the gender gap differed between countries with mean symptoms greater in girls by 0.62 (95% CI [0.29,0.95]) in Ethiopia, 1.76 (95% CI [1.48,2.04]) in India, 2.45 (95% CI [1.86,3.03]) in Peru and 0.62 (95% CI [0.34,0.89]) in Vietnam (Figure 2, Supplementary Table S3).

**Figure 2:**
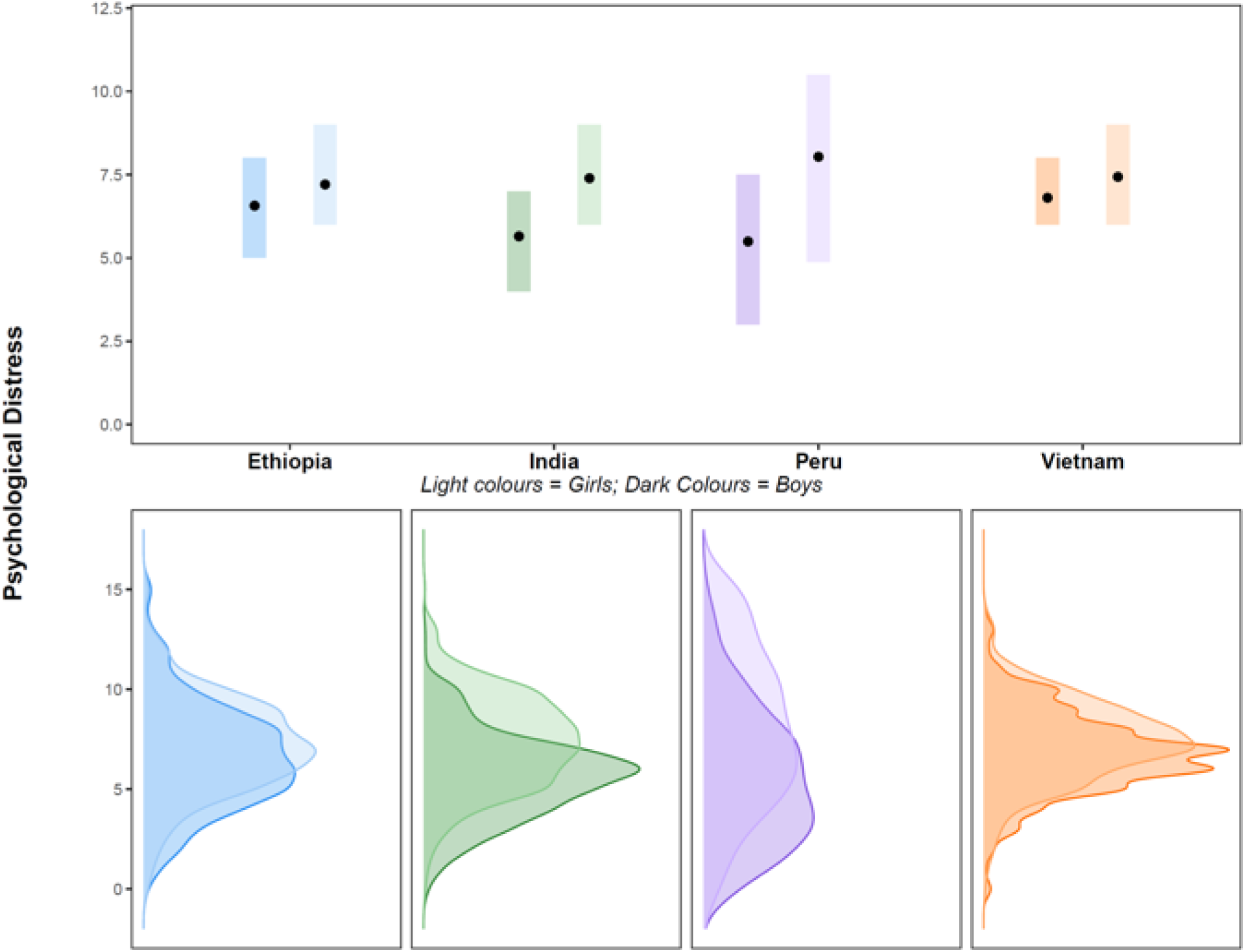
The top panel shows the interquartile range for psychological distress by com1try by gender. Mean psychological distress for girls, displayed by the black dots, is higher in all countries than it is for boys. The lower panel shows the distribution of psychological distress by country by gender.

### Are explanatory factors gendered in their prevalence or distribution?

Table 2 shows the mean values of the explanatory variables by country, highlighting those which differ significantly by gender. Supplementary Figure S2 displays the distributions of these explanatory variables by gender and country. Many of the explanatory factors identified, displayed significant gender differences in their distribution, with some of the differences being consistent across countries, while in other cases there were differences between countries.

**TABLE 2.**
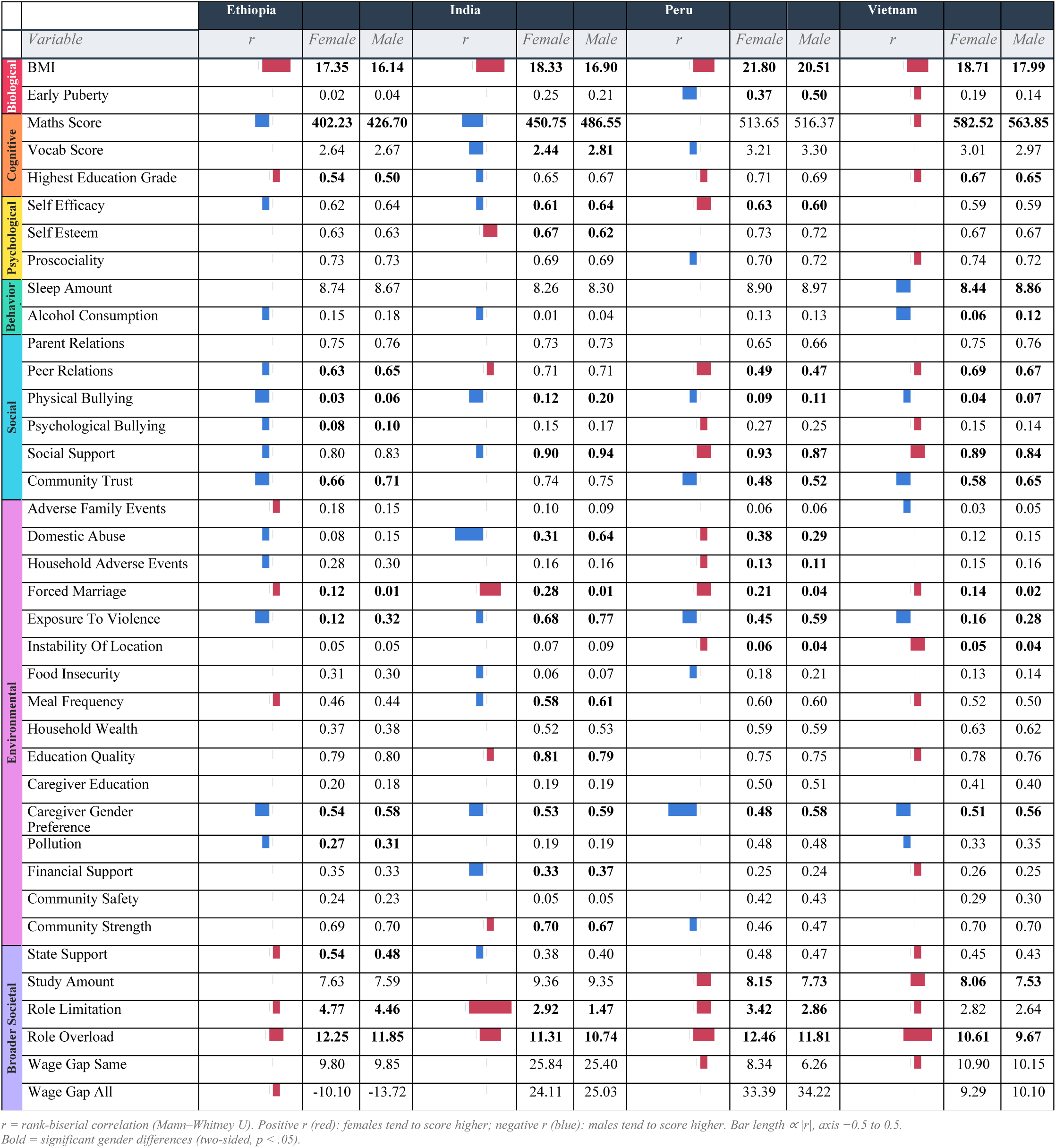
Mean values for each explanatory factor split by gender and country. Bar chart key: Red bar = Female > Male; **Blue bar** = Male > Female. Bar = rank-biserial r (scale 0–0.5). **Bold** cells are those for which the gender difference in the country is significant based on a Mann-Whitney Test at 0.05 significance.

### Are explanatory factors gendered in their associations with subsequent psychological distress?

For each gender (in each country separately), we ran simple linear regressions, with psychological distress as dependent variable and each explanatory factor individually as the independent variable. Figure 3 shows the coefficients, along with their 95% confidence intervals, for each of these regressions. This data is displayed in tabular form in Supplementary Table S4, where the cells in bold are those for which the coefficient is significantly different to zero at 95% confidence.

**Figure 3:**
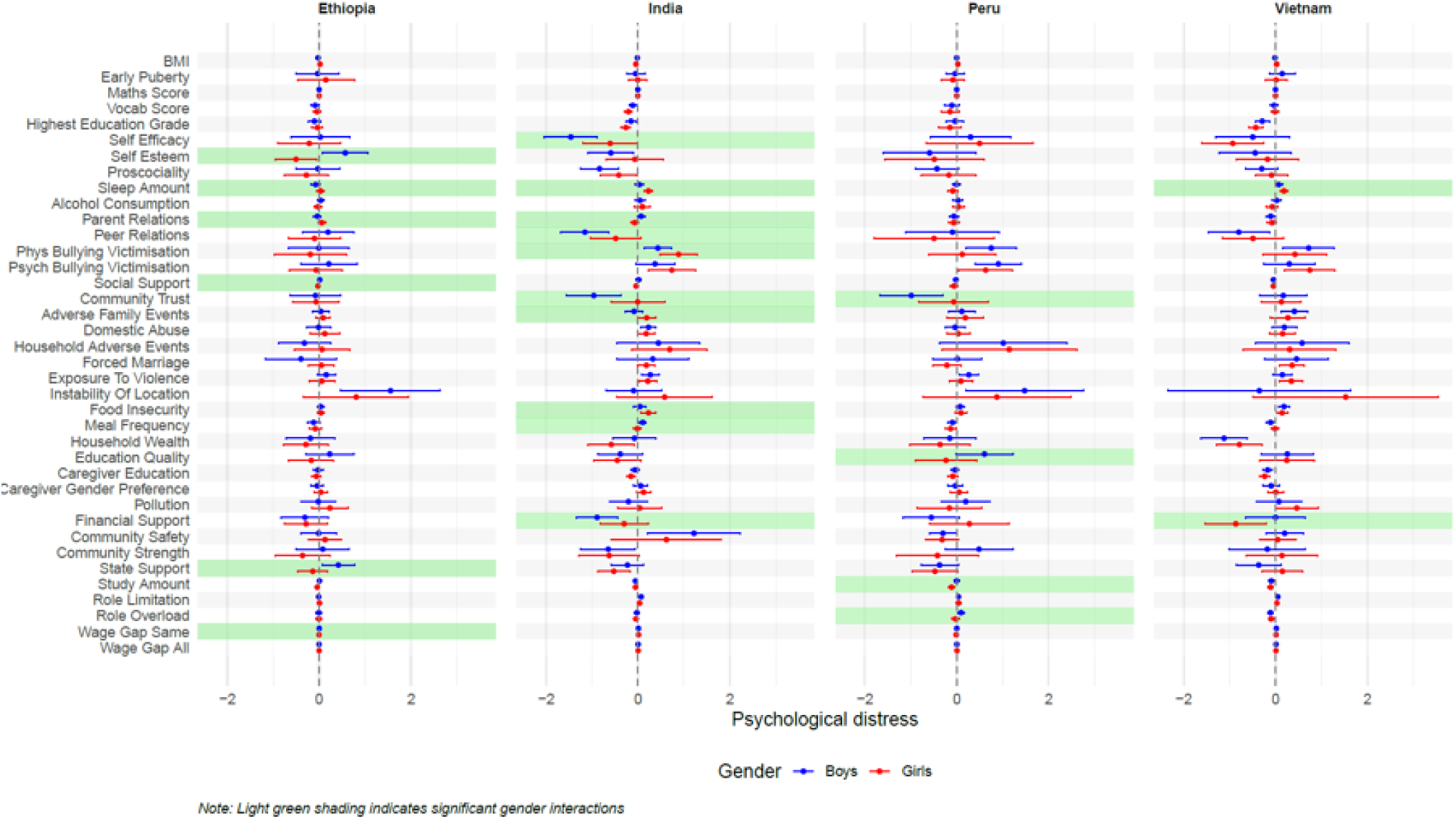
Regression summary. Coefficients and confidence intervals for regressions of each explanatory factor as independent variables and psychological distress as dependent variable. Highlighted cells are those for which the interaction term in regressions of gender and interaction between gender and each explanatory factor as independent variable and psychological distress as dependent variable is different to zero at 0.10 significance.

Many, but not all, of the explanatory factors identified, were significantly associated with distress. A few factors showed consistent associations with subsequent distress outcomes across genders and countries: for instance, cognitive factors such as vocabulary score and highest education grade were consistently associated with lower distress. Most other examined factors showed differences by gender and/or country: for instance, greater community trust was associated with lower distress symptoms in Ethiopia and Peru, higher symptoms in Vietnam and mixed results in India. We discuss the findings by domain area in greater detail in a separate section below.

We formally tested whether the relationship between the explanatory factors and distress differed by gender. For each country individually, we ran simple linear regressions, with distress as the dependent variable and with gender, each explanatory factor, and the interaction between gender and the explanatory factor individually as the three independent variables. Supplementary Table S4 shows highlighted the explanatory factors for which the coefficients of the interaction term are significantly different to zero at 90% confidence. These factors are also highlighted in green in Figure 3 above. We visualise these interactions using predicted margin plots in Supplementary Figure S3.

### Which factors are most predictive of the gender gap in distress?

Finally, we fitted ensemble boosted tree models to predict psychological distress based upon the explanatory factors for each country. Figure 4 displays the relative importance of each factor (Figure 4a) as well as the relative importance of interaction terms between gender and explanatory factors (Figure 4b) for each country.

**Figure 4a:**
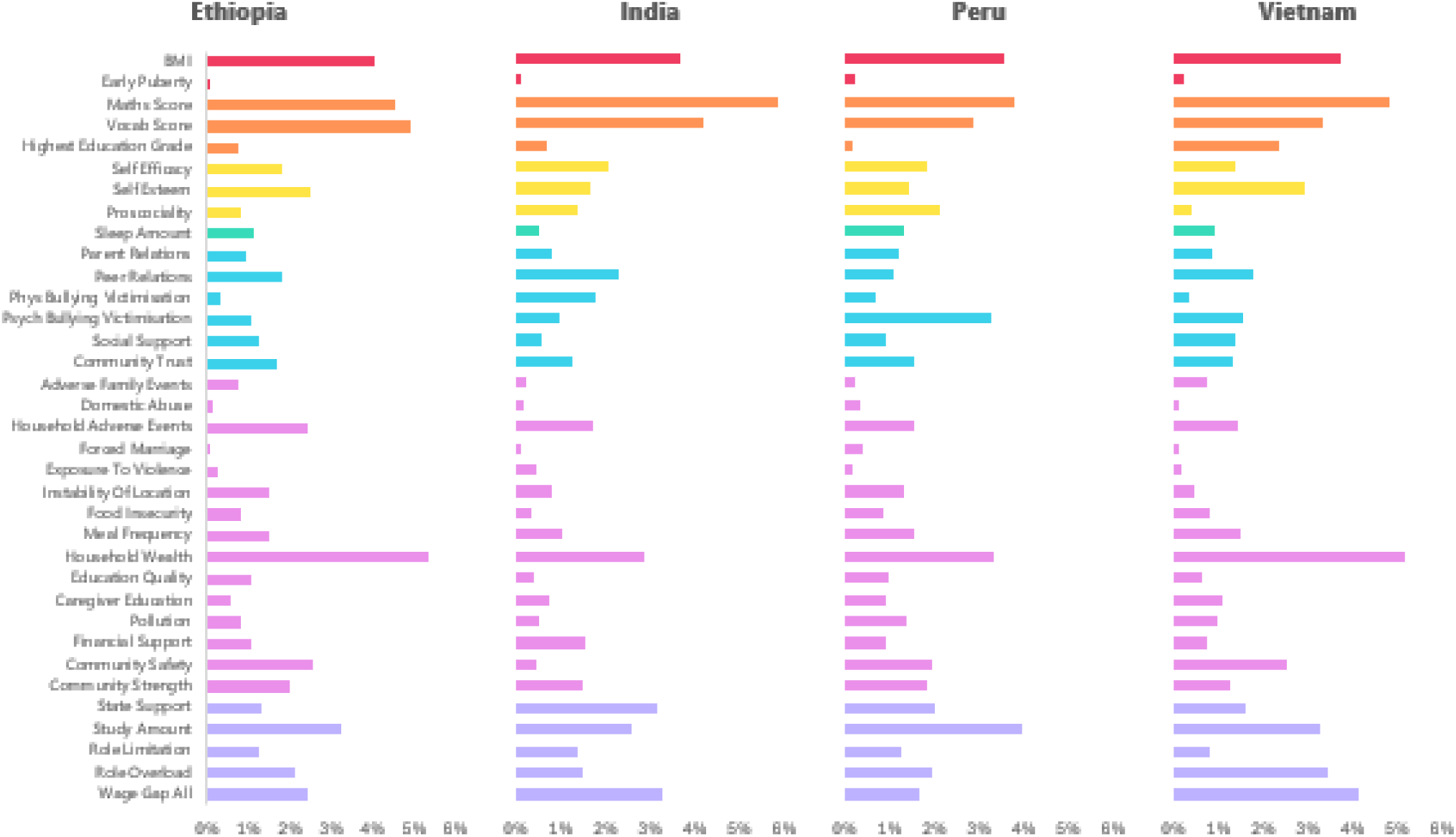
Permutation Feature Importance (PFI) values for the main effects of each explanatory factor by country. PFI equates to the percentage contribution of each factor to the total model performance for that country. In combination with the interaction effects in figure 4b the PFI values for each country sum to 100%.

**Figure 4b:**
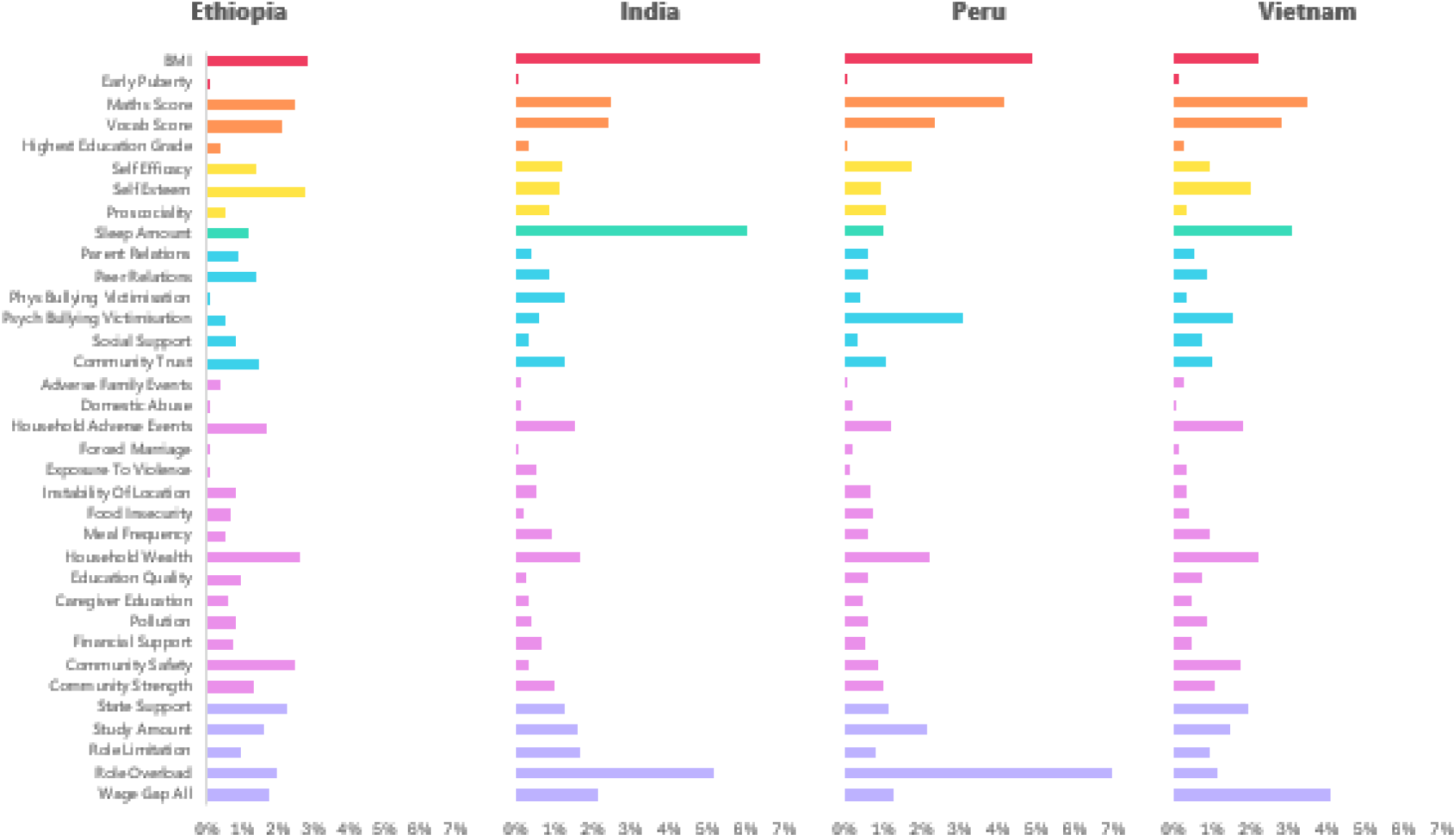
Permutation Feature Importance (PFI) values for the interaction effects of gender with each explanatory factor by country. PFI equates to the percentage contribution of each factor to the total model performance for that country. In combination with the main effects in Figure 4a the PFI values for each country sum to 100%.

### Key findings organised by framework domains

#### Biological Factors

Of the biological factors included, BMI was a more important predictor of psychological distress in the tree models compared to early pubertal timing. BMI was higher in Peru than in the other countries. There were significant gender differences in its distribution with girls having significantly higher BMI than boys in every country. The relationship between BMI and psychological distress was complex, with higher BMI generally being associated with higher psychological distress in girls (except in India where the opposite holds) and generally being associated with lower psychological distress in boys, however, with the exception of the association in Indian girls, none of these relationships were statistically significant.

#### Cognitive Factors

Cognitive factors were consistently important predictors of psychological distress in the tree models for all countries (Figure 4) and also in the regression models (Figure 3), with higher maths and vocabulary abilities being associated with lower psychological distress. There were gender differences in these factors, with boys generally scoring more highly, except in Vietnam where girls outperformed them. The effect of the cognitive factors on psychological distress was not, however, gendered, with both boys and girls typically benefitting from higher cognitive scores. Highest education grade was lowest in Ethiopia. The grade was higher in girls than in boys in Ethiopia and Vietnam. Higher grades were consistently associated with lower psychological distress with a significant relationship for both boys and girls in India and Vietnam.

#### Psychological Factors

Psychological factors, including self-esteem, self-efficacy and prosociality were mostly associated with lower psychological distress. Better self-esteem was generally associated with lower psychological distress, significantly so for boys in India and girls in Ethiopia. Unexpectedly, the opposite relationship was found for boys in Ethiopia, for whom higher self-esteem was associated with greater psychological distress. Better self-efficacy was also generally associated with lower psychological distress (significant in boys in India and girls in India and Vietnam). There was a significant interaction effect in India reflecting a stronger protective effect of higher self-efficacy in boys than in girls (Supplementary Figure S4a). Better prosociality was generally associated with lower psychological distress (significant in boys and girls in India).

#### Behavioural Factors

There was evidence of a gendered impact of sleep on distress, with significant interaction effects in Ethiopia, India and Vietnam reflecting more sleep being associated with higher psychological distress in girls but not boys in these countries. Self-reported alcohol consumption in mid-adolescence was higher for boys than girls in Ethiopia, India and Vietnam, however, we did not observe impacts of alcohol consumption on later distress in any gender across all countries.

#### Social Factors

Social factors fall into two groups, with bullying victimisation (physical and psychological) being associated with higher psychological distress, while peer relations, community trust and social support were generally associated with lower psychological distress. Physical bullying victimisation was more prevalent in boys than in girls in all countries, and highest overall in India. More exposure to physical bullying victimisation was associated with worse psychological distress for boys in India, Peru and Vietnam and girls in India. There was also evidence of a gendered impact in India, where the effect of physical bullying victimisation on psychological distress in girls was higher than in boys. Psychological bullying victimisation was higher in Peru than in other countries. More exposure to psychological bullying victimisation was associated with worse psychological distress for girls in India, Peru and Vietnam and boys in Peru, however there was no evidence of a gendered impact of psychological bullying victimisation on distress.

Community trust was higher in boys than in girls in Ethiopia, Peru and Vietnam. Better community trust was generally associated with lower distress (except in Vietnam), but this was statistically significant only for boys in India and Peru. There was also evidence of a gendered effect where better community trust was associated with lower psychological distress for boys but not girls in India and Peru (Supplementary Figure S4b). Peer relation scores were lower in Peru than in other countries. Better peer relations were generally associated with lower psychological distress (significant in boys in India and Vietnam). There was an interaction effect in India reflecting the above relationship in boys while the relationship in girls, although in the same direction, was not significant.

#### Environmental Factors

Environmental factors were generally important contributors to the boosted tree model’s performance. Household wealth was associated with lower psychological distress in all countries and genders with significant effects for girls in India and Vietnam and boys in Vietnam. There were significant country level differences in household wealth, with Ethiopia having the lowest levels. There were no significant gender differences in the effect of household wealth on psychological distress.

Food insecurity was consistently associated with worse psychological distress in all countries. We observed country level differences, with food insecurity being more prevalent in Ethiopia, and to a lesser extent Peru, than in other countries. While there were no significant gender differences in the prevalence of food insecurity, there were gendered interactions between food insecurity and psychological distress, for example in India, food insecurity had a more adverse effect on psychological distress in girls than in boys (Supplementary Figure S4c).

Other environmental factors such as domestic abuse, forced marriage, exposure to violence, household adverse events, (poor) community safety and (lack of) stability in living location were generally associated with higher psychological distress. There were significant gender and country differences in the prevalence of many of these factors. For example, domestic abuse rates were typically higher in boys except in Peru where rates were higher for girls, while rates in India and Peru were higher than in other countries. Forced marriage rates were consistently higher in girls, and higher in India and Peru. None of these other environmental factors displayed differential effects on psychological distress by gender.

Environmental factors such as community strength and financial support were mostly associated with lower psychological distress. The association of financial support with psychological distress was gendered in some countries, with greater mental health benefits for boys in India and girls in Vietnam.

#### Broader Societal Factors

Broader societal factors fall into two groups, with hours per day spent studying generally being associated with lower psychological distress, while role limitation and gender wage gap were generally associated with higher psychological distress.

Study amount was higher in India than in other countries. Study amounts were higher in girls than in boys in all countries but only significantly so in Peru and Vietnam. Higher study amount was generally associated with lower psychological distress, significantly so in both boys and girls in India and Vietnam, but only for girls in Peru, resulting in a significant interaction effect. A higher gender wage gap was associated with higher psychological distress in all countries and was significant for both boys and girls in India and Vietnam. Role limitation was higher in girls than in boys in all countries. It was mostly associated with higher psychological distress, significantly so in boys and girls in India and boys in Vietnam.

## Discussion

In this paper we first proposed a comprehensive conceptual framework of potential factors that might help to explain the gender gap in adolescent psychological distress. We then used longitudinal data from four countries: Ethiopia, India, Peru and Vietnam to examine a subset of available factors from this framework in explaining the observed gender gaps. We investigated whether these risk factors were gendered in their distribution or gendered in their impacts on distress. We also delineated country differences in the observed contributions of these factors to the gender gap in psychological distress. The gender gap in psychological distress across these four countries varied greatly from 0.62 in Ethiopia and Vietnam to 1.76 in India and 2.45 in Peru, with distress in girls being higher than in boys in all countries.

### The need for a comprehensive multi-domain framework

We found that factors across all the domains in the framework are relevant for adolescent mental health and the observed gender gap, highlighting the importance of considering multifactorial causes of this inequality. The results also highlight the importance of context-based factors (rather than just biological factors), demonstrating the potential for efforts to modify the exposure that girls have to gendered risk factors and to mitigate their impacts on girls’ mental health, helping to reduce these inequalities.

No single dataset will contain information on all potential explanations for the gender gap, and our study was unable to investigate several such factors across multiple domains. For example, data on potentially important factors such as stress biomarkers were not available as these types of data are harder to collect and often not included in large population-based studies. Importantly, many variables related to gendered experiences such as sexual violence and gender discrimination perceptions were also not included in these studies. This is not unusual, and we have found that many large-scale population-based studies do not collect these data despite the widespread nature of these experiences and their potential relevance. The lack of inclusion of factors that might be highly relevant, especially to women, highlights structural issues in data availability and what data holders choose to focus on and exclude.

It is also important to note that factors in the framework are not independent and many are causes of others, especially broader societal factors such as sexism and structural gender inequalities cause several of the individual level behavioural, psychological and biological factors to manifest in gendered ways. One example is self-efficacy, where structural sexism at a broader societal level has been associated with gender differences, whereby equally qualified women report lower levels of self-efficacy than comparable men (Homan, 2024). Another finding is that the gender gap in adolescent mental health is in some cases higher in more gender-equal countries (Campbell et al., 2021). The reasons for this are likely to involve complex interactions between expectations, norms, experiences, attitudes and behaviours. For instance, De Looze et al. (2025) suggested that this relationship is in part mediated by factors such as an increase in academic pressure and a decrease in social support, which have accompanied the increase in gender-equality in many countries over the past 20 years. They hypothesised that these factors have had a greater impact on girls, who have been under increasing pressure to achieve academic and financial success while also fulfilling more traditional gender-based roles, resulting in role overload.

#### Gendered risk factors and gendered impacts

Some factors were clearly gendered in their prevalence across all four countries included in this study. Forced marriage, caregiver gender preference, and role overload were all experienced more by girls than boys, whereas boys were exposed to greater violence and physical bullying victimisation compared to girls. Some risk factors, however, showed opposite patterns of prevalence across countries, with boys reporting better peer relations than girls in Ethiopia but worse peer relations than girls in Peru and Vietnam and girls achieving higher maths scores in Vietnam but lower scores in Ethiopia and India. Other examples include meal frequency (girls more in Ethiopia, boys more in India), social support (girls more in Peru and Vietnam and boys more in India) and self-efficacy (girls more in Peru and boys in India). These findings highlight the highly context specific nature of some of these experiences and norms, further stressing the importance of examining the drivers of mental health and the gender gap in varied global contexts.

Gendered impacts were seen for some factors in most countries, e.g. sleep amount was associated more strongly with distress in girls than boys and greater community trust with lower distress in boys compared to girls. However, for most factors where a significant gender difference in impact is observed, these are specific to one or two countries (e.g. meal frequency, social support, food insecurity, self-esteem etc.). For example, the meaning, experience and access to food might be more gendered in some contexts compared to others. Evidence from India shows that boys have greater dietary diversity and food advantage across childhood due to preferential treatment in the intrahousehold distribution of food (Aurino, 2017).

There are also factors for which the gendered impacts are in opposite directions in different countries. For instance, instability of location was associated more strongly with distress in boys in Ethiopia and Peru, but with girls’ distress in Vietnam. These findings highlight how understanding the social and cultural context of a risk factor is critical, and further research, including qualitative research, to understand how certain risk factors are experienced by girls and boys and how these experiences might vary based on context, social norms and beliefs would shed light on the mechanisms at play.

#### Cross-Country differences

Differences observed in the size of the gender gap in psychological distress in the four countries as well as country differences in the prevalence of explanatory factors and their impact on psychological distress highlight the importance of research on such questions being conducted in different countries. Very little evidence regarding the causes and development of inequalities in adolescent mental health comes from settings outside a handful of high-income White-majority countries. This is despite 90% of the world’s children and young people living in LMICs. Findings from one country context do not seem to translate to other country contexts and it is vital that more is done to understand the drivers of mental health inequalities in a wider range of global contexts.

An example of the importance of considering country context can be illustrated with the bullying victimisation factors in India and Peru. While physical bullying victimisation is more prevalent in India than in Peru, psychological bullying victimisation is more prevalent in Peru. Physical bullying victimisation is more common in boys than in girls in both countries, however the association with psychological distress is different. In India, the impact on psychological distress of both physical and psychological bullying victimisation is greater in girls than boys, while in Peru the impact is greater in boys than in girls. Other researchers have found different factors associated with bullying in the two countries. For example, Pells et al. (2016) found a significant association between economic status and bullying victimisation in India, but not Peru, while in Peru, but not India, the authors found that adolescent bullying victimisation was associated with later (at age 19) reduced self-esteem and worse parent and peer relations, which in turn have a negative impact on psychological distress. Rojas Arangoitia (2011) found evidence that a culture of physical punishment as a disciplinary tool at a high school in Peru resulted in physical violence also being justified in the eyes of students. These findings highlight not only the importance of considering differences in explanatory factors between and even within countries, but also that interactions between factors and their impact on psychological distress may differ across cultural contexts. The fact that even where a risk factor appears universal in its gendered prevalence, the mechanisms connecting it to psychological distress may be culturally specific has important implications for intervention design.

#### Gender norms

Many of the gendered patterns observed in the explanatory factors included in this study are, at least in part, downstream expressions of broader gender norms, where gender norms reflect the social rules determining which behaviours and attributes are valued and deemed appropriate in different genders (Heise et al., 2019). Viewed through this lens, the consistency with which certain factors, such as role overload, forced marriage, and caregiver gender preference, were more prevalent among girls in all four countries reflects traditional gender norms, which constrain girls’ autonomy and opportunities. Similarly, boys’ greater exposure to violence and physical bullying victimisation could be a consequence of norms associated with masculinity which prioritise toughness and physical dominance while encouraging emotional suppression.

These findings are consistent with those of a recent qualitative study conducted by Fine et al. (2025) which drew on focus group discussions with adolescents across 13 countries to explore gendered influences on mental health. They found evidence of gendered patterns in the mechanisms through which gender norms impact mental health. For girls, they observed an indirect impact, whereby gender norms expose them to increased scrutiny, closer oversight, pressures around appearance and elevated risk of gender-based violence, which in turn have an adverse effect on mental health. By contrast, for boys they observed a direct impact, whereby norms such as emotional suppression lead directly to poor mental health outcomes including depression and suicidal ideation. Also, in line with the findings of our study, Fine et al. found cross-cultural variability both in the nature of the gender norms as well as in their impact on mental health outcomes. Taken together, these findings highlight the need for mental health frameworks and policies which address harmful gender norms directly, in addition to addressing their downstream consequences.

The gender gap in depression and distress which emerges in adolescence is known to remain at similar levels during adulthood (Kuehner, 2017). While we would expect the broad domains of our framework to remain relevant across the life course, it is likely that some of the specific factors within these domains change with developmental stage. This has important policy implications. Some of the explanatory factors in our framework (e.g. access to education) are relatively amenable to direct policy intervention, whereas others, such as role overload, may reflect embedded social norms which are more resistant to change and may become even more entrenched in adulthood, requiring longer-term, gender-transformative approaches (Heise et al., 2019; Heymann et al., 2019).

#### Strengths and limitations

We have proposed a comprehensive multi-domain framework to explain the gender gap in adolescent mental health, synthesising findings from the large body of existing research on the topic and building upon existing frameworks. In constructing the framework, we also sought, where possible, to incorporate findings from research conducted outside of western, high-income contexts. Unlike prior papers with proposed frameworks, we go beyond solely proposing the framework and illustrate an application of the framework to data from four diverse countries. We examined a substantial number of the explanatory variables, testing for gender differences in their prevalence, as well as in their association with psychological distress. Finally, in a more exploratory piece of analysis, we used a machine-learning based model to evaluate the performance of the framework in predicting the gender gap in adolescent psychological distress, along with model inspection techniques to understand the contribution of different explanatory variables to the model predictions and performance.

Limitations of our study include the following. Firstly, due to a lack of available data, we were not able to capture all of the factors from our theoretical framework in the quantitative analysis, however, it is unlikely that one dataset with data on all the factors in the framework exists, and it will be necessary to use different data sources to capture the full range of potential causes of the gender gap. Secondly, we focus on binary gender based upon biological sex. There is compelling evidence that gender-minority adolescents are at increased risk of experiencing depression and anxiety (White et al., 2023) and research with more comprehensive measures of gender are also needed, however, due to small numbers of gender minorities, datasets like the ones used in this study would not be suitable and other larger datasets or those that intentionally oversample gender minorities will need to be analysed. Thirdly, since not all data were consistently available at all time-points in the study, we measured many factors at particular ages during adolescence. Fourthly, our measure of psychological distress, the Emotional Symptoms scale of the Strengths and Difficulties Questionnaire, albeit well-validated and widely used, is a brief measure of mild to moderate distress symptoms and does not capture severe symptoms of distress.

Finally, the focus of our study was breadth not depth, resulting in our inclusion of as many factors as possible from our framework in the quantitative analysis. A consequence of this was that we did not aim to provide in-depth investigations into each of the examined factors and instead focused on a breadth of potentially important considerations across several domains. This approach aligned with the multi-domain framework which proposed many different factors and is a useful attempt to understand the breadth of relevant factors driving observed gender inequalities in mental health. This approach also suited the multi-disciplinary cohort data which we used, which had information on many aspects of the children’s lives. Our analysis hopefully lays the foundations for more focused future research exploring some of the gender and country differences identified in greater depth. More in-depth studies focused on smaller subsets of factors using more detailed measures will help further understand their role in mental health development more generally and the gender gap in this. In addition to the gendered prevalence and gendered impact of factors which we considered in our analysis, there is evidence of gender differences in the nature of the experience of different factors. For example, Wilson et al. (2023) highlighted the potential importance of gender differences in the experience of bullying victimisation in contributing to gender differences in the association between bullying and self-injurious thoughts and behaviours. More in-depth studies will be required to understand such effects. It is also likely that some of the examined factors have more complex non-linear relationships with mental health (for example sleep and weight, where both high and low levels might be associated with greater distress), and in-depth studies would be able to investigate non-linear relationships in greater detail.

## Conclusion

The most important contribution of this paper is in developing a framework for the factors contributing to the gender gap in adolescent psychological distress and in providing evidence for the importance of specific factors in the context of the four LMICs included in the study. Just as mental health is a complex phenomenon with interacting bio-psycho-socio-environmental causes, the gender gap in mental ill-health is also complex with explanations across multiple domains. Our findings also demonstrate that the extent and drivers of the observed gender gap vary by country context, highlighting the importance of studying young people’s mental health development in diverse populations, avoiding generalising from one context to another and trying to further understand different contextual factors which drive these mental health inequalities.

## Declarations

### Ethical approval and consent to participate

Each round of data collection for Young Lives cohort has received the necessary ethical approvals and participants consented to take part in their respective studies. Details can be found here: https://www.younglives.org.uk/research-ethics

### Consent for publication

Not applicable

## Availability of data and materials

The data used in this study are available to download for research purposes from the UK Data Service (https://ukdataservice.ac.uk/). The analysis code can be found at github (https://github.com/stevebudapest/Adolescent-Psychological-Distress).

### Funding

SA is funded by a UCL-Wellcome Trust doctoral training fellowship in mental health science.

### Author contributions

[CRediT (Contributor Roles Taxonomy)](https://credit.niso.org/) roles:

- Conceptualization: PP
- Data curation: SA
- Formal analysis: SA
- Methodology: SA, PP
- Supervision: PP
- Visualization: SA
- Writing – original draft: SA, PP
- Writing – review and editing: All authors

## Acknowledgements

The data used in this publication come from Young Lives, a longitudinal study of childhood poverty and transitions to adulthood in Ethiopia, India, Peru and Vietnam (www.younglives.org.uk). Young Lives is funded by UK aid from the Foreign, Commonwealth & Development Office and a number of further funders. The views expressed here are those of the authors. They are not necessarily those of Young Lives, the University of Oxford, FCDO or other funders.

## Reporting

We follow the STROBE reporting guidelines for observational studies – cohort studies (https://www.strobe-statement.org/checklists). We report how we determined our sample size, all data exclusions, all manipulations, and all measures in the study.

## References

1. Adedeji, A., Otto, C., Kaman, A., Reiss, F., Devine, J., & Ravens-Sieberer, U. (2021). Peer Relationships and Depressive Symptoms Among Adolescents: Results From the German BELLA Study. Frontiers in Psychology, 12, 767922. 10.3389/fpsyg.2021.767922

2. Aurino, E. (2017). Do boys eat better than girls in India? Longitudinal evidence on dietary diversity and food consumption disparities among children and adolescents. Economics & Human Biology, 25, 99–111. 10.1016/j.ehb.2016.10.007

3. Bandura, A. (1977). Self-efficacy: Toward a unifying theory of behavioral change. Psychological Review, 84(2), 191–215. 10.1037/0033-295X.84.2.191

4. Bentivegna, F., & Patalay, P. (2022). The impact of sexual violence in mid-adolescence on mental health: A UK population-based longitudinal study. The Lancet. Psychiatry, 9(11), 874–883. 10.1016/S2215-0366(22)00271-1

5. Branje, S. J. T., Hale, W. W., Frijns, T., & Meeus, W. H. J. (2010). Longitudinal associations between perceived parent-child relationship quality and depressive symptoms in adolescence. Journal of Abnormal Child Psychology, 38(6), 751–763. 10.1007/s10802-010-9401-6

6. Breiman, L. (2001). Random Forests. Machine Learning, 45(1), 5–32. 10.1023/A:1010933404324

7. Campbell, O. L. K., Bann, D., & Patalay, P. (2021). The gender gap in adolescent mental health: A cross-national investigation of 566,829 adolescents across 73 countries. SSM – Population Health, 13, 100742. 10.1016/j.ssmph.2021.100742

8. Chen, T., & Guestrin, C. (2016, March 9). XGBoost: A Scalable Tree Boosting System. arXiv.Org. 10.1145/2939672.2939785

9. Das, M., Angeli, F., Krumeich, A. J. S. M., & Van Schayck, O. C. P. (2018). The gendered experience with respect to health-seeking behaviour in an urban slum of Kolkata, India. International Journal for Equity in Health, 17(1), 24. 10.1186/s12939-018-0738-8

10. De Looze, M. E., Cosma, A., Elgar, F. J., Schrijvers, K., Inchley, J., Walsh, S. D., & Stevens, G. W. J. M. (2025). Exploring mechanisms behind the increasing gender gap in adolescent psychological symptoms, 2002–2022: The role of national-level gender equality. Journal of Child Psychology and Psychiatry, 66(5), 737–751. 10.1111/jcpp.14081

11. Demkowicz, O., Jefferson, R., Nanda, P., Foulkes, L., Lam, J., Pryjmachuk, S., Evans, R., Dubicka, B., Neill, L., Winter, L. A., & Nnamani, G. (2025). Adolescent girls’ explanations of high rates of low mood and anxiety in their population: A co-produced qualitative study. BMC Women’s Health, 25(1), 49. 10.1186/s12905-024-03517-x

12. Elmore, A. L., & Crouch, E. (2020). The Association of Adverse Childhood Experiences With Anxiety and Depression for Children and Youth, 8 to 17 Years of Age. Academic Pediatrics, 20(5), 600–608. 10.1016/j.acap.2020.02.012

13. Else-Quest, N. M., Hyde, J. S., & Linn, M. C. (2010). Cross-national patterns of gender differences in mathematics: A meta-analysis. Psychological Bulletin, 136(1), 103–127. 10.1037/a0018053

14. Espinosa, F., Martin-Romero, N., & Sanchez-Lopez, A. (2022). Repetitive Negative Thinking Processes Account for Gender Differences in Depression and Anxiety During Adolescence. International Journal of Cognitive Therapy, 15(2), 115–133. 10.1007/s41811-022-00133-1

15. Fine, S. L., Harrison, A., Rykiel, N. A., & Herrera, M. M. (2025). Adolescents’ Perceptions of Gendered Influences on Mental Health: Results From a 13-Country Qualitative Study. Journal of Adolescent Health, 77(3), 413–420. 10.1016/j.jadohealth.2024.02.036

16. Fredrickson, B. L., & Roberts, T.-A. (1997). Objectification Theory: Toward Understanding Women’s Lived Experiences and Mental Health Risks. Psychology of Women Quarterly, 21(2), 173–206. 10.1111/j.1471-6402.1997.tb00108.x

17. Gajos, J. M., Miller, C. R., Leban, L., & Cropsey, K. L. (2022). Adverse childhood experiences and adolescent mental health: Understanding the roles of gender and teenage risk and protective factors. Journal of Affective Disorders, 314, 303–308. 10.1016/j.jad.2022.07.047

18. Goodman, R. (1997). The Strengths and Difficulties Questionnaire: A research note. Journal of Child Psychology and Psychiatry, and Allied Disciplines, 38(5), 581–586. 10.1111/j.1469-7610.1997.tb01545.x

19. Heise, L., Greene, M. E., Opper, N., Stavropoulou, M., Harper, C., Nascimento, M., Zewdie, D., Darmstadt, G. L., Greene, M. E., Hawkes, S., Heise, L., Henry, S., Heymann, J., Klugman, J., Levine, R., Raj, A., & Rao Gupta, G. (2019). Gender inequality and restrictive gender norms: Framing the challenges to health. The Lancet, 393(10189), 2440–2454. 10.1016/S0140-6736(19)30652-X

20. Heymann, J., Levy, J. K., Bose, B., Ríos-Salas, V., Mekonen, Y., Swaminathan, H., Omidakhsh, N., Gadoth, A., Huh, K., Greene, M. E., Darmstadt, G. L., Darmstadt, G. L., Greene, M. E., Hawkes, S., Heise, L., Henry, S., Heymann, J., Klugman, J., Levine, R., … Rao Gupta, G. (2019). Improving health with programmatic, legal, and policy approaches to reduce gender inequality and change restrictive gender norms. The Lancet, 393(10190), 2522–2534. 10.1016/S0140-6736(19)30656-7

21. Homan, P. (2024). Health consequences of structural sexism: Conceptual foundations, empirical evidence and priorities for future research. Social Science & Medicine, 351, 116379. 10.1016/j.socscimed.2023.116379

22. Hossain, M., & Jukes, M. C. H. (2023). Gender differences in socioemotional skills among adolescents and young adults in Ethiopia, India, Peru and Vietnam. Young Lives. https://ora.ox.ac.uk/objects/uuid:961849d4-9f3d-4c06-8049-79a8dd37ddd3

23. Hyde, J. S., & Mezulis, A. H. (2020). Gender Differences in Depression: Biological, Affective, Cognitive, and Sociocultural Factors. Harvard Review of Psychiatry, 28(1), 4–13. 10.1097/HRP.0000000000000230

24. Hyde, J. S., Mezulis, A. H., & Abramson, L. Y. (2008). The ABCs of depression: Integrating affective, biological, and cognitive models to explain the emergence of the gender difference in depression. Psychological Review, 115(2), 291–313. 10.1037/0033-295X.115.2.291

25. Jeong, J.-E., Joo, S.-H., Hahn, C., Kim, D.-J., & Kim, T.-S. (2019). Gender-Specific Association between Alcohol Consumption and Stress Perception, Depressed Mood, and Suicidal Ideation: The 2010–2015 KNHANES. Psychiatry Investigation, 16(5), 386–396. 10.30773/pi.2019.02.28

26. Johansen, R., Espetvedt, M. N., Lyshol, H., Clench-Aas, J., & Myklestad, I. (2021). Mental distress among young adults – gender differences in the role of social support. BMC Public Health, 21(1), 2152. 10.1186/s12889-021-12109-5

27. Kendler, K. S., Myers, J., & Prescott, C. A. (2005). Sex differences in the relationship between social support and risk for major depression: A longitudinal study of opposite-sex twin pairs. The American Journal of Psychiatry, 162(2), 250–256. 10.1176/appi.ajp.162.2.250

28. Keyes, K. M., & Platt, J. M. (2024). Annual Research Review: Sex, gender, and internalizing conditions among adolescents in the 21st century – trends, causes, consequences. Journal of Child Psychology and Psychiatry, and Allied Disciplines, 65(4), 384–407. 10.1111/jcpp.13864

29. Kuehner, C. (2017). Why is depression more common among women than among men? The Lancet Psychiatry, 4(2), 146–158. 10.1016/S2215-0366(16)30263-2

30. Li, S., Zhang, X., Cai, Y., Zheng, L., Pang, H., & Lou, L. (2023). Sex difference in incidence of major depressive disorder: An analysis from the Global Burden of Disease Study 2019. Annals of General Psychiatry, 22, 53. 10.1186/s12991-023-00486-7

31. Lundberg, S. M., Erion, G., Chen, H., DeGrave, A., Prutkin, J. M., Nair, B., Katz, R., Himmelfarb, J., Bansal, N., & Lee, S.-I. (2020). From local explanations to global understanding with explainable AI for trees. Nature Machine Intelligence, 2(1), 56–67. 10.1038/s42256-019-0138-9

32. Man, X., Liu, J., & Xue, Z. (2022). Effects of Bullying Forms on Adolescent Mental Health and Protective Factors: A Global Cross-Regional Research Based on 65 Countries. International Journal of Environmental Research and Public Health, 19(4), 2374. 10.3390/ijerph19042374

33. Marcotte, D., Fortin, L., Potvin, P., & Papillon, M. (2002). Gender differences in depressive symptoms during adolescence: Role of gender-typed characteristics, self-esteem, body image, stressful life events, and pubertal status. Journal of Emotional and Behavioral Disorders, 10(1), 29–42. 10.1177/106342660201000104

34. McPherson, K. E., Kerr, S., McGee, E., Morgan, A., Cheater, F. M., McLean, J., & Egan, J. (2014). The association between social capital and mental health and behavioural problems in children and adolescents: An integrative systematic review. BMC Psychology, 2(1), 7. 10.1186/2050-7283-2-7

35. Morken, I. S., Viddal, K. R., von Soest, T., & Wichstrøm, L. (2023). Explaining the Female Preponderance in Adolescent Depression-A Four-Wave Cohort Study. Research on Child and Adolescent Psychopathology, 51(6), 859–869. 10.1007/s10802-023-01031-6

36. Muris, P. (2002). Relationships between self-efficacy and symptoms of anxiety disorders and depression in a normal adolescent sample. Personality and Individual Differences, 32(2), 337–348. 10.1016/S0191-8869(01)00027-7

37. Newbury, J. B., Heron, J., Kirkbride, J. B., Fisher, H. L., Bakolis, I., Boyd, A., Thomas, R., & Zammit, S. (2024). Air and Noise Pollution Exposure in Early Life and Mental Health From Adolescence to Young Adulthood. JAMA Network Open, 7(5), e2412169. 10.1001/jamanetworkopen.2024.12169

38. Niemeyer, H., Bieda, A., Michalak, J., Schneider, S., & Margraf, J. (2019). Education and mental health: Do psychosocial resources matter? SSM – Population Health, 7, 100392. 10.1016/j.ssmph.2019.100392

39. Nolen-Hoeksema, S. (2001). Gender Differences in Depression. Current Directions in Psychological Science, 10(5), 173–176. 10.1111/1467-8721.00142

40. Patalay, P., & Demkowicz, O. (2023). Debate: Don’t mind the gap – why do we not care about the gender gap in common mental health difficulties? Child and Adolescent Mental Health, 28(2), 341–343. 10.1111/camh.12647

41. Patalay, P., & Fitzsimons, E. (2021). Psychological distress, self-harm and attempted suicide in UK 17-year olds: Prevalence and sociodemographic inequalities. The British Journal of Psychiatry, 219(2), 437–439. 10.1192/bjp.2020.258

42. Pells, K., Ogando Portela, M. J., & Espinoza Revollo, P. (2016). Experiences of Peer Bullying among Adolescents and Associated Effects on Young Adult Outcomes: Longitudinal Evidence from Ethiopia, India, Peru and Viet Nam (Innocenti Discussion Papers No. 2016/03; Innocenti Discussion Papers, Vol. 2016/03). 10.18356/c2e90aee-en

43. Piccinelli, M., & Wilkinson, G. (2000). Gender differences in depression. Critical review. The British Journal of Psychiatry: The Journal of Mental Science, 177, 486–492. 10.1192/bjp.177.6.486

44. Porter, C., Favara, M., Hittmeyer, A., Scott, D., Jiménez, A. S., Ellanki, R., Woldehanna, T., Duc, L. T., Craske, M. G., & Stein, A. (2021). Impact of the COVID-19 pandemic on anxiety and depression symptoms of young people in the global south: Evidence from a four-country cohort study. BMJ Open, 11(4), e049653. 10.1136/bmjopen-2021-049653

45. Reiss, F. (2013). Socioeconomic inequalities and mental health problems in children and adolescents: A systematic review. Social Science & Medicine (1982), 90, 24–31. 10.1016/j.socscimed.2013.04.026

46. Rojas Arangoitia, V. (2011). ‘I’d rather be hit with a stick—Grades are sacred’: Students’ perceptions of discipline and authority in a public high school in Peru. Young Lives.

47. Rosenberg, M. (1965). The measurement of self-esteem. In M. Rosenberg, Society and the Adolescent Self-Image (pp. 16–36). Princeton University Press. 10.1515/9781400876136-003

48. Salk, R. H., Hyde, J. S., & Abramson, L. Y. (2017). Gender Differences in Depression in Representative National Samples: Meta-Analyses of Diagnoses and Symptoms. Psychological Bulletin, 143(8), 783–822. 10.1037/bul0000102

49. Schwarzer, R., & Jerusalem, M. (1995). Optimistic self-beliefs as a resource factor in coping with stress. In Extreme stress and communities: Impact and intervention (pp. 159–177). Kluwer Academic/Plenum Publishers. 10.1007/978-94-015-8486-9_7

50. Singh, R., Kesarwani, R., & Mukherjee, P. (2019). Exploring well-being among 22-year-old youth in India (Vol. 187). Young Lives. https://ora.ox.ac.uk/objects/uuid:ad0069db-f970-4dc6-a2fe-c9afe55d0858

51. Solmi, F., Sharpe, H., Gage, S. H., Maddock, J., Lewis, G., & Patalay, P. (2021). Changes in the Prevalence and Correlates of Weight-Control Behaviors and Weight Perception in Adolescents in the UK, 1986-2015. JAMA Pediatrics, 175(3), 267. 10.1001/jamapediatrics.2020.4746

52. Stentiford, L., Koutsouris, G., Nash, T., & Allan, A. (2025). Mental health and gender discourses in school: “Emotional” girls and boys “at risk”. Educational Review, 77(5), 1364–1382. 10.1080/00131911.2024.2306947

53. Sun, Y., Zhong, Y., Sun, W., Chu, L., Long, J., & Fan, X. W. (2023). More prevalent and more severe: Gender differences of depressive symptoms in Chinese adolescents. Frontiers in Public Health, 11, 1167234. 10.3389/fpubh.2023.1167234

54. Thompson, A. E., Anisimowicz, Y., Miedema, B., Hogg, W., Wodchis, W. P., & Aubrey-Bassler, K. (2016). The influence of gender and other patient characteristics on health care-seeking behaviour: A QUALICOPC study. BMC Family Practice, 17(1), 38. 10.1186/s12875-016-0440-0

55. Tiggemann, M., & Slater, A. (2015). The Role of Self-Objectification in the Mental Health of Early Adolescent Girls: Predictors and Consequences. Journal of Pediatric Psychology, 40(7), 704–711. 10.1093/jpepsy/jsv021

56. Tone, E. B., & Tully, E. C. (2014). Empathy as a “Risky Strength”: A Multilevel Examination of Empathy and Risk for Internalizing Disorders. Development and Psychopathology, 26(4 0 2), 1547–1565. 10.1017/S0954579414001199

57. Treynor, W., Gonzalez, R., & Nolen-Hoeksema, S. (2003). Rumination Reconsidered: A Psychometric Analysis. Cognitive Therapy and Research, 27(3), 247–259. 10.1023/A:1023910315561

58. UNDP (2023). In Human Development Reports. United Nations. Retrieved 7 September 2025, from https://hdr.undp.org/data-center/thematic-composite-indices/gender-inequality-index

59. Van de Velde, S., Huijts, T., Bracke, P., & Bambra, C. (2013). Macro-level gender equality and depression in men and women in Europe. Sociology of Health & Illness, 35(5), 682–698. 10.1111/j.1467-9566.2012.01521.x

60. White, J., Moore, L., Cannings-John, R., Hawkins, J., Bonell, C., Hickman, M., Zammit, S., & Adara, L. (2023). Association Between Gender Minority Status and Mental Health in High School Students. Journal of Adolescent Health, 72(5), 811–814. 10.1016/j.jadohealth.2022.12.028

61. Wilkinson, A. L., Halpern, C. T., & Herring, A. H. (2016). Directions of the relationship between substance use and depressive symptoms from adolescence to young adulthood. Addictive Behaviors, 60, 64–70. 10.1016/j.addbeh.2016.03.036

62. Wilson, E., Crudgington, H., Morgan, C., Hirsch, C., Prina, M., & Gayer-Anderson, C. (2023). The longitudinal course of childhood bullying victimization and associations with self-injurious thoughts and behaviors in children and young people: A systematic review of the literature. Journal of Adolescence, 95(1), 5–33. 10.1002/jad.12097

63. World Bank Group (2024). World Bank Open Data. Retrieved 7 September 2025, from https://data.worldbank.org

64. World Health Organization (2025). Depression. Retrieved 24 April 2025, from https://www.who.int/health-topics/depression

65. Yong, M., McCarty, C. A., Vander Stoep, A., & McCauley, E. A. (2022). Gender differences in the effects of academic achievement on depressive symptoms during adolescence. Journal of Child and Family Studies, 31(12), 3326–3341. 10.1007/s10826-022-02414-x

66. Zaneva, M., Guzman-Holst, C., Reeves, A., & Bowes, L. (2022). The Impact of Monetary Poverty Alleviation Programs on Children’s and Adolescents’ Mental Health: A Systematic Review and Meta-Analysis Across Low-, Middle-, and High-Income Countries. Journal of Adolescent Health, 71(2), 147–156. 10.1016/j.jadohealth.2022.02.011

