## Supplementary Data for "Why are girls more distressed than boys? A multi-domain framework applied to longitudinal data across four countries"

Affiliations:

**Supplementary Data 1: Details of Fields Used for the Primary Outcome Variable**

The primary outcome measure was the Emotional Symptoms Scale dimension of the Strengths and Difficulties Questionnaire (SDQ), calculated based upon participant responses to the following 5 questions:

I get a lot of headaches, stomach aches or sickness

I worry a lot

I am often unhappy, downhearted or tearful

I am nervous in new situations

I have many fears, and am easily scared

Participants in Ethiopia, India and Vietnam stated whether they strongly disagreed, disagreed, agreed or strongly agreed with each of the above statements to which scores of 0, 1, 2 and 3 were applied respectively. For Peru the response options were "Verdadero", "Mas o menos", "Falso" to which scores of 3, 1.5 and 0 were applied respectively. This ensured that the total score for each country was between 0 and 15.

**Supplementary Figure S1: The proposed framework displaying potential explanatory factors for which data was available in the Young Lives study**

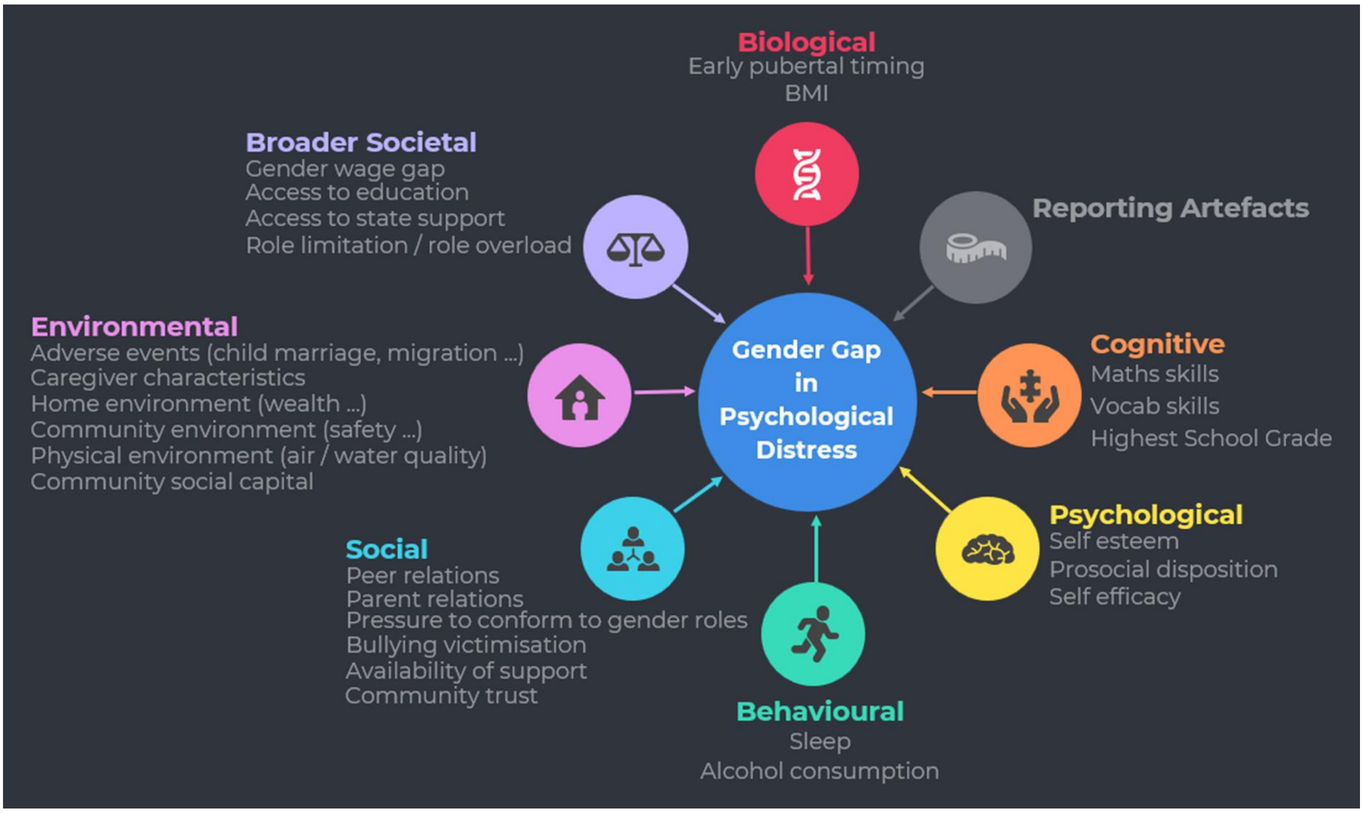

**Supplementary Table S1: Explanatory factors from our framework available in the Young Lives data and used in our quantitative analysis along with rationale for inclusion**

| Biological Domain | |
|---|---|
| **Factor** | **Why is the Factor Included?** |
| Puberty / Early pubertal timing | Research suggests that early pubertal timing is associated with increased risk of psychological distress in girls but not in boys (Kuehner, 2017). |
| Weight | Weight and subsequent weight related feelings and behaviours are differentially associated with distress symptoms in girls and boys (Solmi et al. 2021). |

| Cognitive Domain | |
|---|---|
| **Factor** | **Why is the Factor Included?** |
| Maths / Vocab Skills | There is evidence that academic performance can be predictive of experiencing depressive symptoms in adolescents. Yong et al. (2022) observed a gender difference in this association with poor GPA scores predicting depressive symptoms only in girls. |
| Highest Education Grade | There is a well-documented association between education level and experience of mental health issues (Niemeyer et al., 2019). However, there is evidence of a complex relationship between education level and the gender gap in psychological distress (Keyes and Platt, 2024). |

| Psychological Domain | |
|---|---|
| **Factor** | **Why is the Factor Included?** |
| Prosocial disposition | There is evidence of gender differences in childhood and adolescence in prosociality (Kuehner, 2017). Some researchers have proposed that prosociality can interact with other personality characteristics to confer vulnerability to psychological distress (Tone and Tully, 2014). |
| Self-efficacy | Self-efficacy is an individual's evaluation of their own ability to achieve a particular goal (Bandura, 1977). Research suggests that a gender gap in self-efficacy emerges in late adolescence with boys having higher self-efficacy than girls (Hossain and Jukes, 2023), while low self-efficacy has been associated with development of psychological distress (Muris, 2002). |
| Self-esteem | Research suggests that a gender gap in self-esteem emerges in late adolescence (in certain countries) with boys having higher self-esteem than girls (Hossain and Jukes, 2023), while low self-esteem has been associated with development of depression (Marcotte et al., 2002). |

| Behavioural Domain | |
|---|---|
| **Factor** | **Why is the Factor Included?** |

| | |
|---|---|
| Sleep | Research suggests that starting in adolescence, disrupted sleep is more common in girls than in boys, while sleep disorders are more predictive of depression in adolescent girls than in boys (Sun et al., 2023). |
| Alcohol consumption | There is evidence of a gender-specific association between alcohol consumption and depression (Jeong et al., 2019) with the association being stronger in adolescent girls than in boys. |

| Social Domain | |
|---|---|
| **Factor** | **Why is the Factor Included?** |
| Peer relations | There is evidence identifying different facets of peer relationships as predictors of depressive symptoms in adolescence (Adedeji et al., 2021). |
| Parent relations | There is research indicating that a strong relationship between adolescents and their parents can act as a protective factor against development of psychological distress, although there is evidence that the association can vary by gender and cultural context (Branje et al., 2010; Porter et al., 2021). |
| Bullying victimisation | Research suggests that there are gender differences in the association between different forms of bullying and the development of psychological distress (Man et al., 2022). |
| Availability of support | There is evidence of gender differences in the association between the availability of social support and depression (Kendler et al., 2005), with social support having been found to be a stronger protective factor for young women than for men (Johansen et al., 2021). |

| Environmental Domain | |
|---|---|
| **Factor** | **Why is the Factor Included?** |
| Adverse events | There is a well-documented association between exposure to adverse childhood experiences (ACEs) and psychological distress in later adolescence (Elmore and Crouch, 2020). The impact of ACEs on the gender gap in psychological distress is however complex and less well understood (Keyes and Platt, 2024). |
| Caregiver / parent characteristics | Caregiver characteristics including their wellbeing and education levels have been associated with the wellbeing of the children they care for (Singh et al., 2019). |
| Home environment | Low socioeconomic status has been associated with mental health problems in children and adolescents (Reiss, 2013). |
| Community environment | Community environment has been associated with wellbeing of children growing up in that community (Singh et al., 2019). |
| Physical environment | There is evidence linking exposure to air and noise pollution in childhood to anxiety and depression in later adolescence (Newbury et al., 2024). |

| | |
|---|---|
| Community social capital | Community social capital, defined as the social networks existing between and within households and communities which enhance economic and social conditions, have been associated with mental health outcomes in children and adolescents (McPherson et al., 2014). |

| Broader Societal Domain | |
|---|---|
| **Factor** | **Why is the Factor Included?** |
| Gender equality | Research suggests a complex relationship between gender equality and the gender gap in psychological distress, with some research finding that countries with greater gender equality have a larger mental health gender gap (Campbell et al., 2021). |
| Access to education | There is a well-documented association between education level and experience of mental health issues (Niemeyer et al., 2019). However, there is evidence of a more complex relationship between education level and the gender gap in psychological distress (Keyes and Platt, 2024). |
| Access to state support | While research suggests that state support, for example through poverty alleviation initiatives, can be effective in reducing adolescent psychological distress, there is evidence that conditions attached to such support can be detrimental to mental health in girls (Zaneva et al., 2022). |
| Role limitation / overload | Role limitation, role overload and competing social roles are associated with increased risk of depression in women (Nolen-Hoeksema, 2001). |

**Supplementary Table S2: Detailed description of the fields used from the Young Lives data in our quantitative analysis**

| Domain | Factor | Risk Factor | Field | Description | Measured at Age | Variable Type |
|---|---|---|---|---|---|---|
| Biological | Puberty / Early pubertal timing | Early pubertal timing | menarche; voice; hairchin | Did the child go through puberty before age 10 for girls or 11 for boys (one year below average). | 11 | Binary |
| Biological | Weight | Weight | bmi | BMI | 14 | Continuous |
| Cognitive | Maths / Vocab skills | Maths score | math | IRT standardised maths score | 14 | Continuous |
| Cognitive | Maths / Vocab skills | Vocab score | ppvt_score | IRT score - centered | 14 | Continuous |
| Cognitive | Education | Level of education | hghgrade | Highest grade achieved in school. Multi-point scale. | 18 | Categorical ordered |

| | | | | | | |
|---|---|---|---|---|---|---|
| Psychological | Prosocial disposition | Prosocial disposition | HRDTLKR3; HLPCHLR3; CSRVCMR3; AFFSCHR3 | Based upon 4 questions answered by participants themselves using a 3 point scale. We create a prosociality index from available data. | 14 | Categorical ordered |
| Psychological | Self-efficacy | Self-efficacy | CTRYHDR3; CFTRWRR3;CBRJOBR3; CPLDECR3; CNOCHCR3; CDVJOBR3 | Based upon 6 questions answered by participants themselves using a 5 point scale. We create a self-efficacy index from available data. | 14 | Categorical ordered |
| Psychological | Self-esteem | Self-esteem | CASHCLR3;CASHWKR3; CEMBBKR3; CASHSHR3; CWRUNIR3; CCLTRGR3 | Based upon 6 questions answered by participants themselves using a 5 point scale. We create a self-esteem index from available data. | 14 | Categorical ordered |
| Behavioural | Sleep | Sleep quantity | sleepr3 | Hours per day spent sleeping | 14 | Continuous |
| Behavioural | Alcohol consumption | Alcohol | chalcohol | Alcohol consumption frequency (6 point scale - higher number is more frequent alcohol) | 14 | Categorical ordered |
| Social | Peer relations | Self-reported relationships with peers and community members | CFRNSTR3; CTRTRSR3; CTESMER3; CBLGSCR3;CSHPRSR3; CTRTWSR3 | Based upon 6 questions answered by participants themselves using a 5 point scale. We create a peer-relations index from available data. | 14 | Categorical ordered |

| | | | | | | |
|---|---|---|---|---|---|---|
| Social | Parent relations | Self-reported relationships with parents | SPVIEWR3; TRFAIRR3 | Based upon 2 questions answered by participants themselves using a 4 point scale. We create a parent-relations index from available data. | 14 | Categorical ordered |
| Social | Bullying victimisation - physical | Self-reported physical bullying by other kids | PNCHDR3; HRTPHYR3 | Based upon 2 questions answered by participants themselves using a 5 point scale. We create a physical bullying index from available data. | 14 | Categorical ordered |
| Social | Bullying victimisation - verbal / psychological | Self-reported verbal bullying by other kids | STARNGR3; RFSTLKR3; CLDNMER3; TRBFRNR3; MDEFUNR3 | Based upon 5 questions answered by participants themselves using a 5 point scale. We create a verbal/psychological bullying index from available data. | 14 | Categorical ordered |
| Social | Availability of support | Support for individual | CHPSTYR3; CHWORKR3; CHWRRDR3; CHBLLYR3; CHRLGNR3; CHMNEYR3 | Based upon 6 questions answered by participants themselves with a binary response. We create a social-support index from available data. | 14 | Binary count |
| Social | Trust in the community | Trust in the community | CTRUSTR3; CGOVRGR3; CSFEOWR3 | Based upon 3 questions answered by participants themselves using a 5 point scale. We create a community trust index from available data. | 14 | Categorical ordered |
| Environmental | Adverse Events | Domestic Abuse | BFAMLY; BBYFRN | Ever been beaten up by family member or partner. | by 14 | Binary |
| Environmental | Adverse Events | Exposure to Violence | BTCHR; BSTRNG; BFRND; BEMPLY; | Ever been beaten up by teacher, stranger, friend, other young person. | by 14 | Binary |

| | | | | | | |
|---|---|---|---|---|---|---|
| | | | PNCHD; HRTPHY | | | |
| Environmental | Adverse Events | Forced Marriage / Maternity | marrcohab_age; birth_age | Married at age below 18; given birth to child at age below 18. | by 18 | Binary count |
| Environmental | Adverse Events | Exposure to Household Adverse Events | shregul; shcrime; shecon; shhouse; shfam; shother; shenv | Whether household has been victim of crime, change in regulation, damage, economic circumstances, environmental disaster etc. Sum of adverse events since previous round | by 18 | Binary count |
| Environmental | Adverse Events | Adverse Family Events | dadyrdied; momyrdied | Has participant experienced death of mother, father or both. | by 18 | Binary count |
| Environmental | Adverse Events | Instability of Living Location | moveidr4 | How many changes of living location in the past 4 years. | 14 to 18 | Count |
| Environmental | Caregiver / parent characteristics | Caregiver education | caredu | Caregiver highest level achieved | by 14 | Categorical ordered |
| Environmental | Caregiver / parent characteristics | Caregiver gender attitudes | numson; numdtr; famson; famdtr | Caregiver preference for sons or daughters; caregiver commitment to sons and daughters education. Binary yes/no for evidence of gender preference. | 11 | Binary count |
| Environmental | Home environment | Household resources | wi; hq; sv; cd | Average of indices for wealth, access to services, housing quality and consumer durables. | 14 | Continuous |
| Environmental | Home environment | Food Insecurity | food; foodsec; foodshrt | Has the household experienced food shortages | by 14 | Binary count |
| | Home environment | Meal frequency | FQTOTLR3 | How many times child ate food in past 24 hours. | 14 | Count |

| | | | | | | |
|---|---|---|---|---|---|---|
| Environmental | Community environment | Community safety | VIOLCRM; VLNTCRME; PROSCRM; PRSTITN; YUTHCRM; GANGS; ADLTBNDS; DRUGSLLG; DRUGADCT | Is the community affected by violent crime, prostitution, gangs, drugs. Index based upon the number of threats to community safety | by 14 | Binary count |
| Environmental | Community environment | Education | SV1; R3CSV1 | Child's perception of quality of primary school. 4 point scale from strongly agree to strongly disagree (high score better). | 11-14 | Categorical ordered |
| Environmental | Physical environment | Pollution | water; air; rubbish | Child's perception of quality of air, water and amount of rubbish. 3 point scale | 8 | Categorical ordered |
| Environmental | Community social capital | Community strength | TALKPRR3; ACTPRBR3; PARTAWR3; PROTSTR3; R3CSV5; R3CSV4; R3CTR1; R3CTR4; R3CTR5; R3CTR6 | Caregiver perspectives on sense of trust and safety in community and extent to which community acts together to address common interests. | 14 | Categorical ordered |
| Environmental | Community social capital | Support for household | FINHLPR3 | Number of sources of financial support for the household. 7 point scale. | 14 | Categorical ordered |
| Broader Societal | Gender equality | Gender pay disparity | lnd; plnt; weed; hrvs; post; pstr; othr; lndr; help; cnst; fact; taxi; secr; maid; shop; | Constructed metric for gender pay gap for different types of role. Continuous scale. | 14 | Continuous |

| | | | | | | |
|---|---|---|---|---|---|---|
| | | | tchr; polc; mech; cook; fish; tail; mltr; comp; oth. All with suffix male or fmle | | | |
| Broader Societal | Education | Amount of study | hschool; hstudy; | If child is still enrolled in school, average number of hours per day spent studying. | 11-18 | Continuous |
| Broader Societal | Access to state support | Access to state support | SITUIDR4(7)/LADCURR4(ET); SITUIDR4(9/10)/LADCURR4(IN/VN); SITUIDR4(8) / LADCURR4(PE) | Support from Government/NGOs/ Local Associations. 9 point scale. | 18 | Categorical ordered |
| Broader Societal | Role limitation / overload | Role overload | enrol; hcare; hchore; htask; hwork | If child is still enrolled in school, average number of hours per day spent carrying out domestic chores and working. | 11-18 | Continuous |
| Broader Societal | Role limitation / overload | Role limitation | hcare; hchore; htask | Hours per day spent on caring and domestic tasks | 14 | Continuous |

**Supplementary Table S3: Psychological Distress differs significantly by gender in each of the four countries**

| Statistics | Ethiopia | India | Peru | Vietnam |
|---|---|---|---|---|
| Mean girls | 7.20 | 7.39 | 7.99 | 7.43 |
| Mean boys | 6.58 | 5.63 | 5.54 | 6.81 |
| Mean Gap | 0.62 | 1.76 | 2.45 | 0.62 |
| t-value | 3.63 | 10.55 | 11.44 | 3.53 |
| p | 0.008 | < 0.001 | < 0.001 | 0.012 |
| d | 0.24 | 0.69 | 0.95 | 0.24 |

**Supplementary Figure S2: Distribution of Explanatory Factors by Gender and Country**

***Biological Factors***

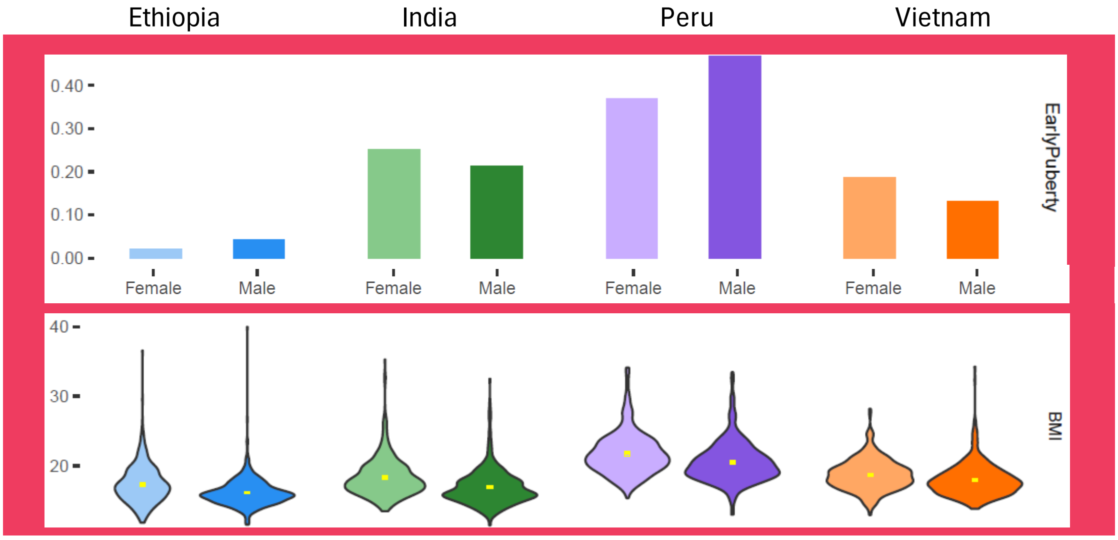


***Cognitive Factors***

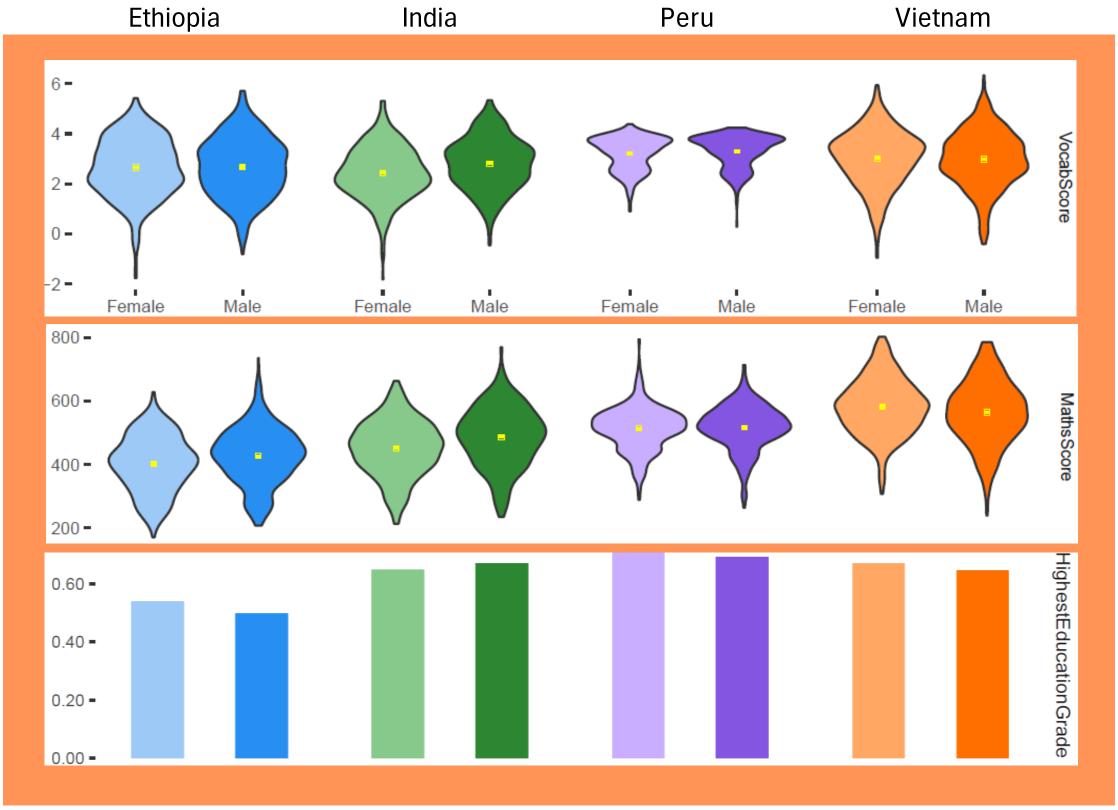

### ***Psychological Factors***

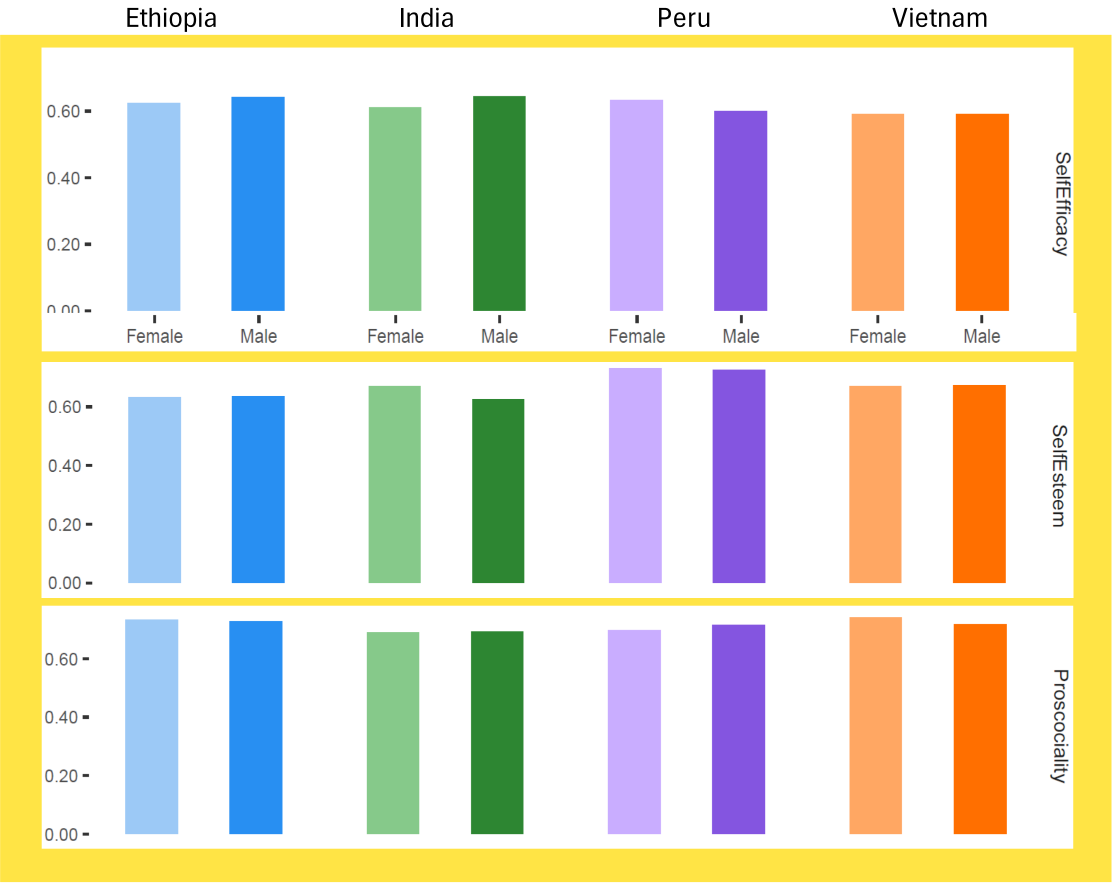


### ***Behavioural Factors***

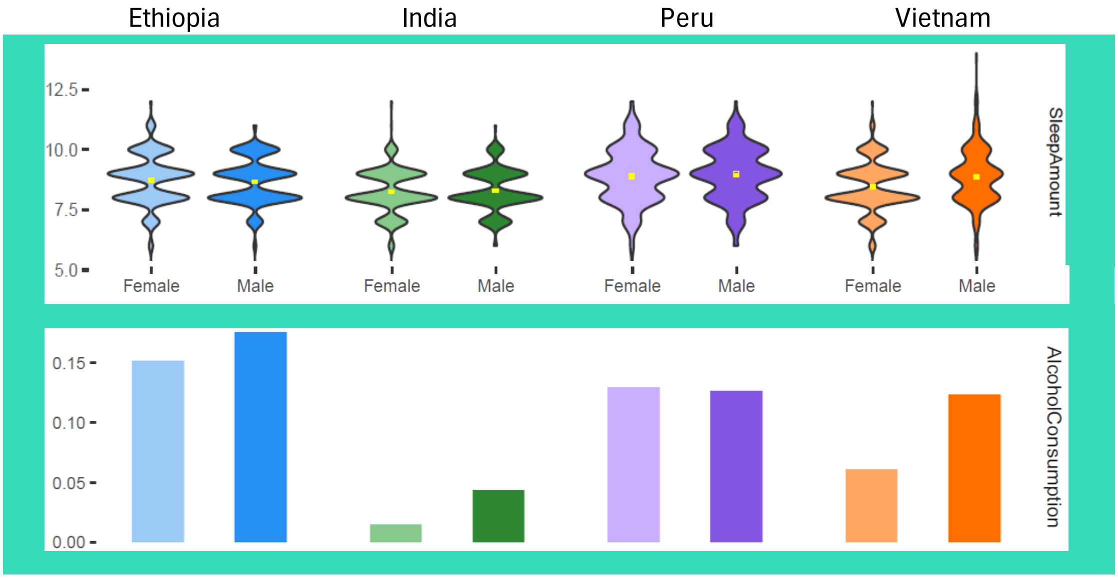

***Social Factors***

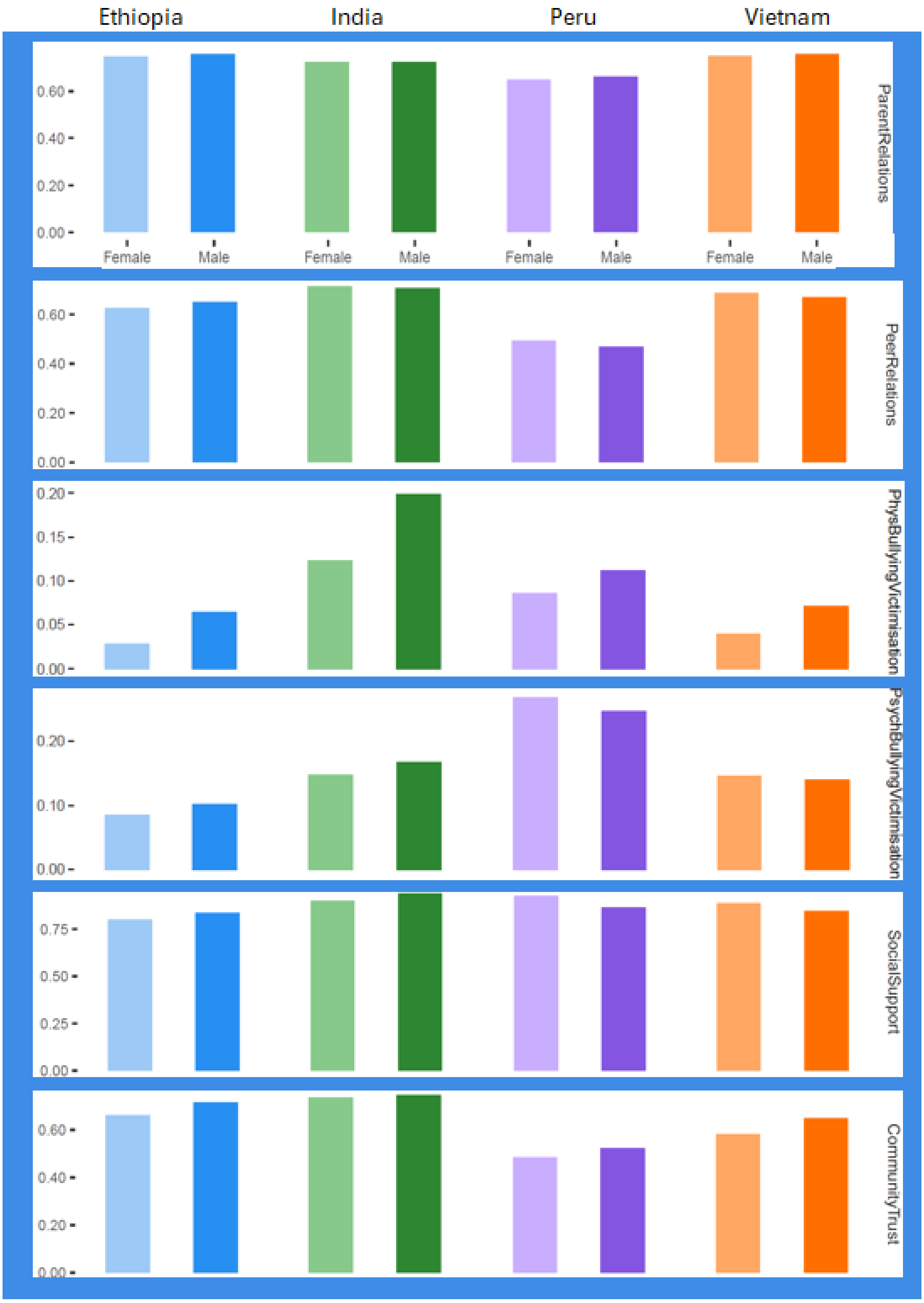

***Environmental Factors***

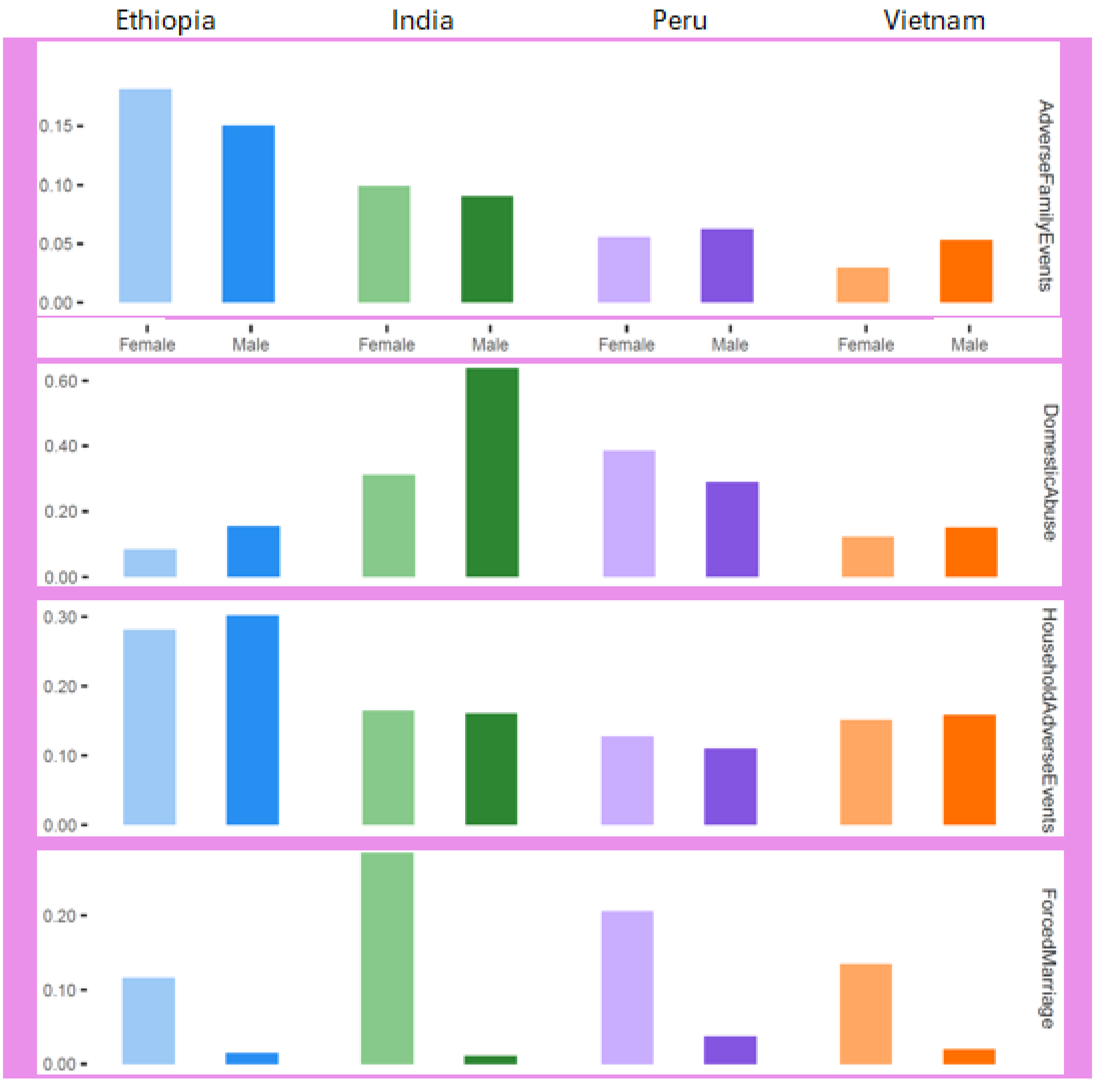

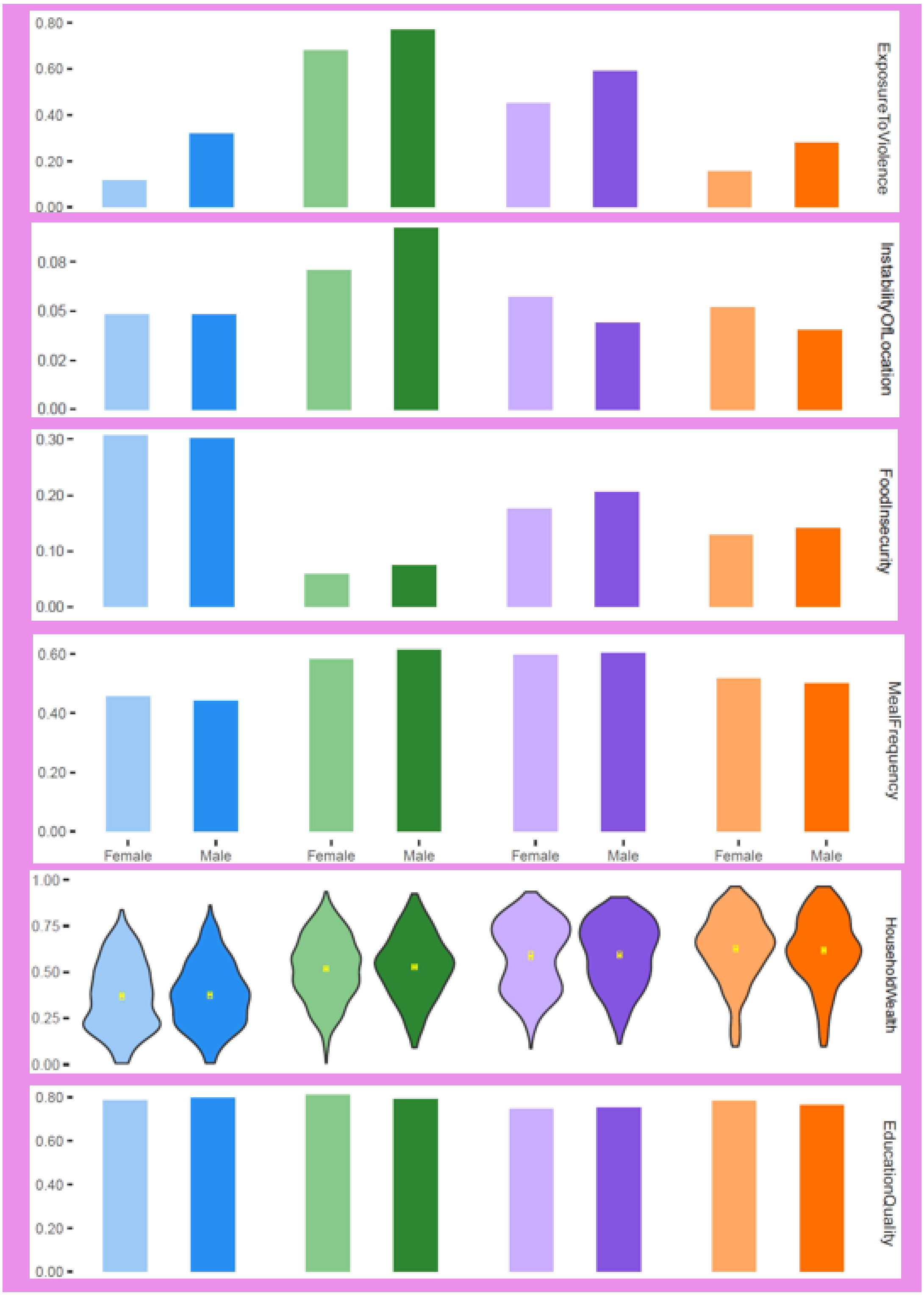

ExposureToViolence
0.00
0.20
0.40
0.60
0.80
InstabilityOfLocation
0.00
0.02
0.05
0.08
FoodInsecurity
0.00
0.10
0.20
0.30
MealFrequency
0.00
0.20
0.40
0.60
Female
Male
Female
Male
Female
Male
Female
Male
HouseholdWealth
0.00
0.25
0.50
0.75
1.00
EducationQuality
0.00
0.20
0.40
0.60
0.80

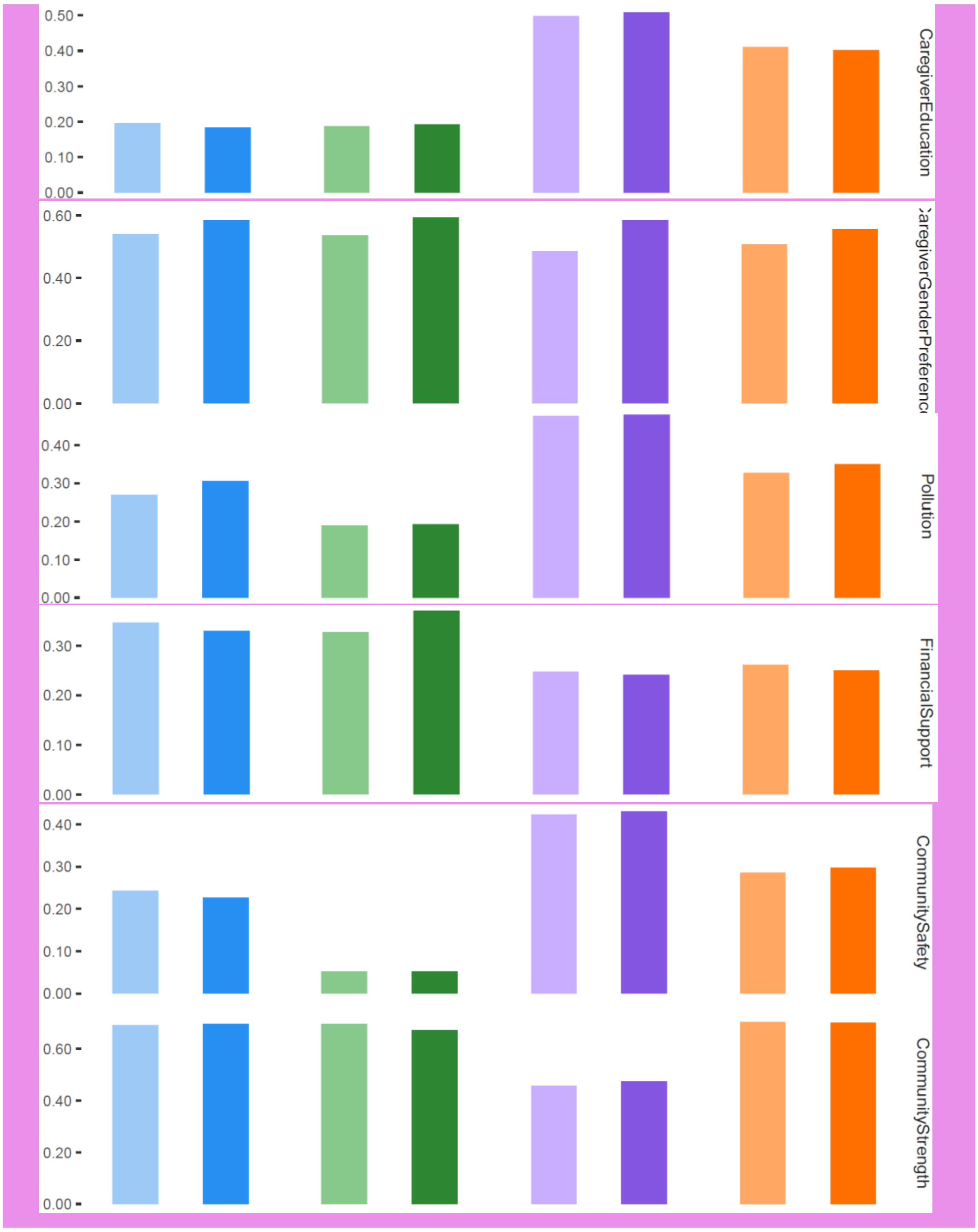
0.50
0.40
0.30
0.20
0.10
0.00
CaregiverEducation
0.60
0.40
0.20
0.00
CaregiverGenderPreferenc
0.40
0.30
0.20
0.10
0.00
Pollution
0.30
0.20
0.10
0.00
FinancialSupport
0.40
0.30
0.20
0.10
0.00
CommunitySafety
0.60
0.40
0.20
0.00
CommunityStrength

***Broader Societal Factors***

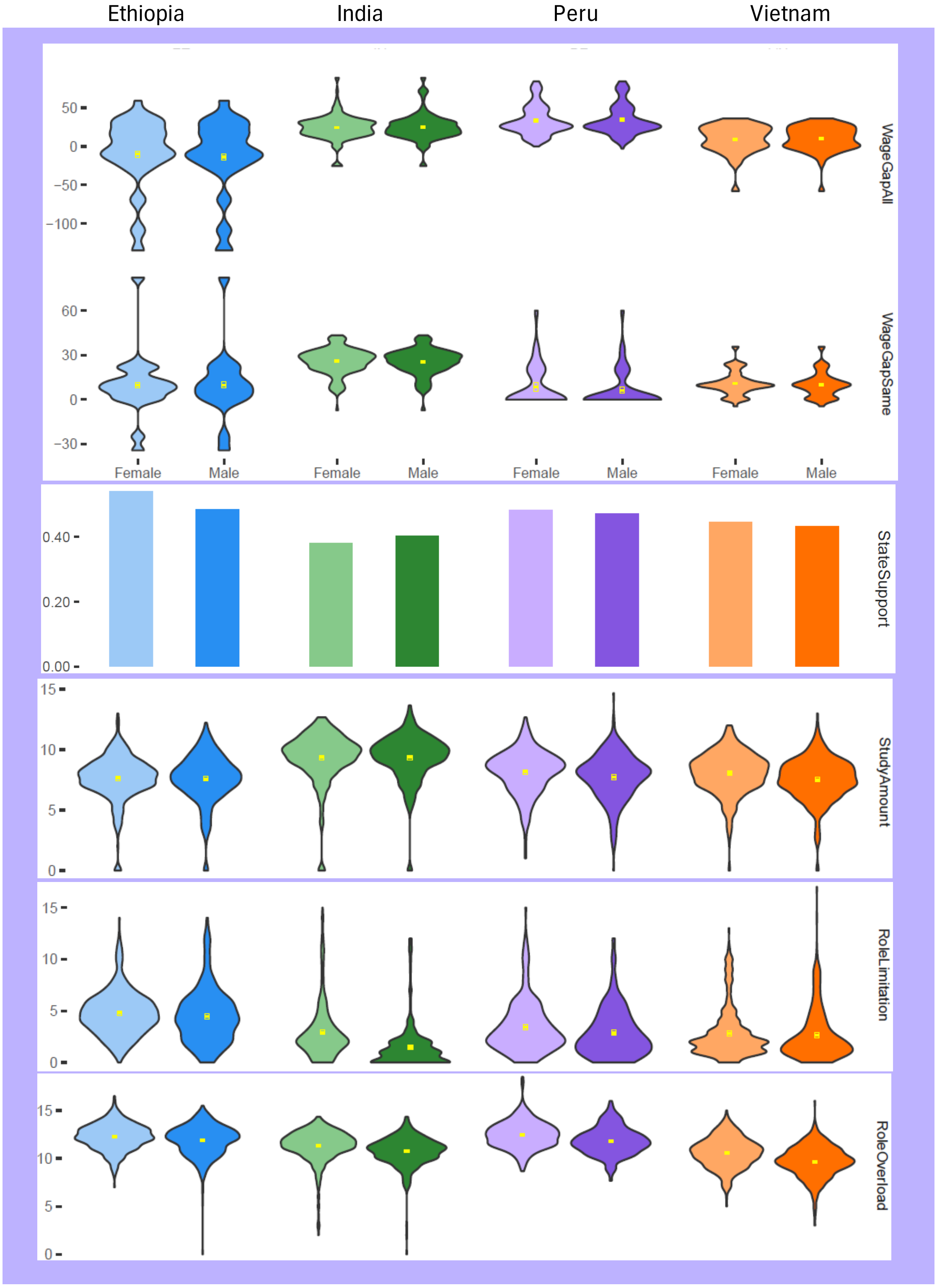

### Supplementary Table S4: Regression Results

Coefficients for regressions of each explanatory factor as independent variable and psychological distress as dependent variable. Numbers in bold are those for which the regression coefficient is different to zero at 0.05 significance.  Highlighted cells are those for which the interaction term in regressions of gender and interaction between gender and each explanatory factor as independent variable and psychological distress as dependent variable is different to zero at 0.10 significance.

| | Ethiopia | | | | | | India | | | | | | Peru | | | | | | Vietnam | | | | | |
|---|---|---|---|---|---|---|---|---|---|---|---|---|---|---|---|---|---|---|---|---|---|---|---|---|
| | Male | | | Female | | | Male | | | Female | | | Male | | | Female | | | Male | | | Female | | |
| **Variable** | **Coeff** | **CI-** | **CI+** | **Coeff** | **CI-** | **CI+** | **Coeff** | **CI-** | **CI+** | **Coeff** | **CI-** | **CI+** | **Coeff** | **CI-** | **CI+** | **Coeff** | **CI-** | **CI+** | **Coeff** | **CI-** | **CI+** | **Coeff** | **CI-** | **CI+** |
| BMI | -0.02 | -0.07 | 0.02 | 0.02 | -0.01 | 0.05 | -0.01 | -0.04 | 0.02 | **-0.05** | **-0.08** | **-0.01** | 0.00 | -0.04 | 0.03 | 0.02 | -0.02 | 0.06 | -0.01 | -0.05 | 0.03 | 0.02 | -0.02 | 0.07 |
| Early Puberty | -0.03 | -0.50 | 0.43 | 0.15 | -0.48 | 0.77 | -0.05 | -0.25 | 0.15 | 0.00 | -0.20 | 0.20 | -0.03 | -0.23 | 0.17 | -0.08 | -0.33 | 0.17 | 0.15 | -0.13 | 0.42 | 0.01 | -0.22 | 0.25 |
| Maths Score | 0.00 | 0.00 | 0.00 | 0.00 | 0.00 | 0.00 | **0.00** | **0.00** | **0.00** | **0.00** | **0.00** | **0.00** | 0.00 | 0.00 | 0.00 | 0.00 | 0.00 | 0.00 | **0.00** | **0.00** | **0.00** | **0.00** | **0.00** | **0.00** |
| Vocab Score | **-0.09** | **-0.17** | **0.00** | -0.05 | -0.13 | 0.03 | **-0.11** | **-0.19** | **-0.03** | **-0.21** | **-0.29** | **-0.13** | -0.11 | -0.27 | 0.06 | -0.14 | -0.34 | 0.05 | -0.03 | -0.12 | 0.05 | -0.01 | -0.09 | 0.07 |
| Highest Education Grade | -0.10 | -0.24 | 0.03 | -0.04 | -0.15 | 0.08 | **-0.15** | **-0.26** | **-0.05** | **-0.26** | **-0.37** | **-0.16** | -0.04 | -0.23 | 0.15 | -0.15 | -0.39 | 0.10 | **-0.29** | **-0.44** | **-0.14** | **-0.43** | **-0.58** | **-0.28** |
| Self Efficacy | 0.02 | -0.61 | 0.66 | -0.22 | -0.90 | 0.47 | **-1.46** | **-2.04** | **-0.88** | **-0.61** | **-1.20** | **-0.01** | 0.30 | -0.57 | 1.17 | 0.50 | -0.66 | 1.66 | -0.50 | -1.30 | 0.31 | **-0.93** | **-1.60** | **-0.26** |
| Self Esteem | **0.57** | **0.08** | **1.06** | **-0.50** | **-0.96** | **-0.05** | **-0.59** | **-1.09** | **-0.09** | -0.07 | -0.70 | 0.56 | -0.59 | -1.61 | 0.43 | -0.49 | -1.57 | 0.59 | -0.44 | -1.23 | 0.35 | -0.17 | -0.85 | 0.51 |
| Proscociality | -0.03 | -0.50 | 0.44 | -0.28 | -0.76 | 0.20 | **-0.84** | **-1.25** | **-0.42** | **-0.42** | **-0.82** | **-0.02** | -0.43 | -0.90 | 0.04 | -0.17 | -0.77 | 0.43 | -0.30 | -0.65 | 0.05 | -0.09 | -0.44 | 0.26 |
| Sleep Amount | -0.08 | -0.18 | 0.02 | 0.03 | -0.05 | 0.12 | 0.03 | -0.06 | 0.12 | **0.23** | **0.14** | **0.32** | -0.01 | -0.09 | 0.08 | -0.09 | -0.19 | 0.02 | 0.07 | -0.01 | 0.15 | **0.19** | **0.10** | **0.27** |
| Alcohol Consumption | 0.03 | -0.04 | 0.10 | -0.03 | -0.10 | 0.05 | 0.04 | -0.06 | 0.15 | 0.10 | -0.08 | 0.27 | 0.03 | -0.08 | 0.14 | 0.04 | -0.08 | 0.16 | 0.02 | -0.07 | 0.12 | -0.07 | -0.20 | 0.05 |
| Parent Relations | -0.04 | -0.12 | 0.04 | 0.06 | -0.02 | 0.14 | 0.07 | -0.01 | 0.15 | -0.07 | -0.15 | 0.01 | -0.06 | -0.16 | 0.04 | -0.07 | -0.18 | 0.05 | **-0.11** | **-0.21** | **0.00** | -0.08 | -0.18 | 0.02 |
| Peer Relations | 0.19 | -0.37 | 0.75 | -0.10 | -0.67 | 0.46 | **-1.15** | **-1.67** | **-0.63** | -0.48 | -1.03 | 0.06 | -0.09 | -1.10 | 0.92 | -0.49 | -1.79 | 0.81 | **-0.80** | **-1.47** | **-0.13** | -0.49 | -1.16 | 0.18 |
| Phys Bullying Victimisation | -0.01 | -0.66 | 0.64 | -0.19 | -0.97 | 0.59 | **0.43** | **0.13** | **0.74** | **0.88** | **0.48** | **1.29** | **0.75** | **0.20** | **1.30** | 0.12 | -0.60 | 0.84 | **0.72** | **0.16** | **1.28** | 0.42 | -0.27 | 1.11 |
| Psych Bullying Victimisation | 0.21 | -0.40 | 0.82 | -0.07 | -0.64 | 0.50 | 0.37 | -0.05 | 0.79 | **0.74** | **0.23** | **1.25** | **0.91** | **0.40** | **1.41** | **0.63** | **0.04** | **1.22** | 0.30 | -0.26 | 0.85 | **0.75** | **0.20** | **1.29** |
| Social Support | 0.01 | -0.02 | 0.05 | -0.03 | -0.06 | 0.00 | 0.01 | -0.04 | 0.07 | -0.04 | -0.09 | 0.00 | -0.02 | -0.06 | 0.03 | -0.06 | -0.13 | 0.01 | **-0.05** | **-0.09** | **0.00** | -0.05 | -0.09 | 0.00 |
| Community Trust | -0.08 | -0.64 | 0.47 | -0.07 | -0.57 | 0.44 | **-0.96** | **-1.56** | **-0.37** | 0.00 | -0.59 | 0.58 | **-0.99** | **-1.68** | **-0.30** | -0.06 | -0.82 | 0.70 | 0.17 | -0.35 | 0.69 | 0.13 | -0.30 | 0.56 |
| Adverse Family Events | 0.04 | -0.14 | 0.21 | 0.09 | -0.06 | 0.24 | -0.08 | -0.28 | 0.11 | **0.19** | **0.00** | **0.38** | 0.11 | -0.18 | 0.40 | 0.19 | -0.21 | 0.59 | **0.41** | **0.12** | **0.70** | 0.27 | -0.11 | 0.64 |
| Domestic Abuse | -0.01 | -0.27 | 0.24 | 0.12 | -0.20 | 0.45 | **0.23** | **0.06** | **0.40** | 0.18 | -0.01 | 0.37 | -0.04 | -0.26 | 0.18 | 0.03 | -0.22 | 0.28 | 0.19 | -0.08 | 0.46 | 0.16 | -0.12 | 0.43 |
| Household Adverse Events | -0.32 | -0.89 | 0.25 | 0.06 | -0.54 | 0.67 | 0.44 | -0.46 | 1.35 | 0.69 | -0.13 | 1.51 | 1.01 | -0.37 | 2.40 | 1.14 | -0.32 | 2.61 | 0.58 | -0.44 | 1.60 | 0.31 | -0.70 | 1.32 |
| Forced Marriage | -0.40 | -1.17 | 0.38 | 0.05 | -0.23 | 0.33 | 0.32 | -0.46 | 1.10 | 0.18 | -0.01 | 0.37 | 0.01 | -0.52 | 0.55 | -0.21 | -0.51 | 0.08 | 0.46 | -0.24 | 1.15 | **0.36** | **0.09** | **0.63** |
| Exposure To Violence | 0.15 | -0.04 | 0.35 | 0.06 | -0.22 | 0.34 | **0.27** | **0.08** | **0.46** | **0.21** | **0.03** | **0.40** | **0.26** | **0.06** | **0.47** | 0.09 | -0.16 | 0.34 | 0.15 | -0.07 | 0.36 | **0.34** | **0.09** | **0.60** |
| Instability Of Location | **1.56** | **0.47** | **2.64** | 0.81 | -0.34 | 1.95 | -0.09 | -0.70 | 0.51 | 0.58 | -0.45 | 1.61 | **1.48** | **0.20** | **2.75** | 0.87 | -0.74 | 2.49 | -0.35 | -2.35 | 1.64 | 1.53 | -0.49 | 3.55 |
| Food Insecurity | 0.03 | -0.04 | 0.10 | 0.04 | -0.03 | 0.11 | 0.04 | -0.09 | 0.18 | **0.23** | **0.07** | **0.38** | 0.07 | -0.02 | 0.17 | 0.09 | -0.03 | 0.22 | **0.18** | **0.07** | **0.30** | **0.14** | **0.03** | **0.26** |
| Meal Frequency | -0.12 | -0.26 | 0.02 | -0.09 | -0.22 | 0.05 | **0.10** | **0.03** | **0.18** | -0.01 | -0.09 | 0.07 | -0.09 | -0.19 | 0.00 | **-0.13** | **-0.24** | **-0.02** | **-0.11** | **-0.20** | **-0.01** | -0.01 | -0.10 | 0.08 |
| Household Wealth | -0.19 | -0.72 | 0.34 | -0.29 | -0.77 | 0.19 | -0.08 | -0.55 | 0.39 | **-0.58** | **-1.09** | **-0.08** | -0.15 | -0.71 | 0.41 | -0.36 | -1.02 | 0.30 | **-1.12** | **-1.63** | **-0.61** | **-0.79** | **-1.29** | **-0.29** |
| Education Quality | 0.23 | -0.30 | 0.75 | -0.17 | -0.66 | 0.31 | -0.38 | -0.88 | 0.11 | -0.45 | -0.97 | 0.07 | **0.61** | **-0.01** | **1.22** | -0.23 | -0.90 | 0.43 | 0.26 | -0.31 | 0.82 | 0.24 | -0.35 | 0.84 |
| Caregiver Education | -0.02 | -0.14 | 0.09 | -0.06 | -0.16 | 0.04 | -0.07 | -0.16 | 0.02 | **-0.15** | **-0.24** | **-0.06** | -0.04 | -0.12 | 0.05 | -0.09 | -0.19 | 0.02 | **-0.17** | **-0.27** | **-0.07** | **-0.24** | **-0.34** | **-0.14** |
| Caregiver Gender Pref | -0.04 | -0.18 | 0.09 | 0.04 | -0.11 | 0.18 | 0.06 | -0.09 | 0.21 | 0.12 | -0.03 | 0.28 | -0.03 | -0.19 | 0.13 | 0.05 | -0.14 | 0.24 | -0.10 | -0.27 | 0.08 | 0.00 | -0.17 | 0.18 |
| Pollution | -0.02 | -0.40 | 0.36 | 0.23 | -0.16 | 0.63 | -0.21 | -0.62 | 0.20 | 0.04 | -0.45 | 0.52 | 0.20 | -0.34 | 0.73 | -0.16 | -0.86 | 0.53 | 0.07 | -0.42 | 0.57 | 0.46 | -0.01 | 0.94 |
| Financial Support | -0.31 | -0.82 | 0.20 | -0.29 | -0.76 | 0.19 | **-0.89** | **-1.35** | **-0.43** | -0.30 | -0.82 | 0.22 | -0.55 | -1.17 | 0.07 | 0.28 | -0.59 | 1.14 | 0.00 | -0.65 | 0.65 | **-0.86** | **-1.53** | **-0.20** |
| Community Safety | -0.01 | -0.39 | 0.37 | 0.13 | -0.24 | 0.49 | **1.22** | **0.21** | **2.23** | 0.62 | -0.58 | 1.82 | **-0.30** | **-0.60** | **-0.01** | -0.32 | -0.68 | 0.04 | 0.20 | -0.21 | 0.61 | 0.04 | -0.35 | 0.44 |
| Community Strength | 0.08 | -0.50 | 0.65 | -0.36 | -0.95 | 0.23 | **-0.65** | **-1.24** | **-0.05** | -0.63 | -1.28 | 0.02 | 0.48 | -0.26 | 1.22 | -0.42 | -1.31 | 0.47 | -0.18 | -1.00 | 0.65 | 0.14 | -0.63 | 0.91 |
| State Support | **0.42** | **0.06** | **0.77** | -0.14 | -0.46 | 0.18 | -0.23 | -0.58 | 0.12 | **-0.53** | **-0.87** | **-0.18** | -0.37 | -0.78 | 0.03 | -0.47 | -0.97 | 0.02 | -0.37 | -0.85 | 0.12 | 0.15 | -0.29 | 0.59 |
| Study Amount | 0.01 | -0.04 | 0.05 | -0.04 | -0.09 | 0.01 | **-0.06** | **-0.10** | **-0.01** | **-0.05** | **-0.10** | **-0.01** | -0.01 | -0.06 | 0.05 | **-0.11** | **-0.18** | **-0.05** | **-0.09** | **-0.15** | **-0.04** | **-0.11** | **-0.16** | **-0.05** |
| Role Limitation | -0.01 | -0.05 | 0.02 | 0.01 | -0.04 | 0.05 | **0.07** | **0.03** | **0.11** | **0.04** | **0.00** | **0.07** | 0.04 | 0.00 | 0.08 | 0.04 | -0.01 | 0.08 | **0.05** | **0.01** | **0.09** | 0.03 | -0.01 | 0.07 |
| Role Overload | -0.01 | -0.07 | 0.05 | -0.01 | -0.07 | 0.06 | -0.02 | -0.07 | 0.03 | -0.04 | -0.10 | 0.01 | **0.09** | **0.02** | **0.16** | -0.03 | -0.11 | 0.06 | **-0.11** | **-0.17** | **-0.05** | **-0.10** | **-0.16** | **-0.04** |
| Wage Gap Same | 0.00 | 0.00 | 0.01 | 0.00 | -0.01 | 0.00 | **0.01** | **0.00** | **0.02** | **0.01** | **0.00** | **0.02** | 0.00 | -0.01 | 0.01 | -0.01 | -0.03 | 0.00 | **0.02** | **0.00** | **0.03** | 0.01 | 0.00 | 0.02 |
| Wage Gap All | 0.00 | 0.00 | 0.00 | 0.00 | 0.00 | 0.00 | 0.00 | 0.00 | 0.01 | **0.01** | **0.00** | **0.01** | 0.00 | 0.00 | 0.01 | 0.00 | 0.00 | 0.01 | **0.01** | **0.00** | **0.01** | **0.01** | **0.00** | **0.01** |

### Supplementary Figures S3: Predicted margin plots for the interaction terms which are significantly different to zero at 0.1 confidence

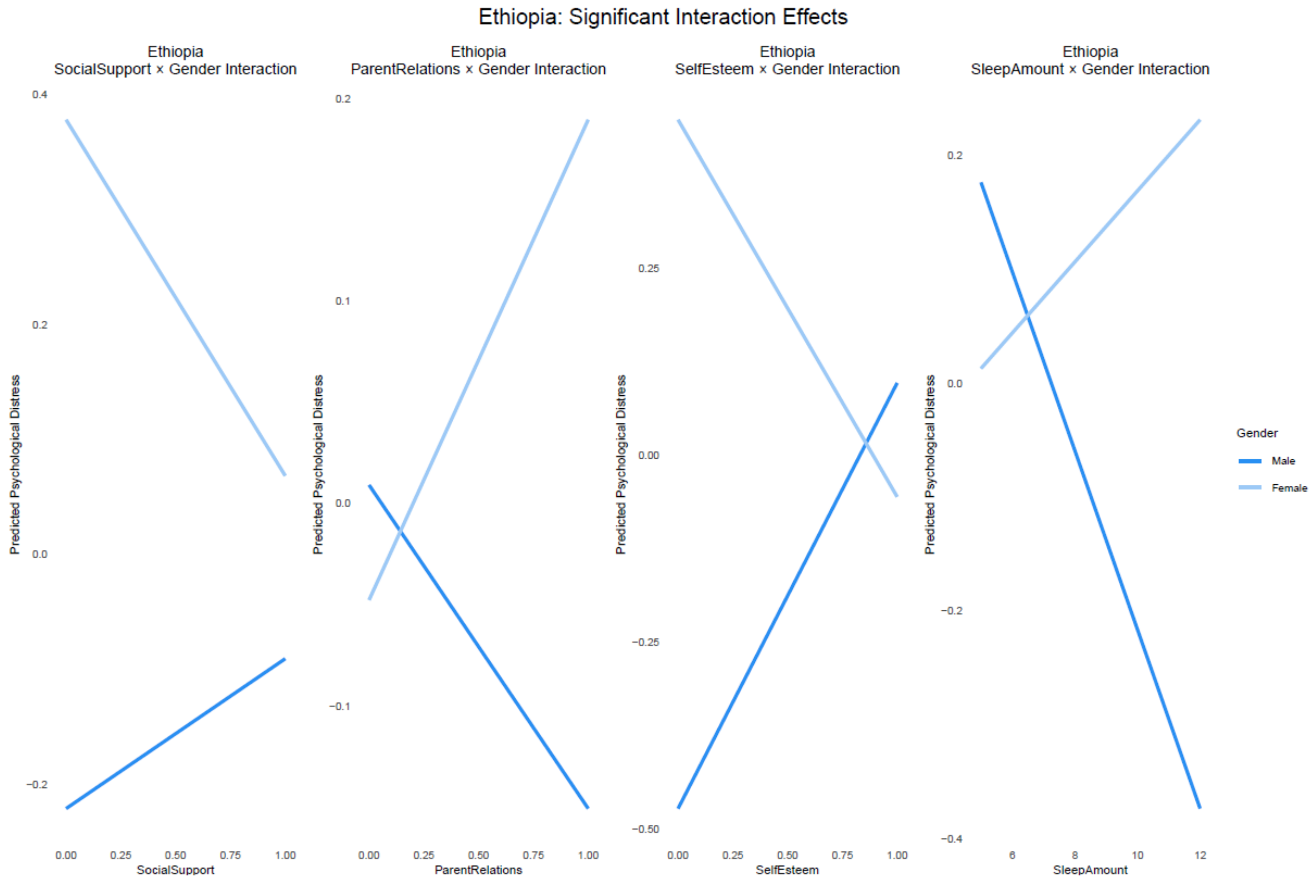
Ethiopia: Significant Interaction Effects
Ethiopia
SocialSupport × Gender Interaction
Ethiopia
ParentRelations × Gender Interaction
Ethiopia
SelfEsteem × Gender Interaction
Ethiopia
SleepAmount × Gender Interaction
Predicted Psychological Distress
SocialSupport
ParentRelations
SelfEsteem
SleepAmount
Gender
Male
Female

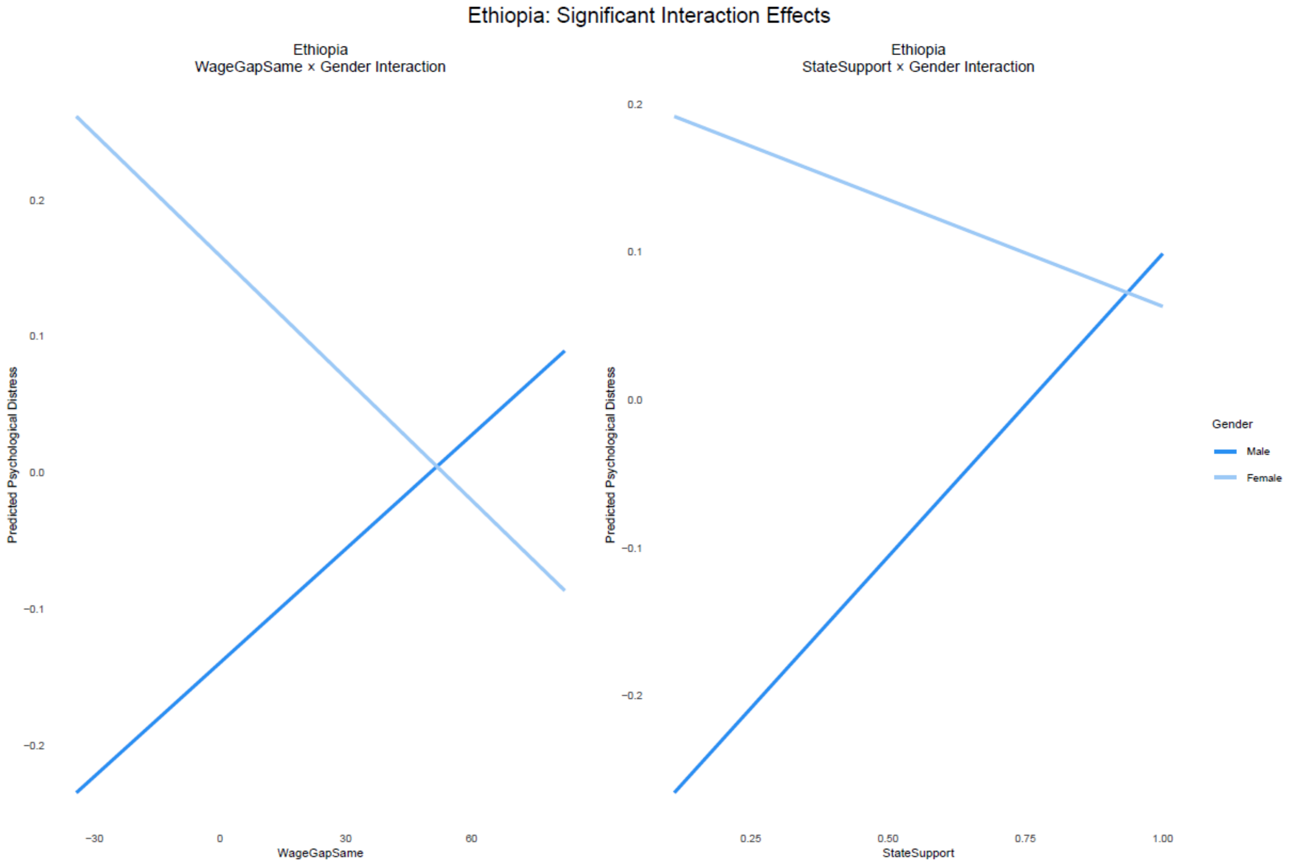
Ethiopia: Significant Interaction Effects
Ethiopia
WageGapSame × Gender Interaction
Ethiopia
StateSupport × Gender Interaction
Predicted Psychological Distress
WageGapSame
StateSupport
Gender
Male
Female

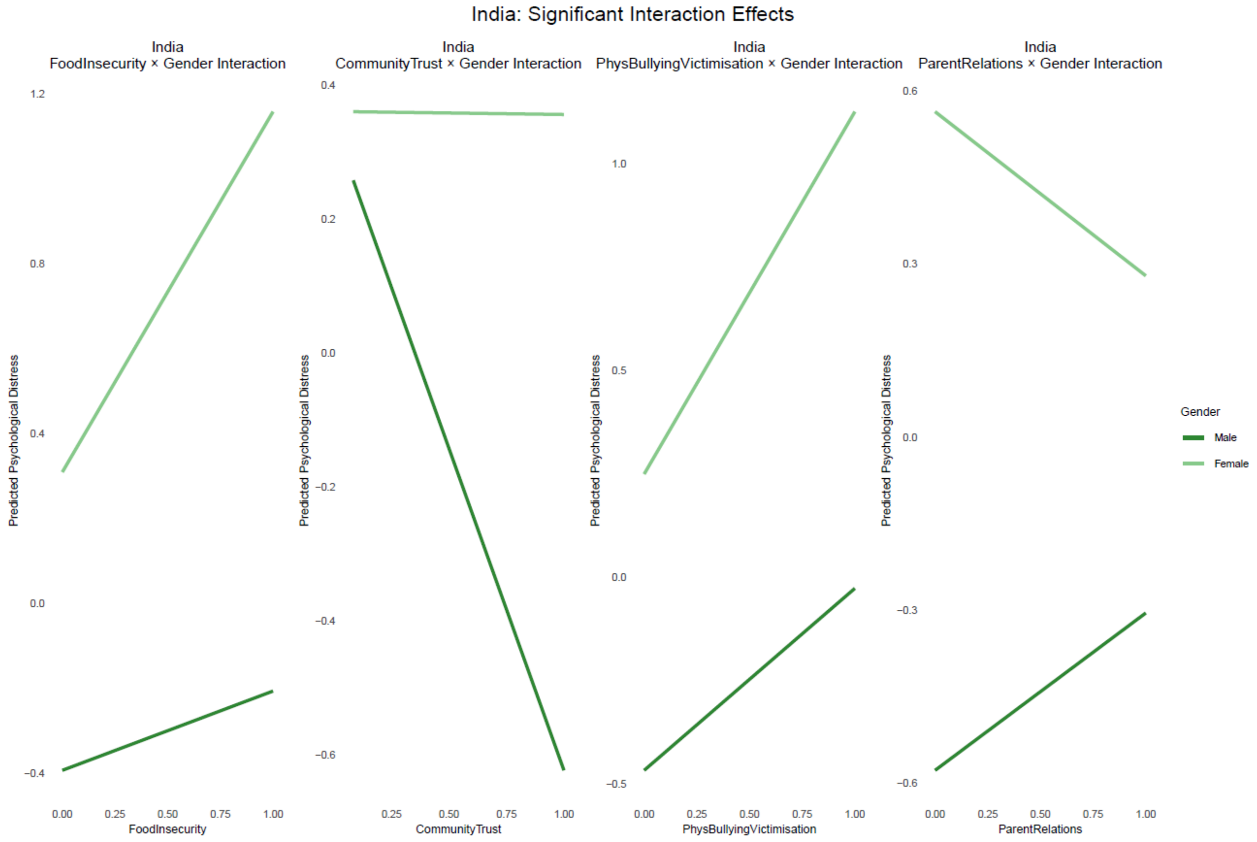

India: Significant Interaction Effects
India
FoodInsecurity × Gender Interaction
India
CommunityTrust × Gender Interaction
India
PhysBullyingVictimisation × Gender Interaction
India
ParentRelations × Gender Interaction
Predicted Psychological Distress
FoodInsecurity
CommunityTrust
PhysBullyingVictimisation
ParentRelations
Gender
Male
Female


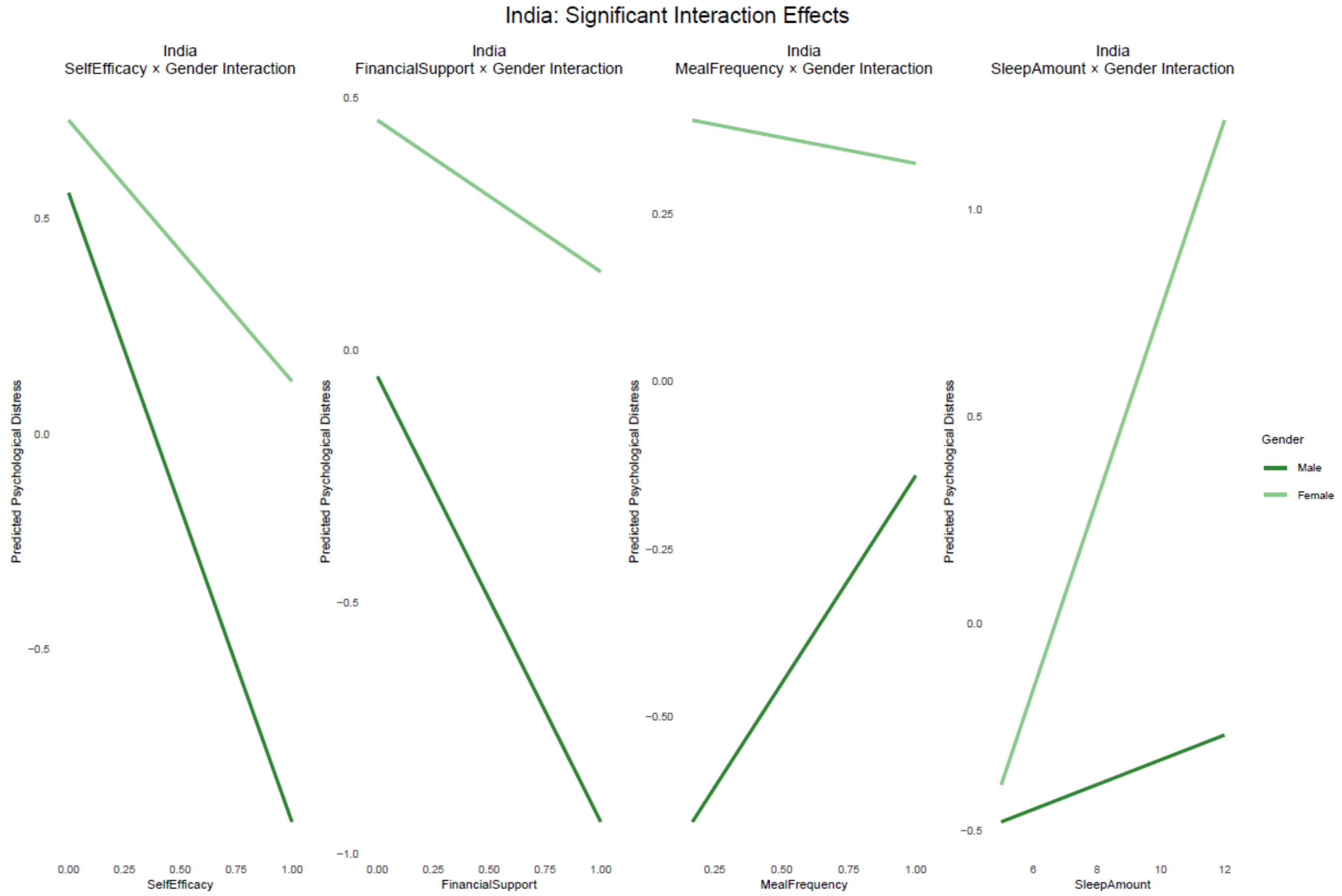

India: Significant Interaction Effects
India
SelfEfficacy × Gender Interaction
India
FinancialSupport × Gender Interaction
India
MealFrequency × Gender Interaction
India
SleepAmount × Gender Interaction
Predicted Psychological Distress
SelfEfficacy
FinancialSupport
MealFrequency
SleepAmount
Gender
Male
Female

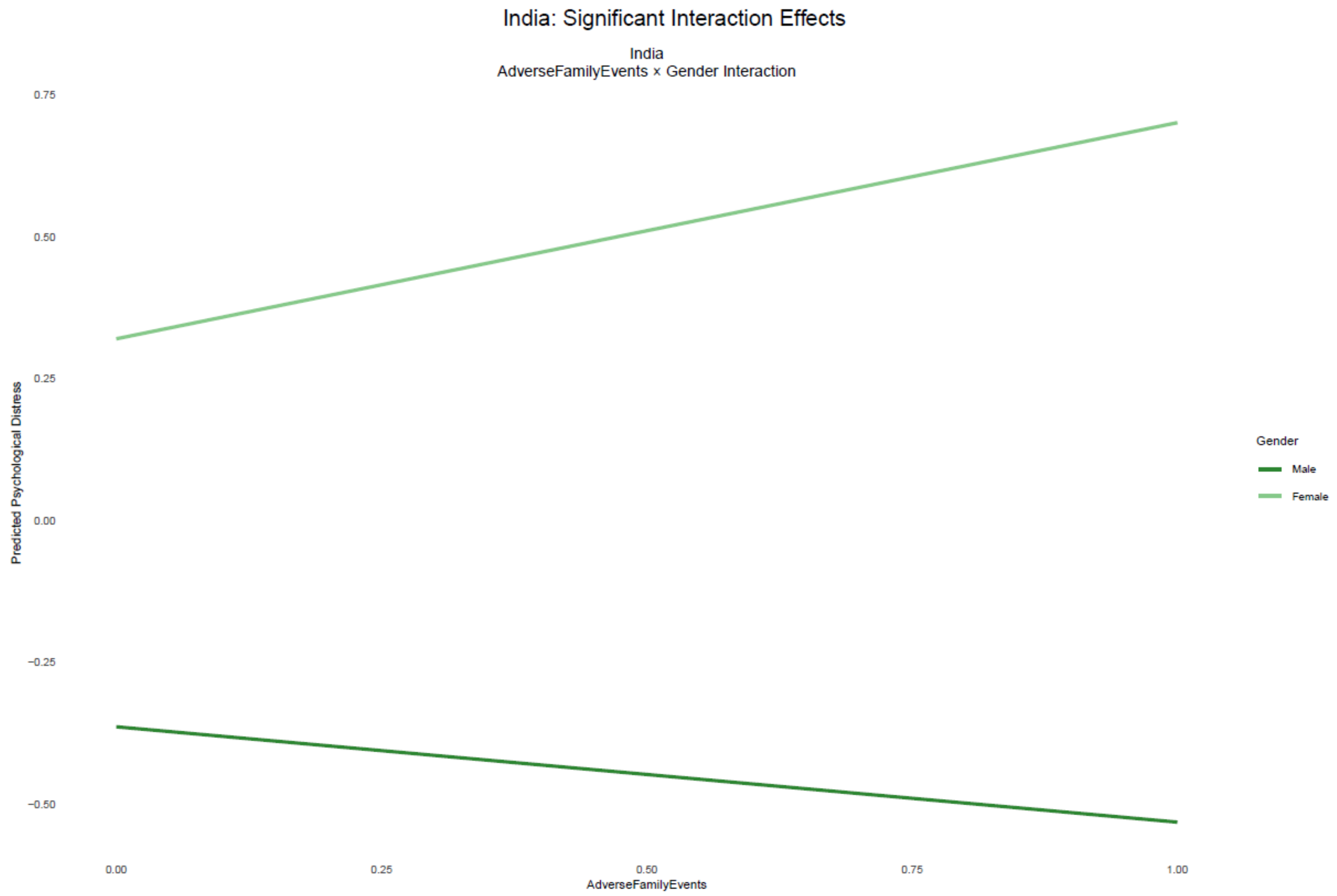
India: Significant Interaction Effects
India
AdverseFamilyEvents × Gender Interaction
Predicted Psychological Distress
0.75
0.50
0.25
0.00
−0.25
−0.50
0.00
0.25
0.50
0.75
1.00
AdverseFamilyEvents
Gender
Male
Female

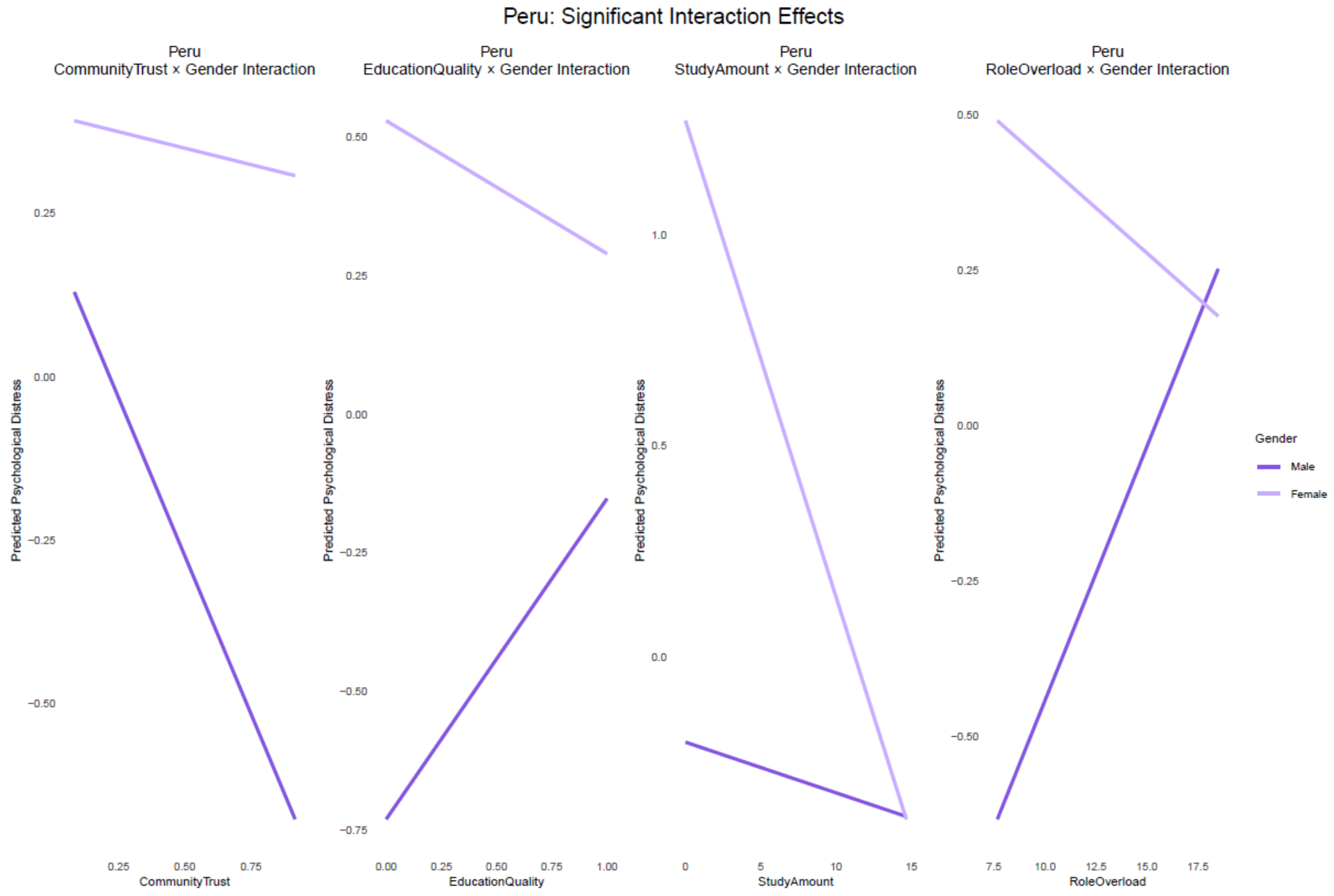
Peru: Significant Interaction Effects
Peru
CommunityTrust × Gender Interaction
Peru
EducationQuality × Gender Interaction
Peru
StudyAmount × Gender Interaction
Peru
RoleOverload × Gender Interaction
Predicted Psychological Distress
0.25
0.00
−0.25
−0.50
0.25
0.50
0.75
CommunityTrust
0.50
0.25
0.00
−0.25
−0.50
−0.75
0.00
0.25
0.50
0.75
1.00
EducationQuality
1.0
0.5
0.0
0
5
10
15
StudyAmount
0.50
0.25
0.00
−0.25
−0.50
7.5
10.0
12.5
15.0
17.5
RoleOverload
Gender
Male
Female

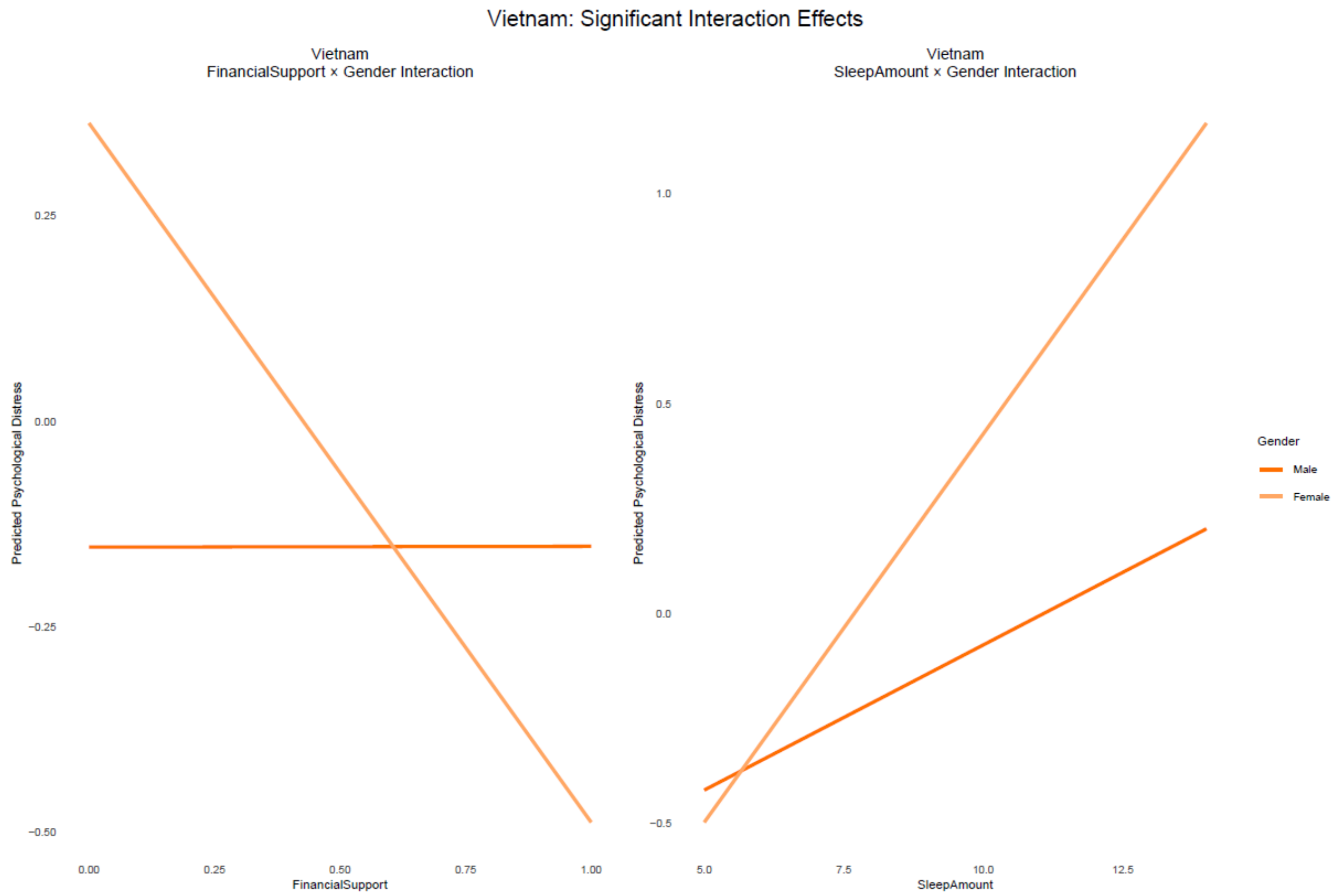


**Supplementary Figures S4: Predicted margin plots for highlighted interactions**

**Supplementary Figure S4a: Better self efficacy and self esteem have stronger associations with lower psychological distress in boys than in girls in India**

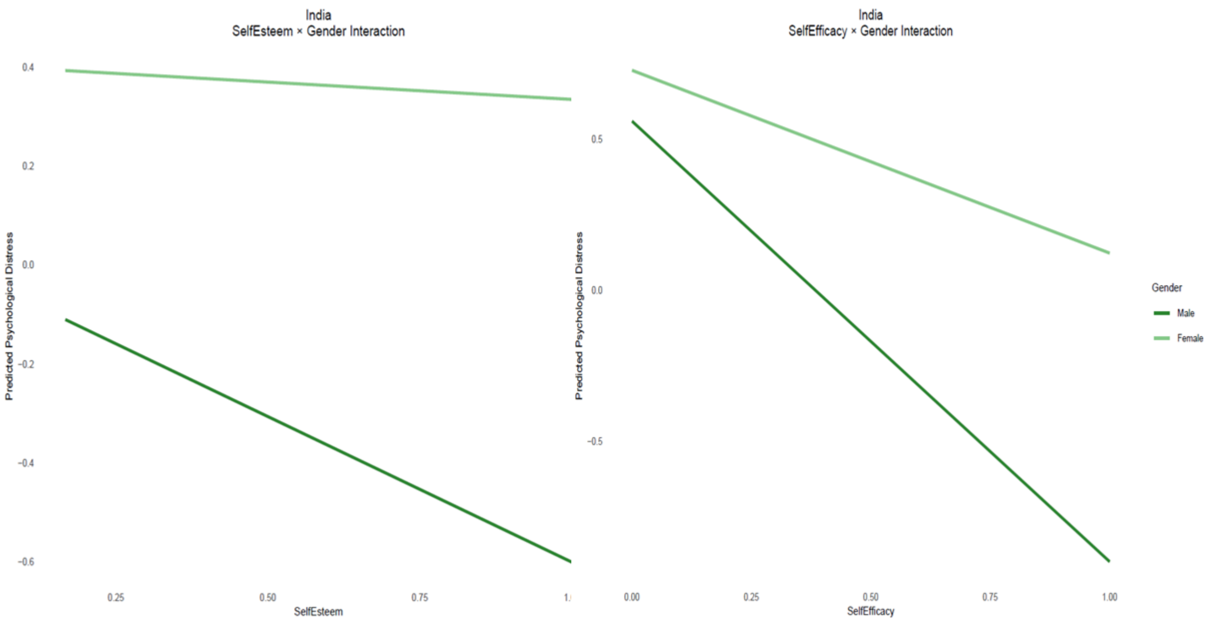


**Supplementary Figure S4b: Community trust is significantly associated with lower distress in boys but not girls in India (left chart) and Peru (right chart)**

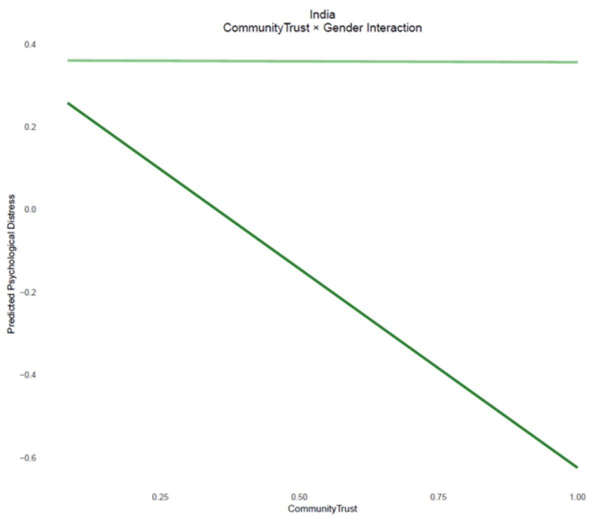
India
CommunityTrust × Gender Interaction
Predicted Psychological Distress
0.4
0.2
0.0
−0.2
−0.4
−0.6
0.25
0.50
0.75
1.00
CommunityTrust

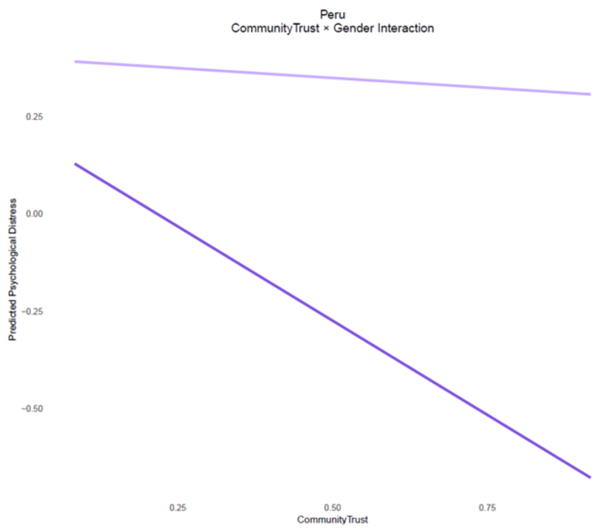
Peru
CommunityTrust × Gender Interaction
Predicted Psychological Distress
0.25
0.00
−0.25
−0.50
0.25
0.50
0.75
CommunityTrust

**Supplementary Figure S4c: Food insecurity is significantly associated with greater distress in girls but not in boys in India.**

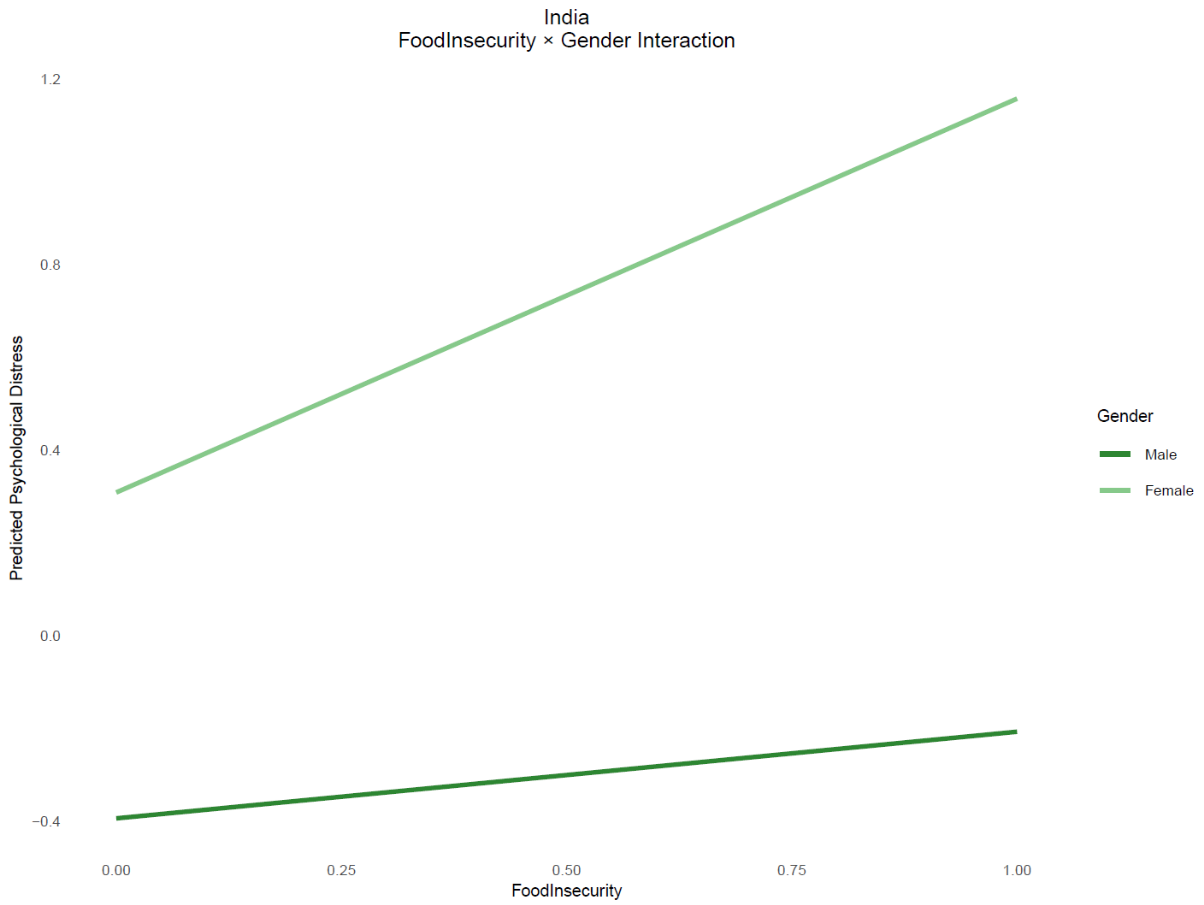

**Supplementary Figures 5: SHAP and PFI Values for Explanatory Variables in the Ensemble Tree Models for Each Country**

***Ethiopia***

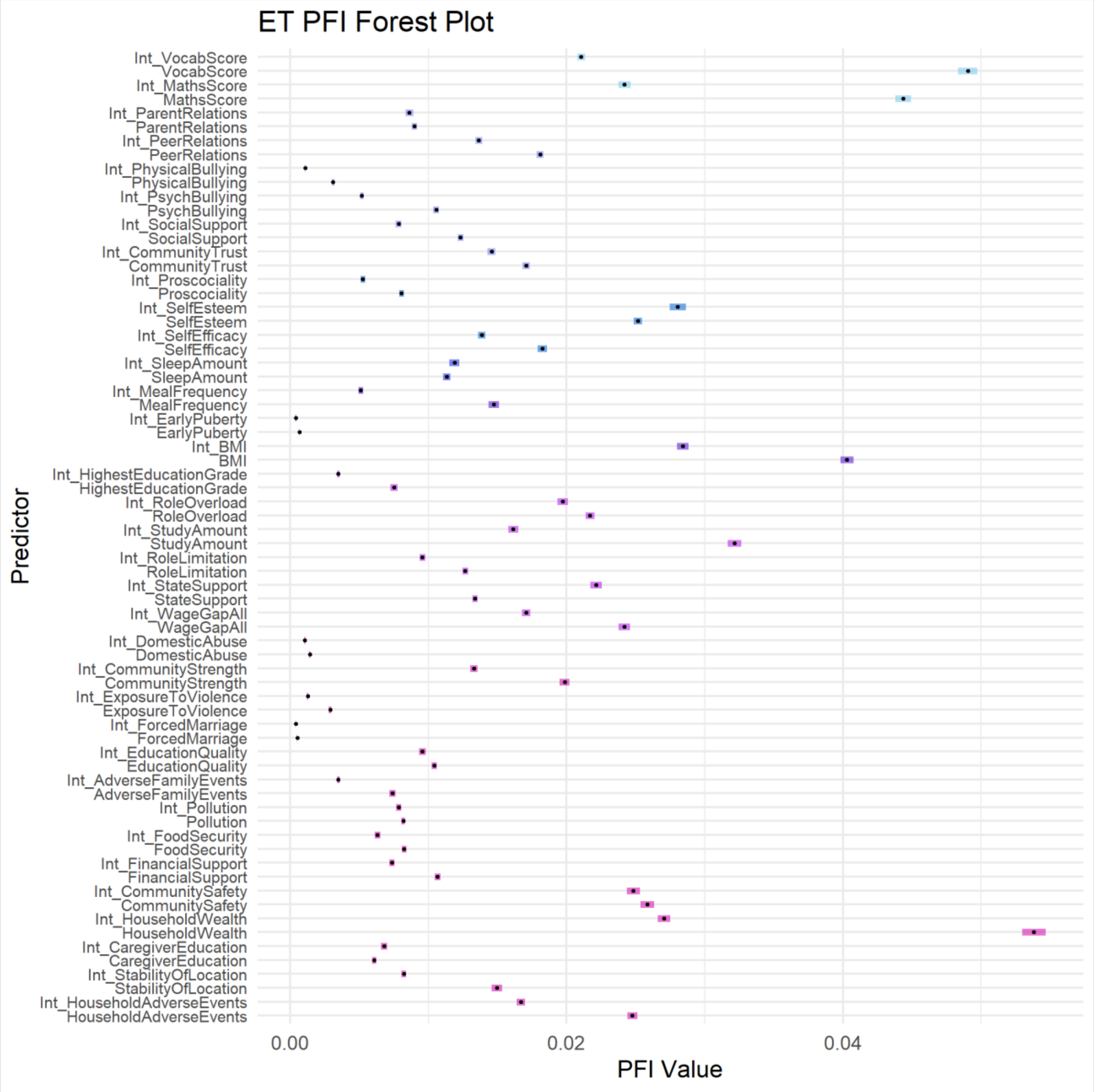

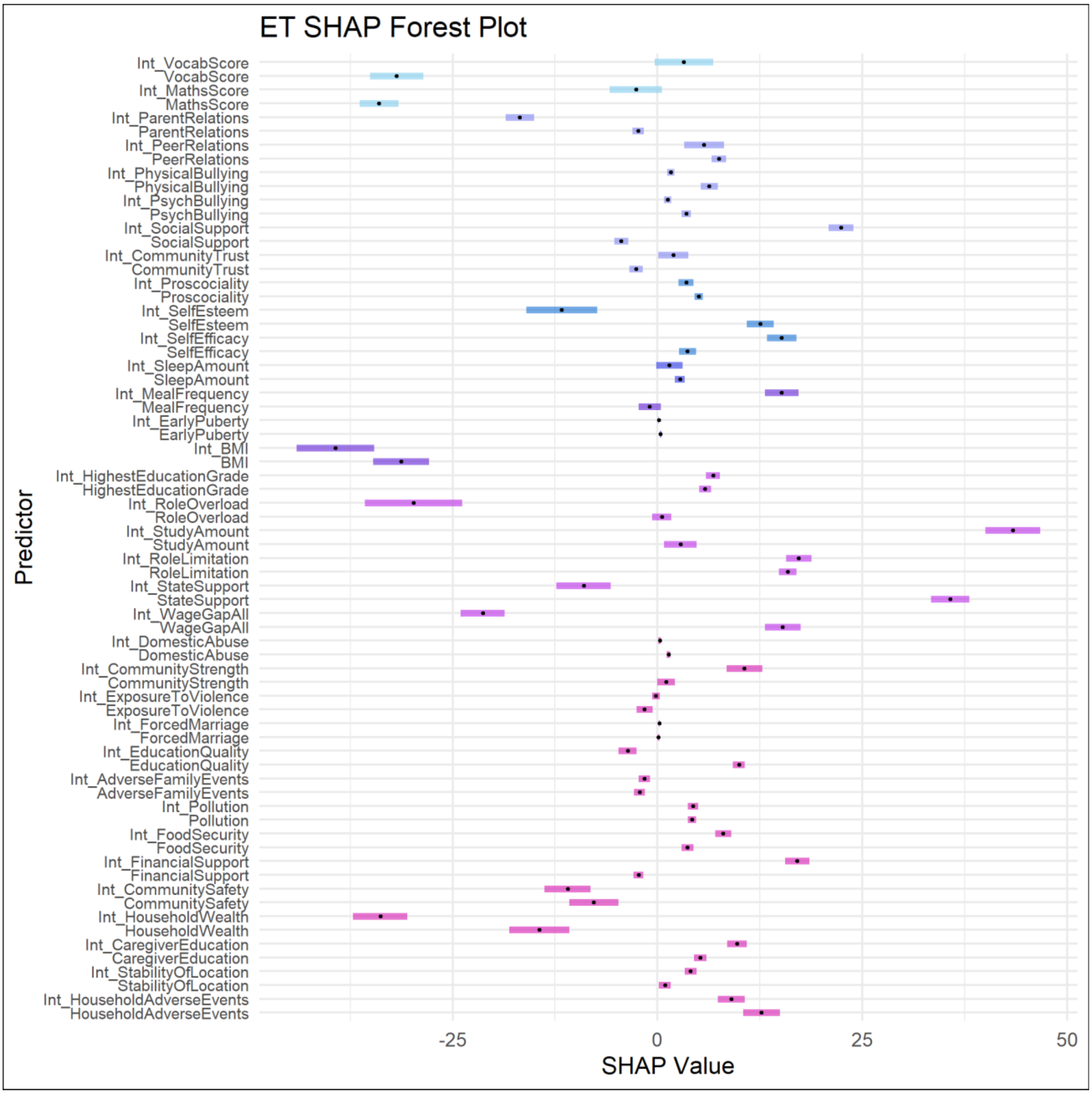

ET SHAP Forest Plot
Int_VocabScore
VocabScore
Int_MathsScore
MathsScore
Int_ParentRelations
ParentRelations
Int_PeerRelations
PeerRelations
Int_PhysicalBullying
PhysicalBullying
Int_PsychBullying
PsychBullying
Int_SocialSupport
SocialSupport
Int_CommunityTrust
CommunityTrust
Int_Proscociality
Proscociality
Int_SelfEsteem
SelfEsteem
Int_SelfEfficacy
SelfEfficacy
Int_SleepAmount
SleepAmount
Int_MealFrequency
MealFrequency
Int_EarlyPuberty
EarlyPuberty
Int_BMI
BMI
Int_HighestEducationGrade
HighestEducationGrade
Int_RoleOverload
RoleOverload
Int_StudyAmount
StudyAmount
Int_RoleLimitation
RoleLimitation
Int_StateSupport
StateSupport
Int_WageGapAll
WageGapAll
Int_DomesticAbuse
DomesticAbuse
Int_CommunityStrength
CommunityStrength
Int_ExposureToViolence
ExposureToViolence
Int_ForcedMarriage
ForcedMarriage
Int_EducationQuality
EducationQuality
Int_AdverseFamilyEvents
AdverseFamilyEvents
Int_Pollution
Pollution
Int_FoodSecurity
FoodSecurity
Int_FinancialSupport
FinancialSupport
Int_CommunitySafety
CommunitySafety
Int_HouseholdWealth
HouseholdWealth
Int_CaregiverEducation
CaregiverEducation
Int_StabilityOfLocation
StabilityOfLocation
Int_HouseholdAdverseEvents
HouseholdAdverseEvents
Predictor
-25
0
25
50
SHAP Value

***India***

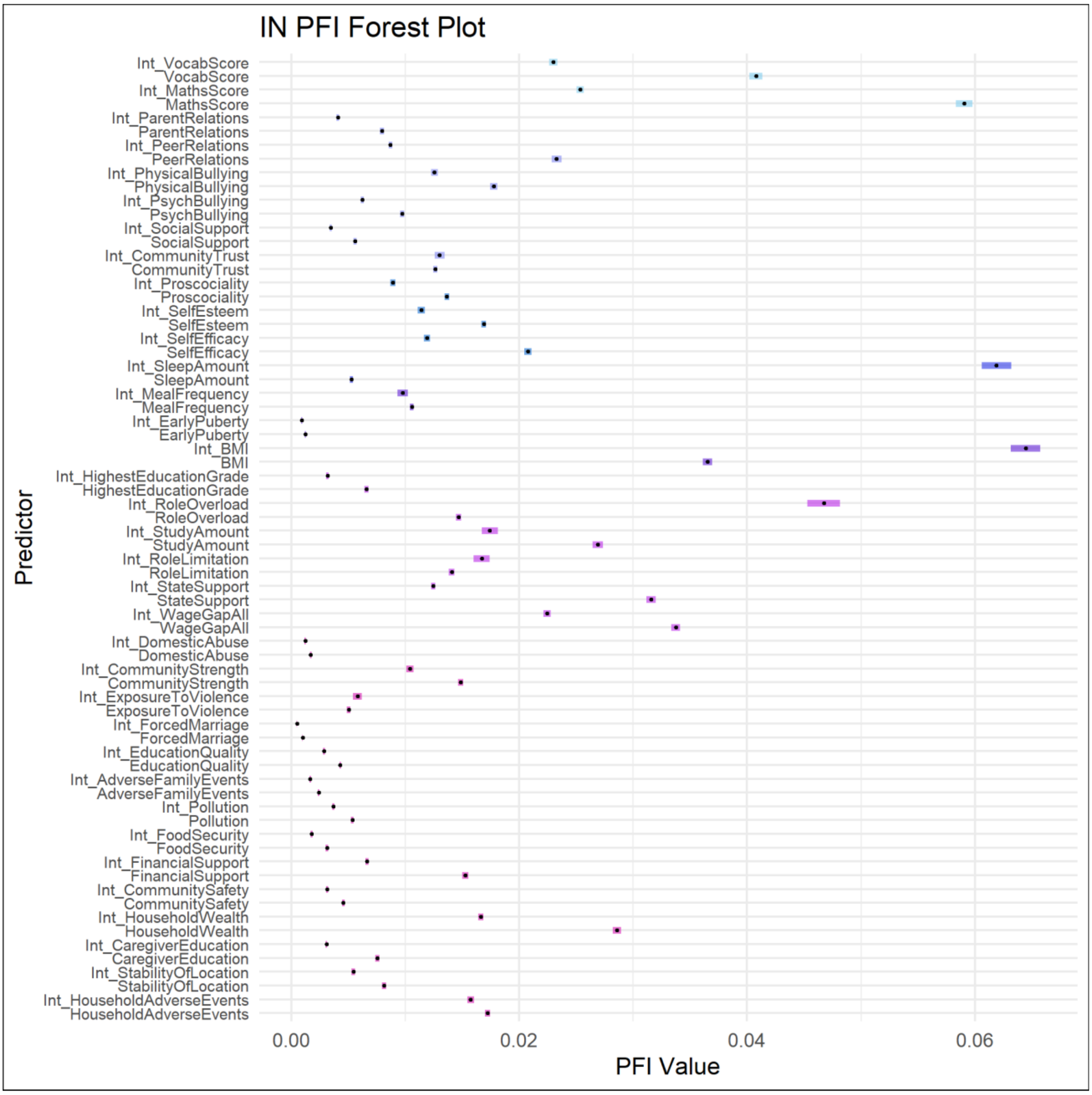

IN PFI Forest Plot
Predictor
PFI Value
0.00
0.02
0.04
0.06
Int_VocabScore
VocabScore
Int_MathsScore
MathsScore
Int_ParentRelations
ParentRelations
Int_PeerRelations
PeerRelations
Int_PhysicalBullying
PhysicalBullying
Int_PsychBullying
PsychBullying
Int_SocialSupport
SocialSupport
Int_CommunityTrust
CommunityTrust
Int_Proscociality
Proscociality
Int_SelfEsteem
SelfEsteem
Int_SelfEfficacy
SelfEfficacy
Int_SleepAmount
SleepAmount
Int_MealFrequency
MealFrequency
Int_EarlyPuberty
EarlyPuberty
Int_BMI
BMI
Int_HighestEducationGrade
HighestEducationGrade
Int_RoleOverload
RoleOverload
Int_StudyAmount
StudyAmount
Int_RoleLimitation
RoleLimitation
Int_StateSupport
StateSupport
Int_WageGapAll
WageGapAll
Int_DomesticAbuse
DomesticAbuse
Int_CommunityStrength
CommunityStrength
Int_ExposureToViolence
ExposureToViolence
Int_ForcedMarriage
ForcedMarriage
Int_EducationQuality
EducationQuality
Int_AdverseFamilyEvents
AdverseFamilyEvents
Int_Pollution
Pollution
Int_FoodSecurity
FoodSecurity
Int_FinancialSupport
FinancialSupport
Int_CommunitySafety
CommunitySafety
Int_HouseholdWealth
HouseholdWealth
Int_CaregiverEducation
CaregiverEducation
Int_StabilityOfLocation
StabilityOfLocation
Int_HouseholdAdverseEvents
HouseholdAdverseEvents

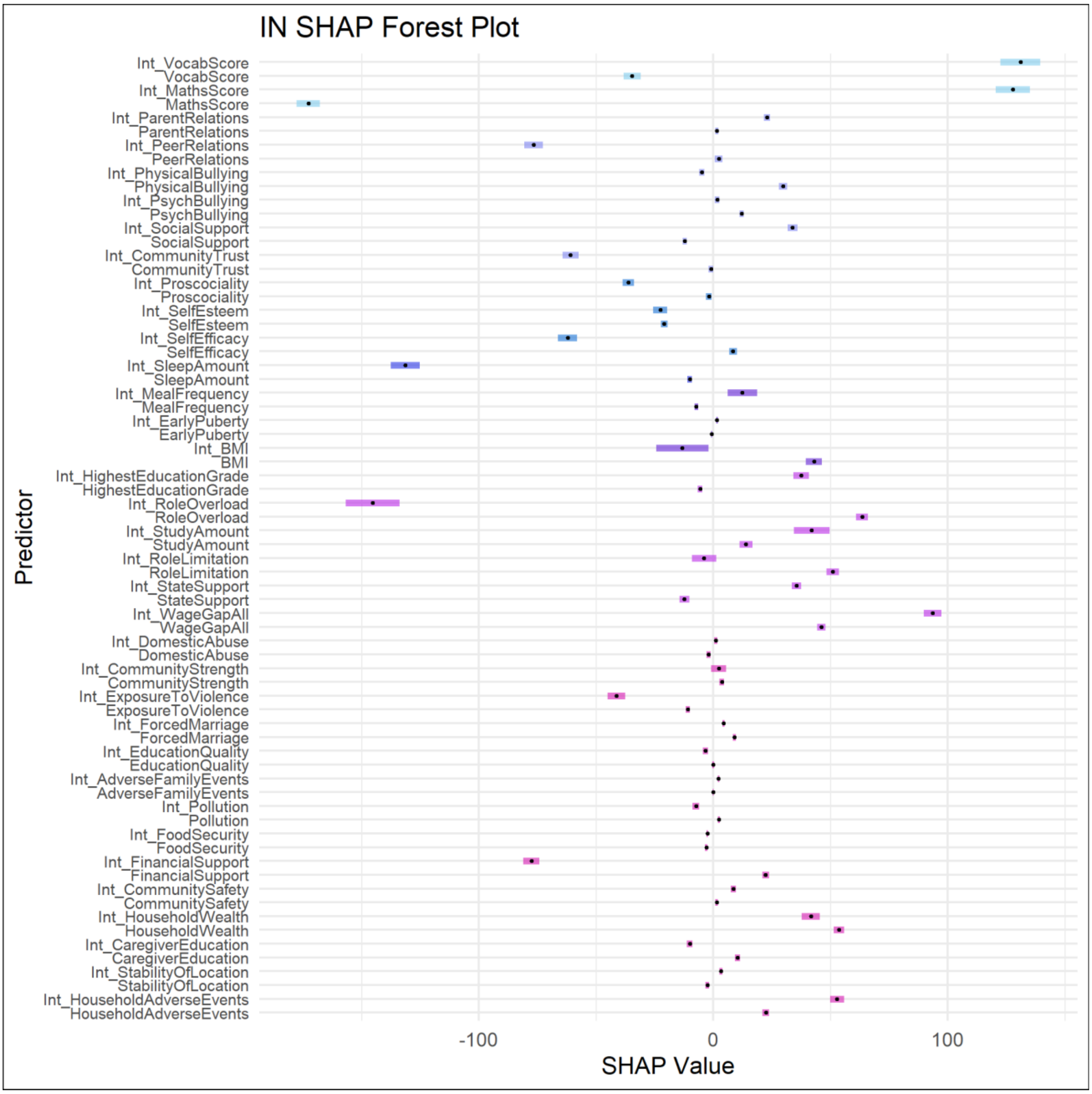
IN SHAP Forest Plot
Predictor
SHAP Value
-100
0
100
Int_VocabScore
VocabScore
Int_MathsScore
MathsScore
Int_ParentRelations
ParentRelations
Int_PeerRelations
PeerRelations
Int_PhysicalBullying
PhysicalBullying
Int_PsychBullying
PsychBullying
Int_SocialSupport
SocialSupport
Int_CommunityTrust
CommunityTrust
Int_Proscociality
Proscociality
Int_SelfEsteem
SelfEsteem
Int_SelfEfficacy
SelfEfficacy
Int_SleepAmount
SleepAmount
Int_MealFrequency
MealFrequency
Int_EarlyPuberty
EarlyPuberty
Int_BMI
BMI
Int_HighestEducationGrade
HighestEducationGrade
Int_RoleOverload
RoleOverload
Int_StudyAmount
StudyAmount
Int_RoleLimitation
RoleLimitation
Int_StateSupport
StateSupport
Int_WageGapAll
WageGapAll
Int_DomesticAbuse
DomesticAbuse
Int_CommunityStrength
CommunityStrength
Int_ExposureToViolence
ExposureToViolence
Int_ForcedMarriage
ForcedMarriage
Int_EducationQuality
EducationQuality
Int_AdverseFamilyEvents
AdverseFamilyEvents
Int_Pollution
Pollution
Int_FoodSecurity
FoodSecurity
Int_FinancialSupport
FinancialSupport
Int_CommunitySafety
CommunitySafety
Int_HouseholdWealth
HouseholdWealth
Int_CaregiverEducation
CaregiverEducation
Int_StabilityOfLocation
StabilityOfLocation
Int_HouseholdAdverseEvents
HouseholdAdverseEvents

*Peru*

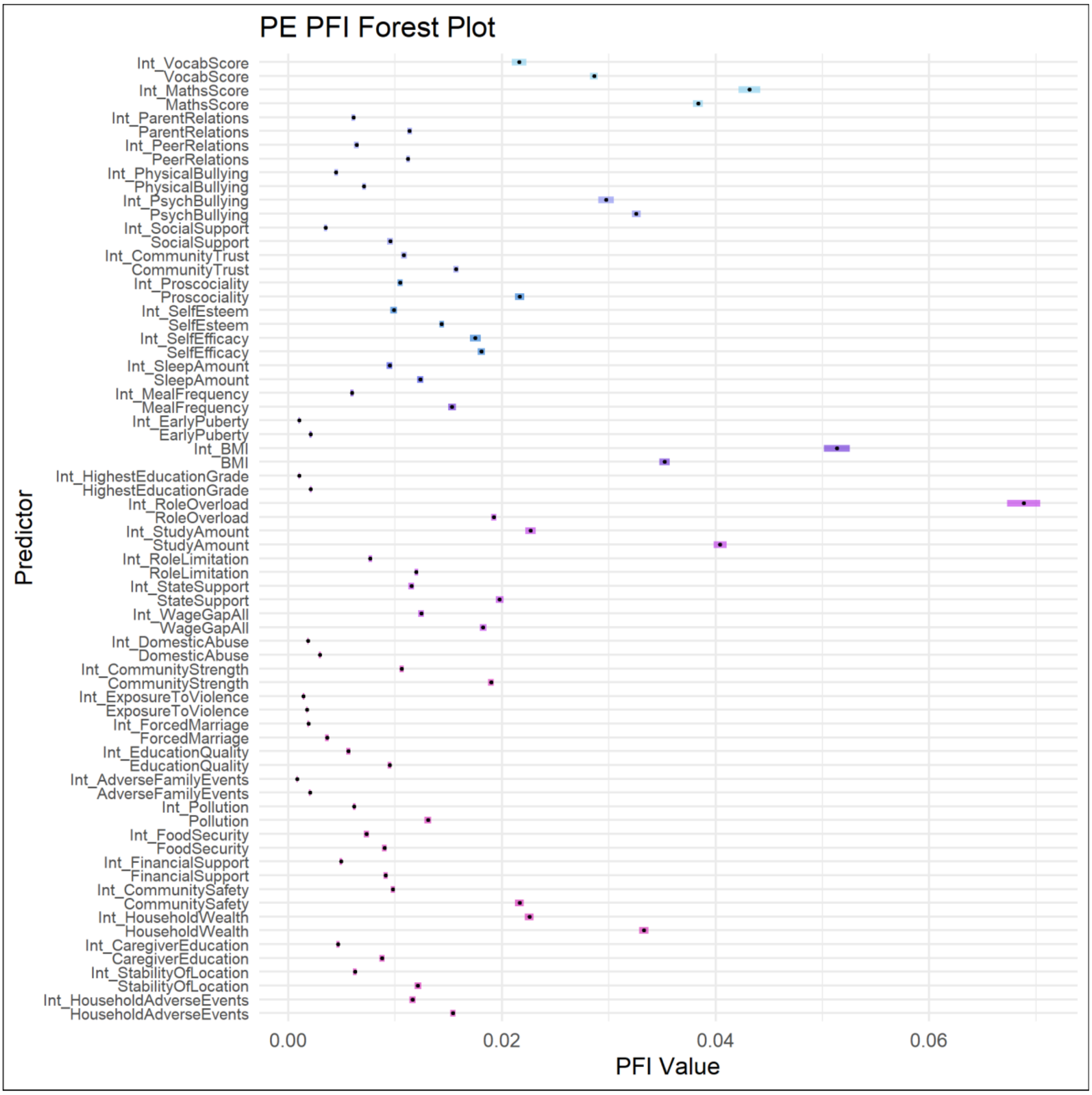
PE PFI Forest Plot
Int_VocabScore
VocabScore
Int_MathsScore
MathsScore
Int_ParentRelations
ParentRelations
Int_PeerRelations
PeerRelations
Int_PhysicalBullying
PhysicalBullying
Int_PsychBullying
PsychBullying
Int_SocialSupport
SocialSupport
Int_CommunityTrust
CommunityTrust
Int_Proscociality
Proscociality
Int_SelfEsteem
SelfEsteem
Int_SelfEfficacy
SelfEfficacy
Int_SleepAmount
SleepAmount
Int_MealFrequency
MealFrequency
Int_EarlyPuberty
EarlyPuberty
Int_BMI
BMI
Int_HighestEducationGrade
HighestEducationGrade
Int_RoleOverload
RoleOverload
Int_StudyAmount
StudyAmount
Int_RoleLimitation
RoleLimitation
Int_StateSupport
StateSupport
Int_WageGapAll
WageGapAll
Int_DomesticAbuse
DomesticAbuse
Int_CommunityStrength
CommunityStrength
Int_ExposureToViolence
ExposureToViolence
Int_ForcedMarriage
ForcedMarriage
Int_EducationQuality
EducationQuality
Int_AdverseFamilyEvents
AdverseFamilyEvents
Int_Pollution
Pollution
Int_FoodSecurity
FoodSecurity
Int_FinancialSupport
FinancialSupport
Int_CommunitySafety
CommunitySafety
Int_HouseholdWealth
HouseholdWealth
Int_CaregiverEducation
CaregiverEducation
Int_StabilityOfLocation
StabilityOfLocation
Int_HouseholdAdverseEvents
HouseholdAdverseEvents
Predictor
0.00
0.02
0.04
0.06
PFI Value

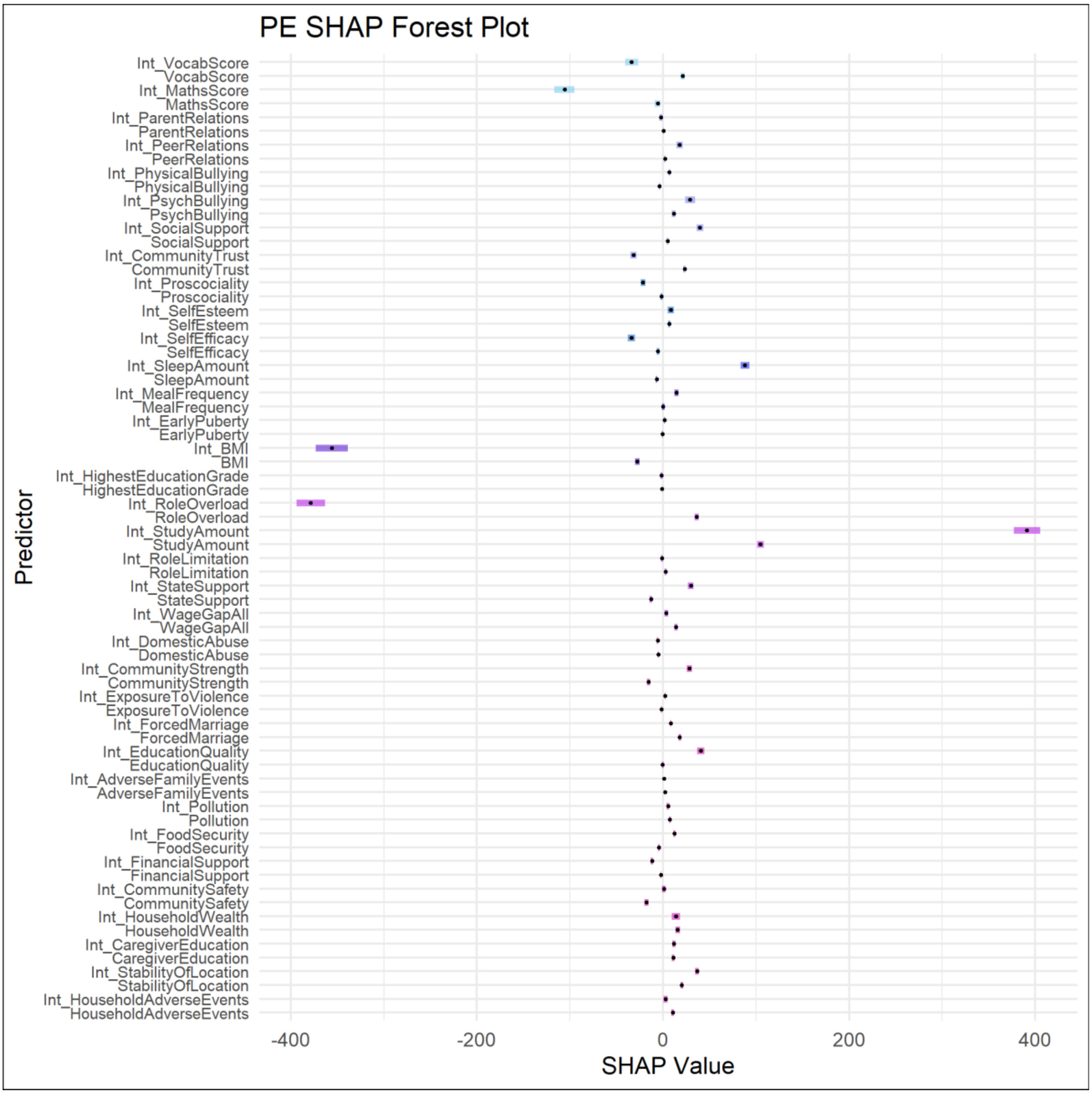
PE SHAP Forest Plot
Predictor
Int_VocabScore
VocabScore
Int_MathsScore
MathsScore
Int_ParentRelations
ParentRelations
Int_PeerRelations
PeerRelations
Int_PhysicalBullying
PhysicalBullying
Int_PsychBullying
PsychBullying
Int_SocialSupport
SocialSupport
Int_CommunityTrust
CommunityTrust
Int_Proscociality
Proscociality
Int_SelfEsteem
SelfEsteem
Int_SelfEfficacy
SelfEfficacy
Int_SleepAmount
SleepAmount
Int_MealFrequency
MealFrequency
Int_EarlyPuberty
EarlyPuberty
Int_BMI
BMI
Int_HighestEducationGrade
HighestEducationGrade
Int_RoleOverload
RoleOverload
Int_StudyAmount
StudyAmount
Int_RoleLimitation
RoleLimitation
Int_StateSupport
StateSupport
Int_WageGapAll
WageGapAll
Int_DomesticAbuse
DomesticAbuse
Int_CommunityStrength
CommunityStrength
Int_ExposureToViolence
ExposureToViolence
Int_ForcedMarriage
ForcedMarriage
Int_EducationQuality
EducationQuality
Int_AdverseFamilyEvents
AdverseFamilyEvents
Int_Pollution
Pollution
Int_FoodSecurity
FoodSecurity
Int_FinancialSupport
FinancialSupport
Int_CommunitySafety
CommunitySafety
Int_HouseholdWealth
HouseholdWealth
Int_CaregiverEducation
CaregiverEducation
Int_StabilityOfLocation
StabilityOfLocation
Int_HouseholdAdverseEvents
HouseholdAdverseEvents
-400
-200
0
200
400
SHAP Value

***Vietnam***

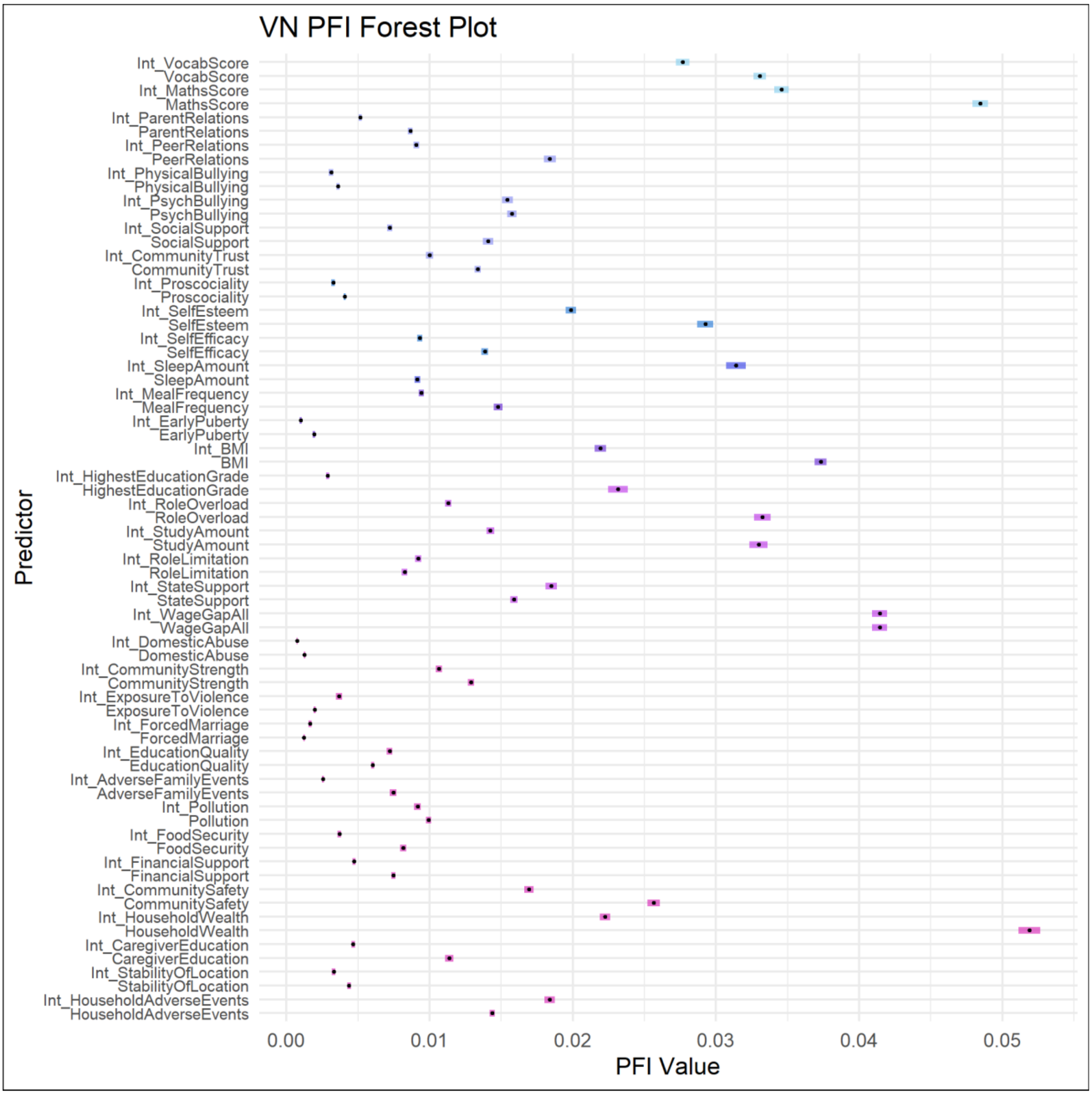

VN PFI Forest Plot
Predictor
PFI Value
Int_VocabScore
VocabScore
Int_MathsScore
MathsScore
Int_ParentRelations
ParentRelations
Int_PeerRelations
PeerRelations
Int_PhysicalBullying
PhysicalBullying
Int_PsychBullying
PsychBullying
Int_SocialSupport
SocialSupport
Int_CommunityTrust
CommunityTrust
Int_Proscociality
Proscociality
Int_SelfEsteem
SelfEsteem
Int_SelfEfficacy
SelfEfficacy
Int_SleepAmount
SleepAmount
Int_MealFrequency
MealFrequency
Int_EarlyPuberty
EarlyPuberty
Int_BMI
BMI
Int_HighestEducationGrade
HighestEducationGrade
Int_RoleOverload
RoleOverload
Int_StudyAmount
StudyAmount
Int_RoleLimitation
RoleLimitation
Int_StateSupport
StateSupport
Int_WageGapAll
WageGapAll
Int_DomesticAbuse
DomesticAbuse
Int_CommunityStrength
CommunityStrength
Int_ExposureToViolence
ExposureToViolence
Int_ForcedMarriage
ForcedMarriage
Int_EducationQuality
EducationQuality
Int_AdverseFamilyEvents
AdverseFamilyEvents
Int_Pollution
Pollution
Int_FoodSecurity
FoodSecurity
Int_FinancialSupport
FinancialSupport
Int_CommunitySafety
CommunitySafety
Int_HouseholdWealth
HouseholdWealth
Int_CaregiverEducation
CaregiverEducation
Int_StabilityOfLocation
StabilityOfLocation
Int_HouseholdAdverseEvents
HouseholdAdverseEvents
0.00
0.01
0.02
0.03
0.04
0.05

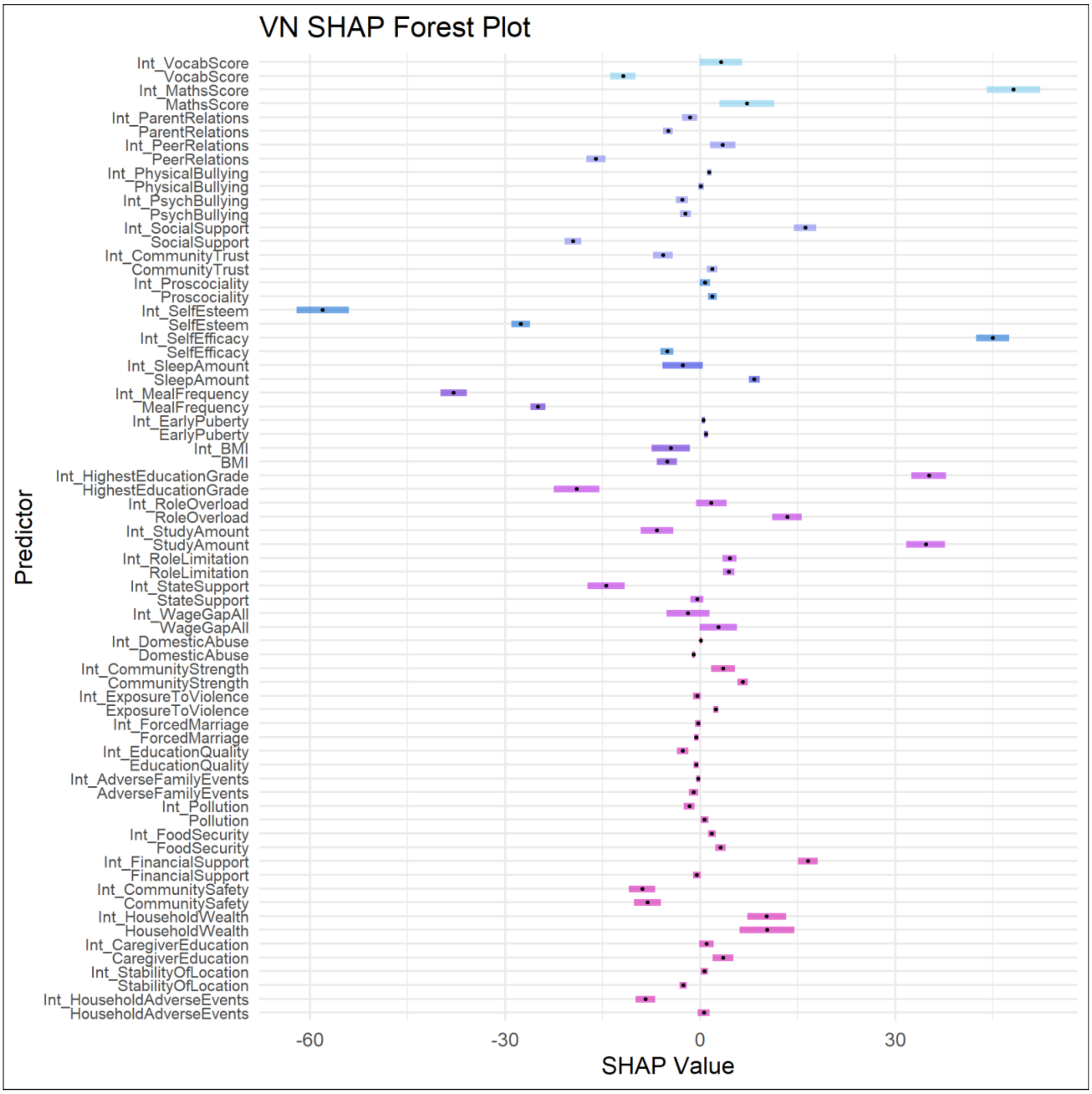
VN SHAP Forest Plot
Predictor
Int_VocabScore
VocabScore
Int_MathsScore
MathsScore
Int_ParentRelations
ParentRelations
Int_PeerRelations
PeerRelations
Int_PhysicalBullying
PhysicalBullying
Int_PsychBullying
PsychBullying
Int_SocialSupport
SocialSupport
Int_CommunityTrust
CommunityTrust
Int_Proscociality
Proscociality
Int_SelfEsteem
SelfEsteem
Int_SelfEfficacy
SelfEfficacy
Int_SleepAmount
SleepAmount
Int_MealFrequency
MealFrequency
Int_EarlyPuberty
EarlyPuberty
Int_BMI
BMI
Int_HighestEducationGrade
HighestEducationGrade
Int_RoleOverload
RoleOverload
Int_StudyAmount
StudyAmount
Int_RoleLimitation
RoleLimitation
Int_StateSupport
StateSupport
Int_WageGapAll
WageGapAll
Int_DomesticAbuse
DomesticAbuse
Int_CommunityStrength
CommunityStrength
Int_ExposureToViolence
ExposureToViolence
Int_ForcedMarriage
ForcedMarriage
Int_EducationQuality
EducationQuality
Int_AdverseFamilyEvents
AdverseFamilyEvents
Int_Pollution
Pollution
Int_FoodSecurity
FoodSecurity
Int_FinancialSupport
FinancialSupport
Int_CommunitySafety
CommunitySafety
Int_HouseholdWealth
HouseholdWealth
Int_CaregiverEducation
CaregiverEducation
Int_StabilityOfLocation
StabilityOfLocation
Int_HouseholdAdverseEvents
HouseholdAdverseEvents
-60
-30
0
30
SHAP Value
